# Examining the cognitive phenotype in Neurofibromatosis 1: a systematic review of functional imaging studies

**DOI:** 10.1101/2025.01.27.25321195

**Authors:** Anna Wild, Maria Tziraki, Caroline Lea-Carnall, Shruti Garg

## Abstract

Neurofibromatosis 1 (NF1) is a rare genetic condition, frequently associated with learning difficulties. Functional neuroimaging techniques have been used to investigate brain mechanisms underlying learning difficulties in NF1, but these remain poorly understood. We conducted a systematic review of functional neuroimaging studies in NF1 from 2000-2024 aiming to develop a neurobiological framework for understanding the NF1 cognitive phenotype. A total of 44 studies were identified as relevant across functional magnetic resonance imaging (fMRI), electroencephalography (EEG), magnetic resonance spectroscopy (MRS), positron emission topography (PET), transcranial magnetic stimulation (TMS) and arterial spin labelling (ASL) techniques. Findings suggest mechanisms underlying learning in NF1 may be characterised as excitation/ inhibition imbalance, dysconnectivity between brain regions and perturbations in neural activation patterns. We suggest, disruptions in GABAergic signalling, beginning in early development, may alter the whole brain function, explaining the differential patterns of brain activity, connectivity, and brain chemistry. As most of the reviewed studies were cross-sectional in nature, future research should focus on longitudinal designs to better understand developmental changes in brain mechanisms and how these processes evolve over time. Finally, it is essential to explore environmental modifiers in shaping the cognitive phenotypes associated with NF1.

## 1 Introduction

Neurofibromatosis 1 (NF1) is an autosomal dominant neurocutaneous condition caused by an inherited or spontaneous mutation of the NF1 gene located on chromosome 17q11.2, encoding for neurofibromin (Ballester et al., 1990; Evans et al., 2010). Clinical manifestations include cafe au lait spots, Lisch nodules, neurofibromas and optic gliomas, as well as an increased susceptibility to the development of malignant tumours (Ratner & Miller, 2015). Cognitive and behavioural function are variably affected, with at least some form of learning difficulty in approximately 70% of cases, although this may be severe in up to 10% of affected individuals (Hyman et al., 2005). Co-occurring difficulties may include attention deficit hyperactivity disorder (ADHD) (30-50%), autism spectrum conditions (30%), intellectual impairment (40%) and emotional and psychosocial problems (40%) (Garg et al., 2013; Mautner et al., 2015; Pride et al., 2013; Pride et al., 2012; van der Vaart et al., 2016). Consequently, these difficulties contribute to poor academic achievement, quality of life, and social functioning (Graf et al., 2006; Huijbregts & de Sonneville, 2011; Hyman et al., 2005). Specific cognitive deficits are in the domains of attentional and executive function, especially in terms of cognitive flexibility and planning (Galasso et al., 2014; Pride et al., 2012), deficits in working memory (Plasschaert et al., 2016; Pobric et al., 2022) and inhibition (Diggs-Andrews & Gutmann, 2013; Huijbregts & de Sonneville, 2011; Plasschaert et al., 2016; Ribeiro et al., 2015).

Animal models of NF1 have elucidated the neurobiological mechanisms resulting from the loss of neurofibromin function which regulates RAS/mitogen activated protein kinases (MAPK) pathway related signalling (Costa et al., 2002). Loss of function mutations of the NF1 gene consequently result in hyperactivity of the RAS signalling cascade, with downstream activation of the MAPK, and mammalian target of rapamycin (mTOR) signalling, impacting γ-aminobutyric acid (GABA) neurotransmission, synaptic plasticity and long-term potentiation (Lobbous et al., 2020; Molina & Adjei, 2006). Subsequently, these models have revealed an important role of GABAergic overactivity and the disrupted GABA/Glutamate homeostasis in explaining NF1 related cognitive difficulties (Costa et al., 2002; Cui et al., 2008; Molosh & Shekhar, 2018; Shilyansky et al., 2010).

Despite mechanistic insights from *Nf1^+/−^* animal models, there has been little progress in treatment discovery, suggesting the neurobiological mechanisms of cognitive dysfunction in NF1 may be more complex in humans. Drugs such as statins which have been shown to inhibit RAS/MAPK activity and reverse the cognitive phenotype in *Nf1^+/−^* mice (Li et al., 2005), have failed to successfully translate to effective therapeutic options for humans (Agouridis et al., 2023). Likewise, other treatments like Lamotrigine, which has also rescued cognitive deficits in *Nf1^+/−^*mice (Omrani et al., 2015), and more recently MEK inhibitors, have shown limited efficacy in improving cognitive outcomes in NF1 (de Blank et al., 2022; Ottenhoff et al., 2024; Walsh et al., 2021). Collectively, this translational failure suggests that animal models may not fully recapitulate the functional complexity of human brain dysfunction.

Functional neuroimaging studies can help us understand the neurobiological basis of cognitive functioning (Liegeois & Elward, 2020). Modern techniques such as functional magnetic resonance imaging (fMRI) and electroencephalography (EEG) allow measurement of neural activity underlying cognitive processing with different spatial and temporal resolutions. Other techniques such as MR spectroscopy (MRS) allow measurement of brain metabolites and positron emission tomography (PET) can measure underlying metabolic activity, blood flow and regional chemical composition. Furthermore, use of non-invasive brain stimulation techniques such as transcranial magnetic stimulation (TMS) allow investigation into mechanisms of neuroplasticity. These functional neuroimaging techniques when combined, allow us to build a comprehensive picture of brain function, identifying causal pathways of symptom emergence in humans for development of treatment for learning difficulties

The aim of this review is to systematically synthesise all functional neuroimaging studies in NF1, integrating findings from functional, metabolic, and neurophysiological research to develop a comprehensive framework for understanding the brain mechanisms underlying the cognitive phenotype in NF1.

## 2 Methods

### 2.1 Eligibility criteria

We included all functional neuroimaging studies in NF1, which were acquired in combination with cognitive tasks. These included fMRI, MRS, PET, TMS, and EEG. Both randomised control trials and observational studies such as cohort, case–control and cross-sectional studies were eligible for inclusion, as long as they reported baseline metrics. Studies were included that reported associations between brain metrics and clinical or cognitive assessments or group comparisons on those metrics. Authors MT and AW assessed records according to the following inclusion criteria:

#### Inclusion Criteria

- Empirical research published in peer-reviewed journals appearing in either preprint or final version, or studies published in conference proceedings or thesis.
- Articles published in English.
- Used validated cognitive tasks or clinical diagnostic tools.
- Used quantitative neurophysiological, metabolic or MRI metrics.
- Involved human participants of any age with NF1 diagnosis

#### Exclusion Criteria

- Used only structural or volumetric imaging methods.
- Case series and reports, owing to the high potential for bias in these study designs.
- Published before the year 2000.

### 2.2. Search Strategy

The review complies with the Preferred Reporting Items for Systematic Reviews and Meta-Analyses (PRISMA) 2020 reporting standards (Page et al., 2021). The guideline was expanded using the Synthesis Without Meta-analysis (SWiM) (Campbell et al., 2020) guideline for data synthesis, but meta-analyses of effect estimates were not possible due to diverse study characteristics. The protocol was prospectively registered on PROSPERO: (CRD42022327084:https://www.crd.york.ac.uk/prospero/display_record.php?RecordI D=327084).

The following databases were searched for references: MedLine (Ovid), PsycINFO (Ovid), PsycArticles (Ovid), Embase (Ovid), Scopus and PubMed, using Medical Subject Headings (MeSH) and keywords defined based on patient intervention comparison and outcome framework (PICO) **(Table 1)**. Identification of key vocabulary was performed using selection of a representative sample of key documents from each method so as to capture all relevant literature. Comprehensive literature searches were carried out from 14^th^ April 2022 to 9^th^ January 2025.

**Table 1.** List of main terms used for our search.

|  | Population | Intervention | Outcome |
| --- | --- | --- | --- |
| **Synonyms and additional free text terms for each aspect** | ‘Neurofibromatosis type 1’ or ‘NF1’ | ‘Magnetic resonance imaging’ or ‘fmri’ or ‘connectivity’ or ‘electroencephalography’ or ‘erp’ or ‘meg’ or ‘transcranial magnetic stimulation’ or ‘tms’ or ‘positron emission tomography’ or ‘pet’ or ‘fnirs’ or ‘arterial spin Labeling’ or ‘asl’ or ‘perfusion’ or ‘spectroscopy’ or ‘mrs’ | ‘attention’ or ‘cogniti*’ or ‘function’ or ‘adhd’ or ‘attention deficit disorder with hyperactivity’ or ‘asd’ or ‘autism spectrum disorder’ or ‘social cognition’ or ‘behav*’ or ‘performance’ |
| **Limits applied** | No age limits |  | Not before 2000<br>Not case reports, clinical research guidelines, meta-analyses or reviews |

Searches were performed by combining the participant search terms, such as ‘neurofibromatosis type 1’, and intervention search terms ‘magnetic resonance imaging’. The final set of search terms included cognitive, social and behavioural outcomes in individuals with NF1, namely ‘cogniti*’. The search strategy was adapted for each database and included a combination of MeSH and terms to capture all available records on the topic (for the full search strategy of each database **(Supplementary File 1)**. In addition, we manually searched the reference lists of included studies to identify further relevant publications.

### 2.3 Identification of studies and selection process

A total of 4306 records were initially identified across all databases by reviewers MT and AW and imported to the systematic review data management platform Covidence (https://www.covidence.org/; accessed January 2025). Following automated removal of duplicates, 646 remained for title and abstract screening which subsequently left 64 studies. Full text screening, and data extraction were all performed independently by reviewers (AW, SG and CLC). Disagreement was resolved by discussion and studies were included following 100% consensus. One study was added by manual searching of the references (Bluschke et al., 2017a). The final number of studies included in the review was 44. The screening process is summarised in the flowchart **(Fig. 1)**.

**Fig 1.**
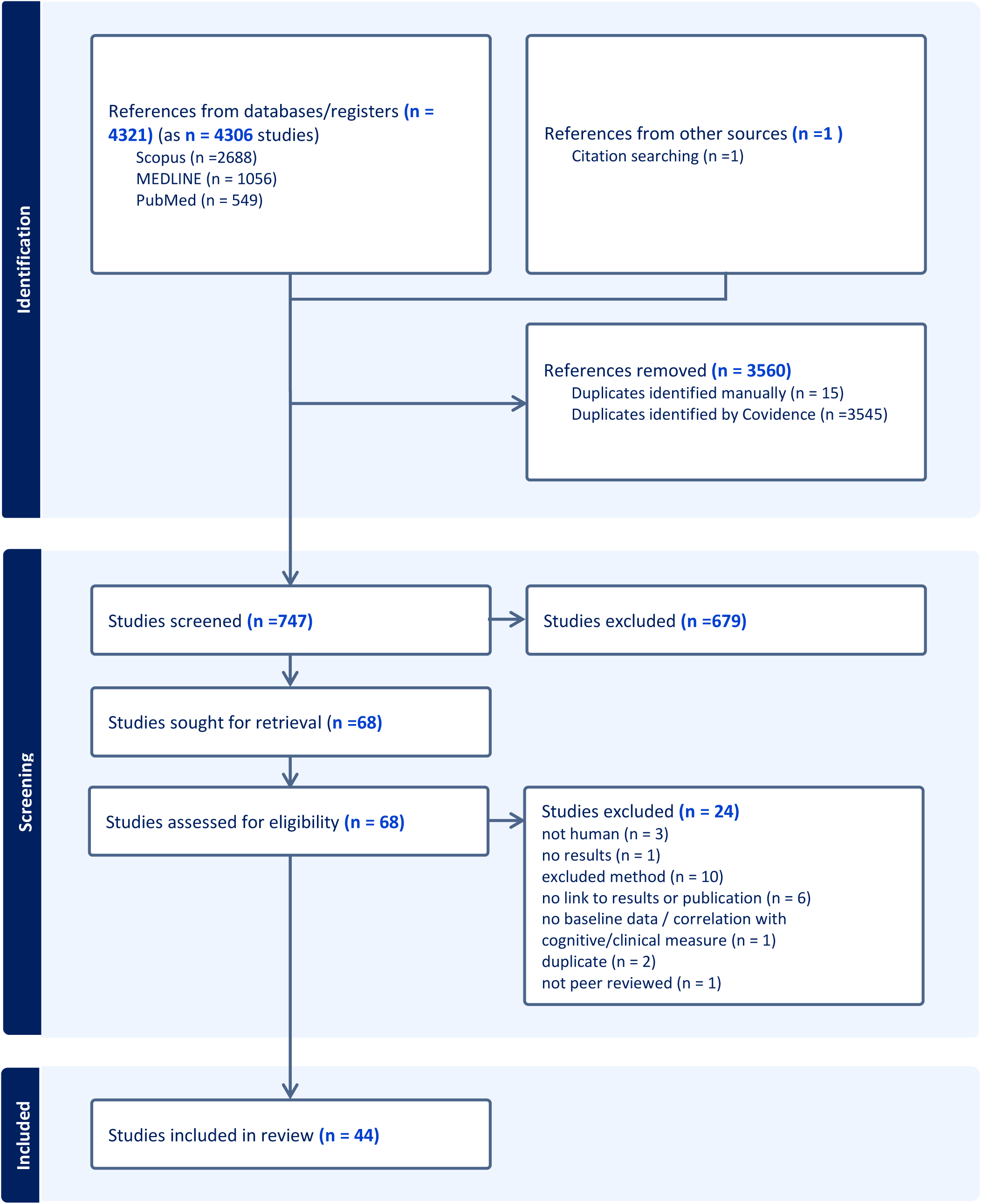
PRISMA study selection flow chart (Page et al., 2021).

### 2.4 Data collection and extraction

A data extraction form was developed **(Supplementary File 2)**, which helped to guide the recording of comprehensive information about the reviewed studies and facilitate interpretation of findings, as well as quality assessment. The data extraction form was piloted twice by MT on a random selection of 10 included resources. Extractions were checked for consensus, and the form was revised according to feedback and the final version was implemented using Extraction 2.0.

We extracted the following information from all included studies: lead author, publication year and country, study design, sample characteristics (total number of recruited and final sample tested, gender, handedness), recruitment strategy and participant preparation procedure (diagnostic criteria for inclusion of NF1 sample and selection criteria for control groups, exclusion criteria for NF1 and control samples at each research stage, matching strategy, mean, standard deviation and age range), brain assessment methods and quantitative variables (brain regions, measures of assessment for each variable of interest, type of statistical model and significance level, comparison of each variable between groups, correlations with quantitative cognitive or behavioural measures, effect direction, and methods used to control confounding and missing data), behavioural task or clinical assessment or questionnaire, rater and group comparisons reported on those measures **(Supplementary File 3)**.

### 2.5 Quality assessment

Quality appraisal was embedded in the data extraction process, and selection of the sections and questions to include in Extraction 2 was guided by the guidelines stated in the Cochrane Handbook for Systematic Reviews of Interventions (Li et al., 2024a). The total count for each checklist described below was used to prioritise studies for textual description and ranking of studies in each table included in the results section, to reflect the quality assessment we conducted. Authors independently evaluated study quality and recorded key data using a standardised data extraction form (Extraction 2, Covidence) which included the following sections:

1. A modified version of Joanna Briggs Institute (JBI) critical appraisal checklist was used for checking the overall study methodology (Munn et al., 2020).
2. For EEG studies, reporting of recording characteristics and instruments, stimulus timing parameters, preprocessing and artifact rejection techniques, and statistical analyses were checked (Keil et al., 2014).
3. For TMS studies, reporting of recording characteristics and instruments, stimulation, pulse and timing parameters, data preprocessing and threshold, hotspot, and MEP determination, and data analysis methods and type of outputs were checked (Chipchase et al., 2012).
4. For fMRI, PET and ASL studies, reporting of mock scanning protocol and sedation, hardware system description and acquisition parameters, quality control and motion parameter checks, data preprocessing and intra and intersubject registration methodology, smoothing and artifact removal use, task design specification, statistical modelling and inference description were checked (Nichols et al., 2017; Poldrack et al., 2008)
5. For MRS studies, reporting of hardware system description and acquisition parameters, water suppression and quality control methods, triggering and or motion correction method, data analysis methods and type of outputs were checked (Lin et al., 2021).

The full data extraction sheet can be found in **Supplementary File 2**.

### 2. 6 Data synthesis

Due to the expected heterogeneity between the studies in terms of the age of the participants and method of brain assessment, no meta-analysis of the quantitative metrics was planned. Instead, a narrative review approach was used to best deal with the methodological variability. Hence, we used non-quantitative synthesis; the main criteria used for reporting were directness in relation to the review question and score on data extraction, which reflected the quality of the study design and reporting of the methodology. We then pooled results across studies in cases where a similar methodology and/or outcome measure was reported. Specifically, Synthesis Without Meta-analysis (SWiM) (Campbell et al., 2020) reporting guidelines helped guide the narrative review synthesis, as meta-analyses of effect estimates were not appropriate, due to diverse study characteristics. SWiM elaborates on PRISMA items 14 and 21, using a 9-item checklist covering key features of data synthesis.

## 3 Results

The studies were grouped by intervention type as follows (in line with SWiM criteria):

### 3.1 Studies using task-based and resting-state functional brain imaging techniques

**Definition Box 1.** fMRI terms used in the text.

|  |  |
| --- | --- |
| Functional connectivity (FC) | The temporal correlation in BOLD activity between spatially remote regions – different to effective connectivity which refers to a causal interaction. Can be divided into model based (seed-based correlation analysis) and model free methods for computing FC based in |
|  | decomposition (e.g. principal component analysis) and clustering principles. |
| Static FC | Averaged FC calculated over the entire scan |
| Dynamic FC | Considers fluctuations in connectivity over time by segmenting the acquisitions into windows of time and calculating FC per window. |
| Global correlation analysis | Average correlation between seed regions/ROIs with all other brain regions |
| Local correlation analysis | How voxels in a small region are correlated with each other e.g. regional homogeneity |
| Fraction of amplitude of low frequency fluctuations (fALFF) | Quantifies the power of low-frequency fluctuations (<0.1HZ) in the BOLD signal and provides information about the intensity of spontaneous neuronal activity. fALFF provides a normalised quantity that it is thought to reflect local activity at rest |
| Graph theory | Computational technique modelling the brain as a network of nodes connected by edges. Allows insight into organisation of the network |
| Number of edges | The number of connections between nodes. |
| Modularity | A measure of the strength of the division of a network into communities or clusters. A community is a group of nodes that are more densely connected to each other than to nodes outside the group. |
| Degree centrality | A measure of the importance or influence of a node in a network based on the number of connections it has. The degree centrality of a node is defined as the fraction of nodes it is connected to in the network. |
| Global efficiency | A measure of how efficiently information can be exchanged across the network. The global efficiency of a graph is the average of the reciprocals of the shortest path lengths between all pairs of nodes. |
| Participant coefficient | Measures how evenly a node is connected to different communities in the graph. It provides insight into whether a node acts as a connector across communities. |

(Farahani et al., 2019; Tomson et al., 2015; Zou et al., 2008)

#### 3.1.1 Resting-state fMRI

Eight studies examined FC in resting-state fMRI data. Mennigen et al. (2019) investigated static, dynamic, and higher-dimensional properties of FC in adults with NF1 and typically developing controls. Static FC measurements found hypoconnectivity between frontal cognitive control areas and the cerebellum, and the latter also exhibited less anticorrelation with the posterior middle temporal gyrus in NF1. Hyperconnectivity within the frontal cognitive control areas was also evident independent of IQ (Mennigen et al., 2019). Dynamic FC analyses suggested NF1 participants spent more time in a state of whole brain hypoconnectivity relative to TD controls with no effect of IQ or ADHD (Mennigen et al., 2019). The pattern of hypoconnectivity is like that seen in psychosis and the authors speculate whether this might be a nonspecific indicator of overall brain health.

Corticocortical and corticostriatal pathways were investigated in two studies. Baudou et al. (2022) investigated cortico-striatal FC and its association with procedural motor learning in a sample of children between 8-12 years old. The authors found higher connectivity in the cortico-striatal regions mapping onto the right angular gyrus. Despite this difference not relating to procedural motor learning performance, it was associated with general motor abilities such that increased connectivity was evident in participants with poorer motor skills. Similarly, Shofty et al. (2019) found reduced anterior-posterior FC in the default mode network, reduced FC between the striatum and frontoparietal networks and increased striatal FC within the limbic network in NF1 (Shofty et al., 2019).

FC and its association with cognitive and social impairments were investigated by Loitfelder et al. (2015). Increased FC was observed in the NF1 group between left anterior cingulate cortex and left amygdala with the insular cortex and frontoparietal operculum. These connectivity differences showed associations with reduced social, cognitive and behavioural function. None of the group differences were associated with IQ. The authors suggest that increased FC contributes to dysfunction rather than acting as a compensatory increase.

Yoncheva et al. (2017) examined training effects of a computerised working memory program on resting-state fMRI. The intervention decreased fALFF in the cerebellum and thalamus. Post-intervention local synchrony, measured via regional homogeneity (a measure of local intrinsic synchrony), was enhanced in areas responsible for attentional allocation during visual processing (left occipital/fusiform gyrus) and decreased in the right superior frontal sulcus. Task performance increased on speed and accuracy measures (Yoncheva et al., 2017). Chabernaud et al. (2012) investigated the effects of 12 weeks lovastatin treatment in adolescents with NF1 on FC using a seed-based correlation analysis. After 12 weeks of Lovastatin treatment, the authors reported local FC reduced and long-range FC increased in the dorsomedial prefrontal cortex and posterior cingulate cortex clusters, a pattern of FC seen in controls. Local FC decreased in restrosplenial, temporal pole and ventromedial prefrontal cortex clusters.

Nemmi et al. (2019) used T1W structural imaging, diffusion weighted and resting-state fMRI. Using machine learning methods, the authors found that structural imaging, particularly mean diffusivity, performed better at discriminating the NF1 from the control group compared to the functional metrics. The authors investigated local and global as a static FC measure along with fraction of amplitude of low frequency fluctuations (fALFF). Global FC and fALFF could not discriminate between NF1 and controls, although local FC analysis could discriminate between the groups, although were not as good as the structural metrics at discriminating groups. The authors suggest more nuanced measures of connectivity such as graph theory metrics may help to better discriminate between NF1 and controls.

Tomson et al. (2015) also investigated resting-state FC differences in NF1 versus TD controls, comparing number of edges and modularity clustering patterns with other traditional graph theory metrics in Box 1 (degree, global efficiency, modularity coefficient and participation coefficient) (Tomson et al., 2015). Similar to Shofty et al. (2019), the NF1 group had reduced long range anterior-posterior connectivity, as well as weaker bilateral edges and altered modularity clustering relative to controls. Modularity results identified 19 pairs of nodes in visual and default mode networks that exhibited greater clustering in controls than NF1. However, traditional graph theory metrics did not differ between groups, indicative of the groups having similar global functional network organisation.

In summary, findings of resting-state FC studies suggest increased local connectivity within frontal regions, reduced long range connectivity and overall whole brain hypoactivity (Loitfelder et al., 2015; Mennigen et al., 2019; Shofty et al., 2019; Yoncheva et al., 2017).

**Table 2.**
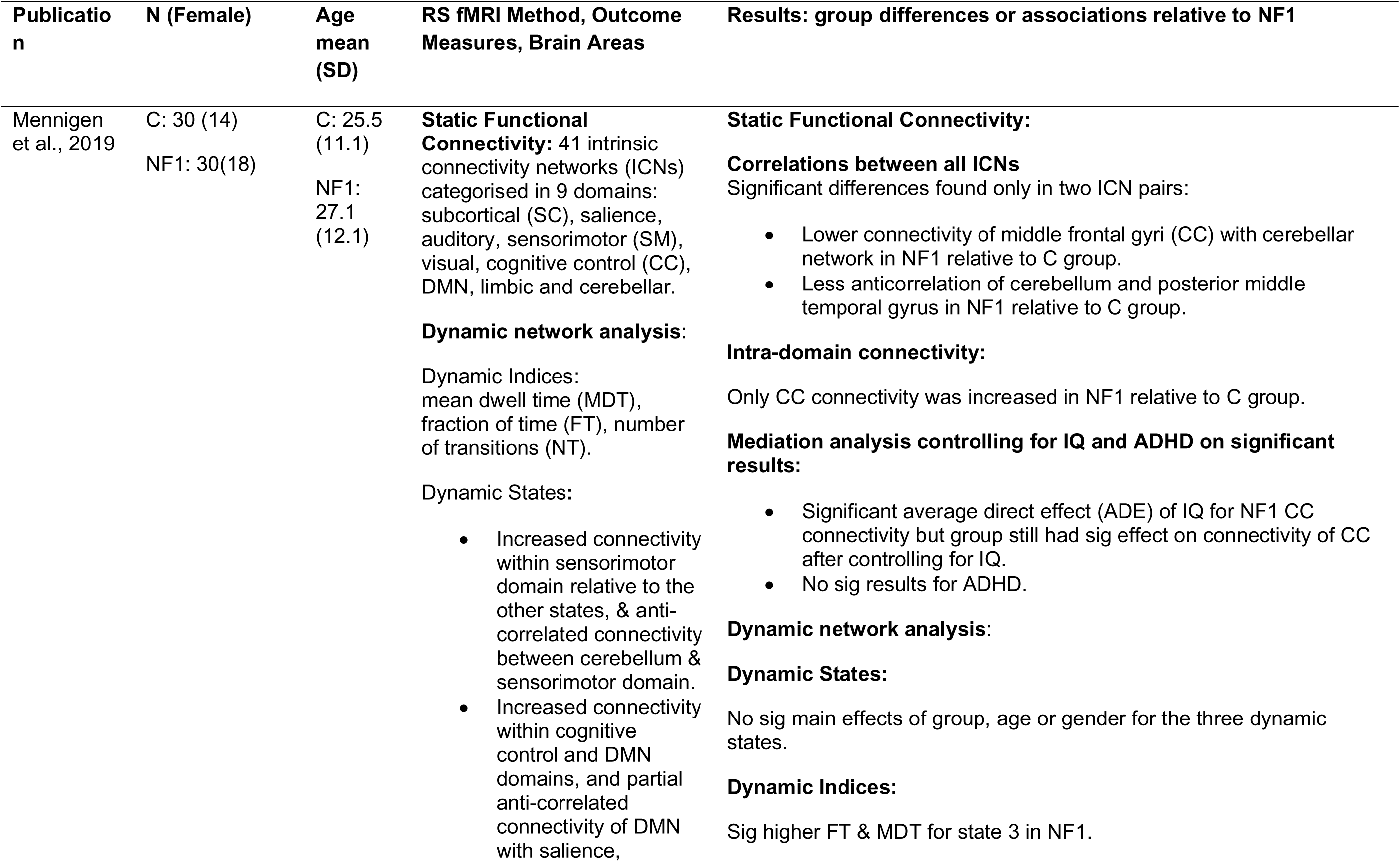
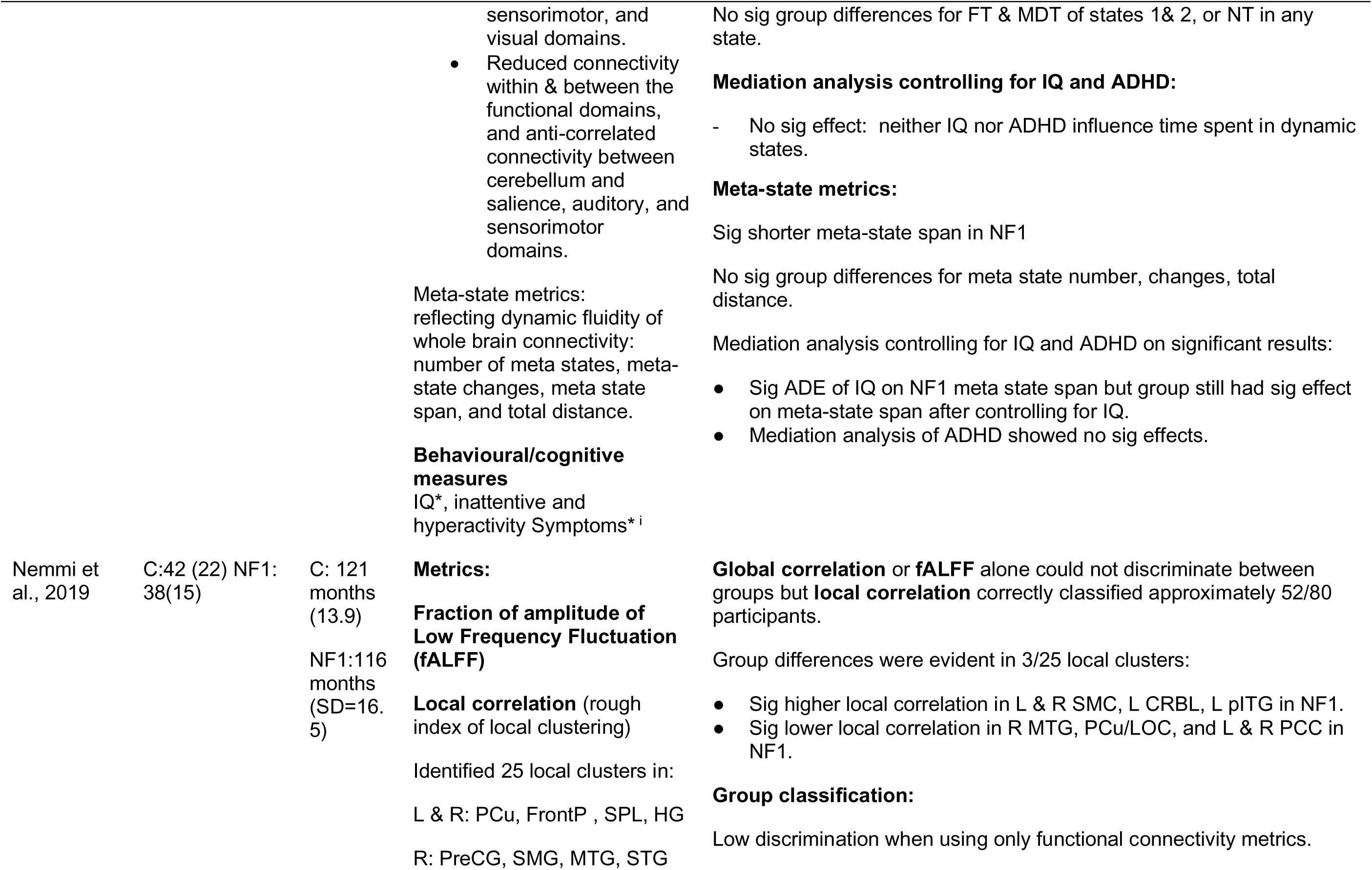
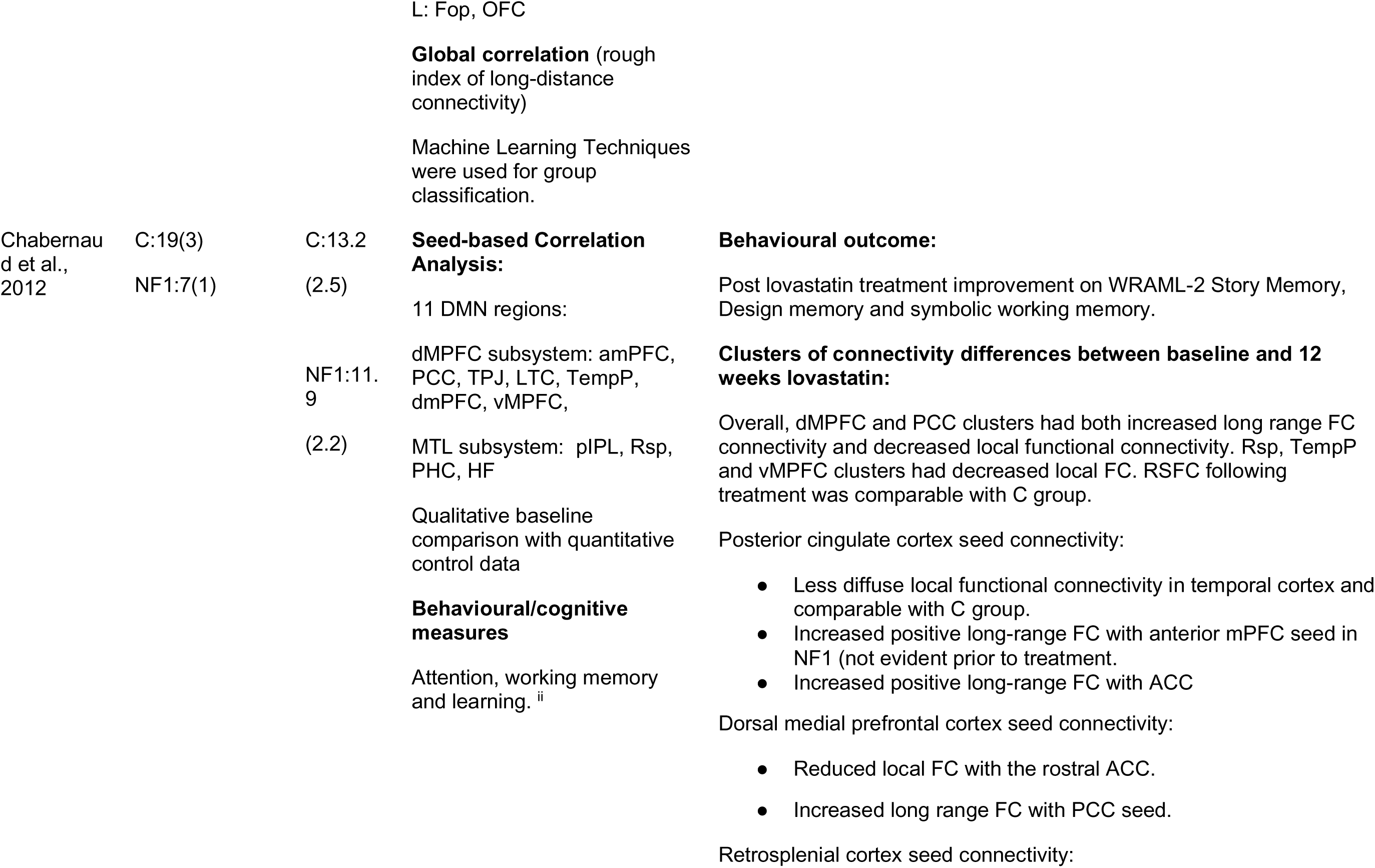
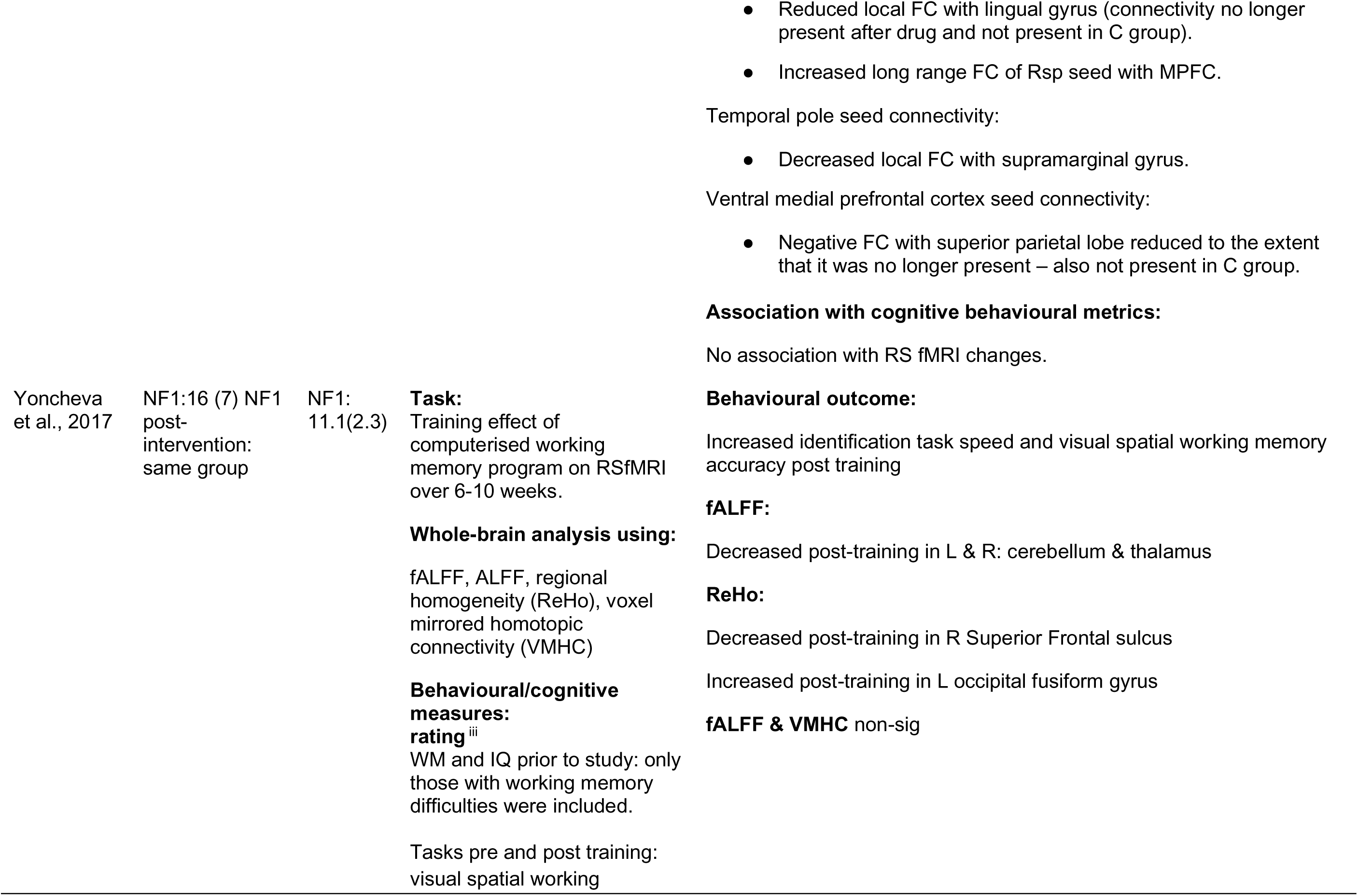
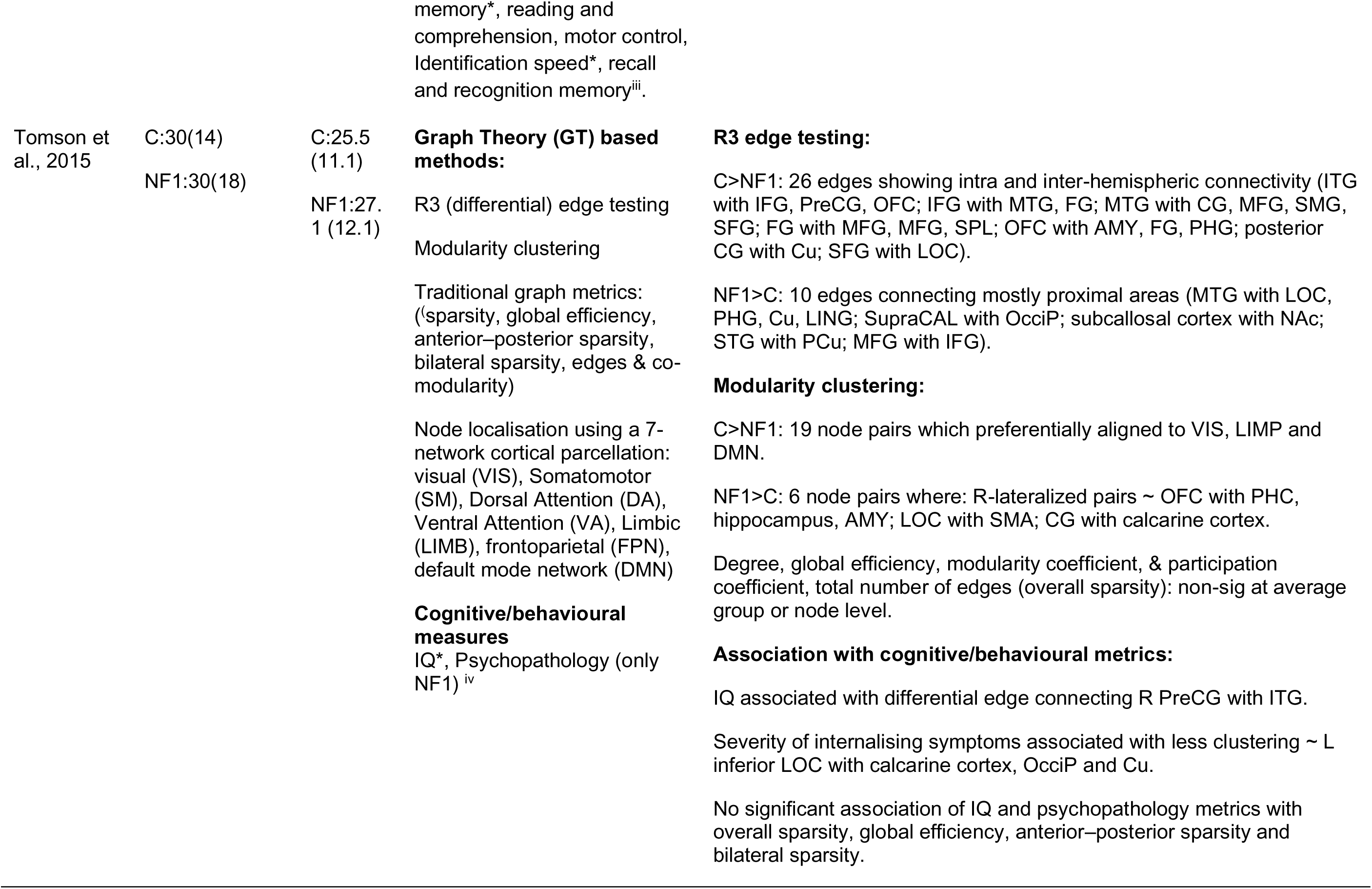
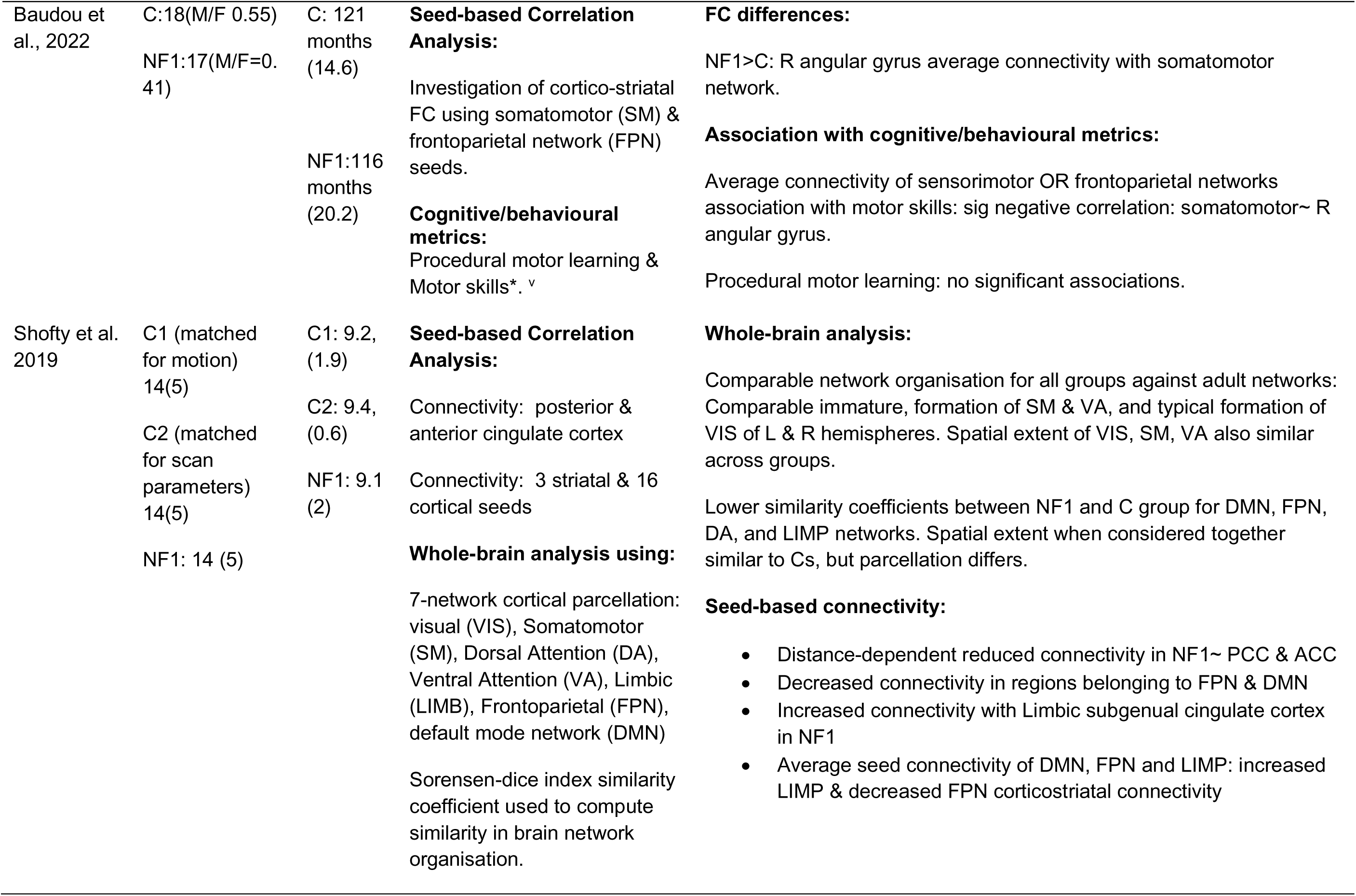
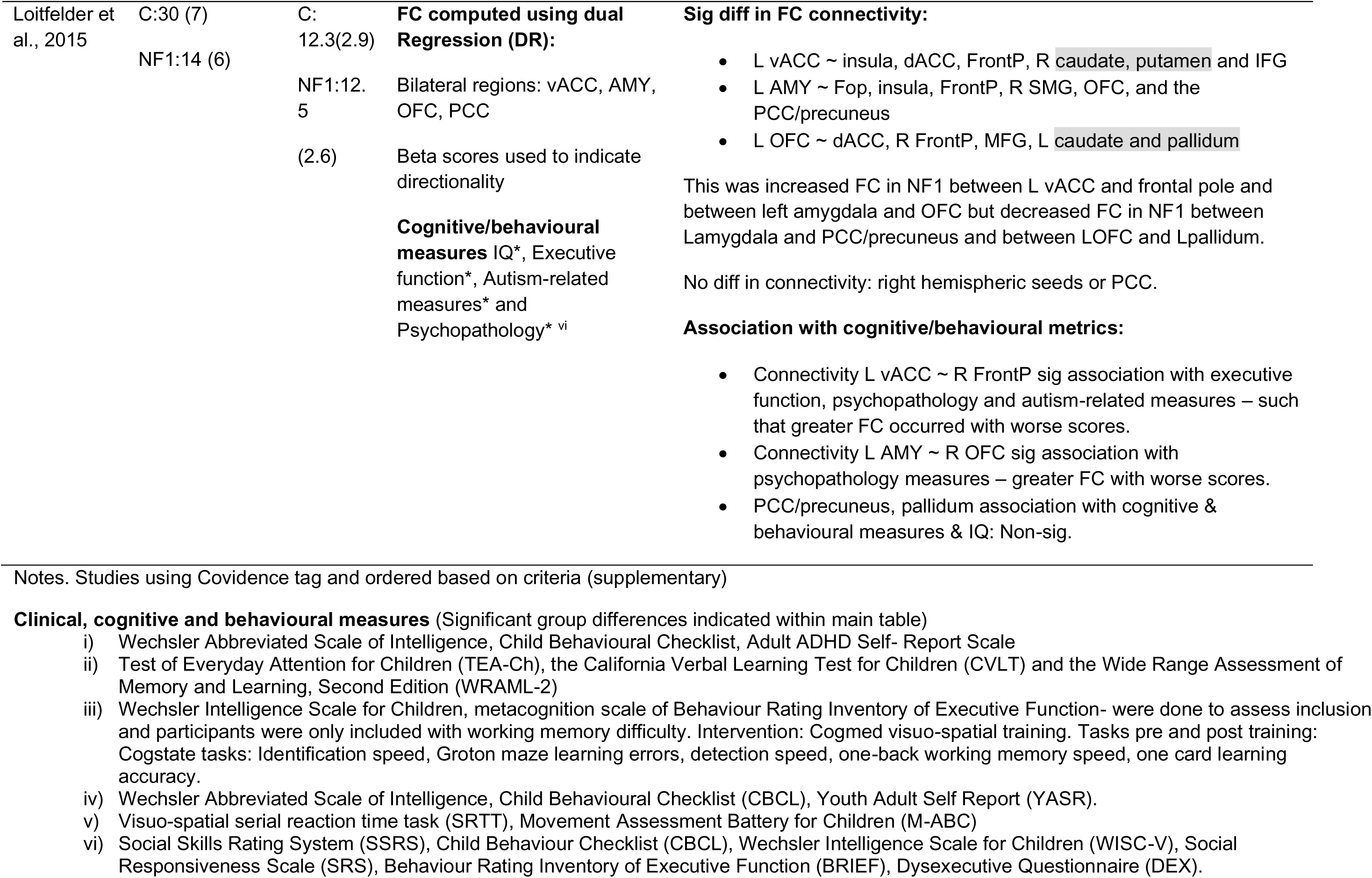
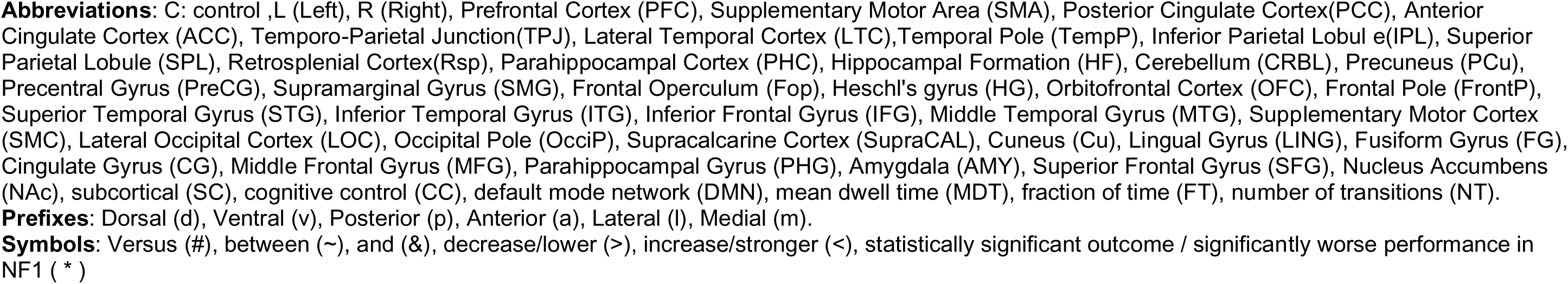
Resting-state fMRI studies.

#### 3.1.2 Task fMRI

##### Cognitive control

Three studies investigated cognitive control using tasks relating to attention and inhibition. Overall, activation appeared to be reduced in NF1 with differences in neural activation patterns compared to controls (Jonas et al., 2017; Pride et al., 2017; Pride et al., 2018). Pride et al. (2018) used an auditory oddball paradigm to examine neural activation patterns of attentional networks in NF1 versus controls and found that children with NF1 show unilateral, and less extensive task-based activation, mainly in the ventral attentional networks. Whole brain analysis revealed right insula activation during the target versus non target condition in controls but not NF1. In addition, reduced activation in the right temporo-parietal junction was associated with increased ADHD traits, but only in controls. In contrast, reduced anterior cingulate cortex activation was associated with poorer ability to control attention in the NF1 group. The authors suggest that there is a dysfunctional ventral attention stream, causing impaired processing of bottom-up information that consequently promotes easier distraction and poor cognitive control (Pride et al., 2018).

Neural activation patterns of response inhibition also differ in NF1 (Jonas et al., 2017; Pride et al., 2017). Pride et al. (2017) focused on the evaluation of response inhibition-related brain networks during a classic Go/No-go paradigm. Region of interest analysis showed NF1 had less activation during response inhibition in the inferior frontal gyrus, inferior occipital gyrus, fusiform gyrus and pre-supplementary motor area. These differences were noted even though the groups were found to have comparable task performance. Interestingly, reduced right inferior frontal gyrus activation in NF1 was also associated with faster response times and thus greater impulsivity as well as being associated with reduced sustained attention (Pride et al., 2017).

Similarly, children and adolescents with NF1 also showed atypical activation of the neural substrates of reward-related decision making, which is intrinsically connected with inhibitory control (Jonas et al., 2017). Jonas et al. (2017) probed the neural circuitry of reward (decision making) using a gambling task in children with NF1 compared to controls. Controls made more risky decisions than the NF1 group. Significant clusters of decreased task-related activity were observed in NF1 versus controls when participants were making safe (supramarginal gyrus and angular gyrus) and risky (regions including the paracingulate cortex and anterior cingulate cortex) choices. Furthermore, the NF1 group did not show the typical increase in activity of the striatum during positive feedback processing, a cognitive process reliant on prefrontal-striatal circuits. Overall risk-taking behaviour triggered an opposite pattern of activation in the two groups; a higher risk-tendency in controls resulted in reduced posterior cingulate cortex and frontal pole activation, whereas in NF1 the same regions showed higher activation with higher risk-tendency. Prospect of reward did not improve inhibitory control in NF1 (Burton et al., 2021), indicating that the poor functioning of this reward circuit in NF1 and subsequent lack of positive feedback may be contributing to poor cognitive control. There was no influence of IQ or ADHD on risky decision making in either group (Jonas et al., 2017).

Finally, motor control was investigated by Silva et al. (2018) using a finger tapping task whereby participants were instructed to tap both fingers either simultaneously or alternating. Controls exhibited greater activation in extrapyramidal motor regions (putamen, red nucleus, medial prefrontal cortex and cerebellum) and had better tapping time precision than NF1.

##### Working memory

Working memory was investigated in 3 studies using visuospatial working memory tasks (Garg et al., 2022; Ibrahim et al., 2017; Shilyansky et al., 2010). The prefrontal working memory circuitry was consistently hypoactive in NF1, including the left dorsolateral prefrontal cortex and right intra parietal sulcus (Ibrahim et al., 2017) and frontal parietal and striatal regions (Shilyansky et al., 2010). Moreover, lower right dorsolateral prefrontal cortex activation was associated with worse task performance (Shilyansky et al., 2010), emphasising the lack of efficiency of the prefrontal circuit. Similarly, Ibrahim et al. (2017) found that as the task became more difficult, the NF1 group exhibited a more diffuse brain activation pattern. This did not occur in controls, who instead showed increased difference in activation due to task difficulty in the right lateral occipital cortex compared to NF1. This is likely a result of reduced efficiency of working memory circuitry and therefore the need to recruit more diffuse regions in NF1 as the task becomes more difficult.

Garg et al. (2022) compared active versus sham non-invasive brain stimulation in children with NF1. In a single-blind cross over trial NF1 participants received transcranial direct current stimulation (tDCS) whilst performing an n-back working memory task. The NF1 group aged 11–17 years exhibited extensive task-dependent neural activation in regions recruited for working memory (bilateral prefrontal cortex, intraparietal sulcus and pre supplementary motor area) during the 2-back versus 0-back task There was no reported comparison of the NF1 group with a control group in this study.

##### Visual and phonological processing

Two studies investigated visual spatial process in in NF1. Billingsley et al. (2004) investigated neural substrates of visuospatial abilities using a mental rotation task and found that NF1 participants specifically recruited areas such as the medial temporal gyrus and lateral occipital cortex, whereas the controls recruited areas such as the inferior frontal gyrus and superior temporal gyrus, to reach comparable levels of performance. The authors suggest that alternative regions are recruited in NF1 to compensate for inefficiencies in typical visuospatial processing pathways. Likewise, Clements-Stephens et al. (2008) examined neural activity associated with visuospatial processing using the Judgement of Line orientation task. The NF1 group tended to recruit left hemispheric regions as opposed to the right hemisphere in controls, further suggesting altered neural recruitment processes.

Visual processing was investigated by Violante et al. (2012) using two distinct stimuli to evoke the magnocellular and parvocellular pathways, finding reduced activation of the visual cortex for both types of low-level visual stimulation in NF1. These differences did not covary with age and were similar across adults and children. Outside the visual areas, the magnocellular-tuned stimulus produced neural activation in anterior brain areas (anterior cingulate cortex, retrosplenial cortex and medial prefrontal cortex), whereas the control group showed reduced activation. The authors suggest that these anomalous activation patterns in NF1 may be due to failure of default mode network deactivation during task conditions. Alternatively, as brain development is altered in NF1, brain activation differences may be due to a different pattern of task relevant regions all together.

Further supporting the altered recruitment patterns in NF1, Billingsley et al. (2003) examined the regional hemodynamic activity associated with phonological processing in adolescent NF1 and control participants. Both the auditory and visual phonological tasks elicited relatively higher recruitment of regions such as the medial occipital cortex, lateral occipital cortex and medial temporal gyrus in NF1 participants relative to controls. Poorer phonological auditory task performance was associated with greater signal change in the right superior temporal gyrus in NF1. Participants with NF1 also showed greater activity bilaterally for the auditory task in the inferior frontal gyrus relative to parietal cortices, but controls showed an equivalent degree of activity in these regions.

**Table 3.**
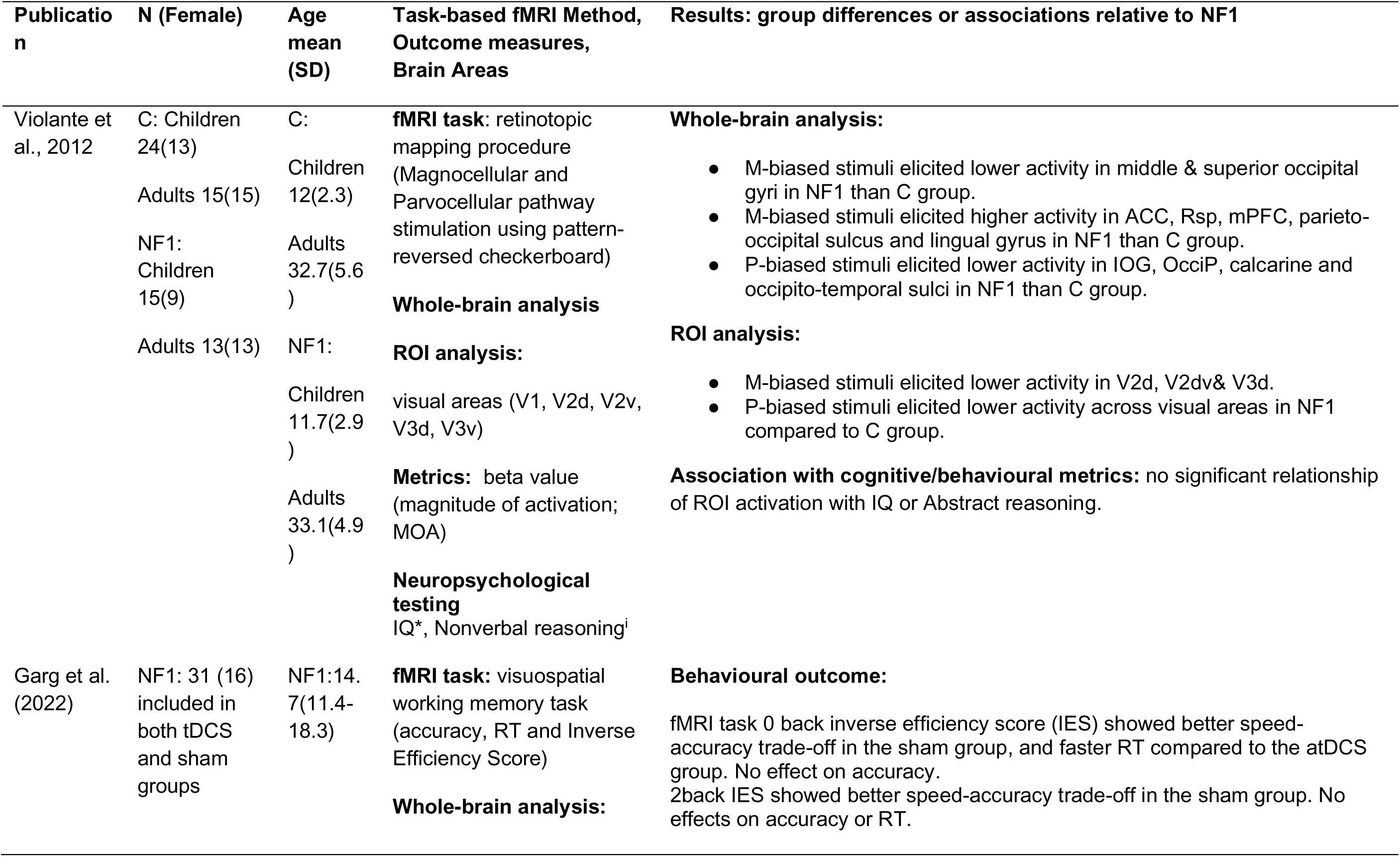
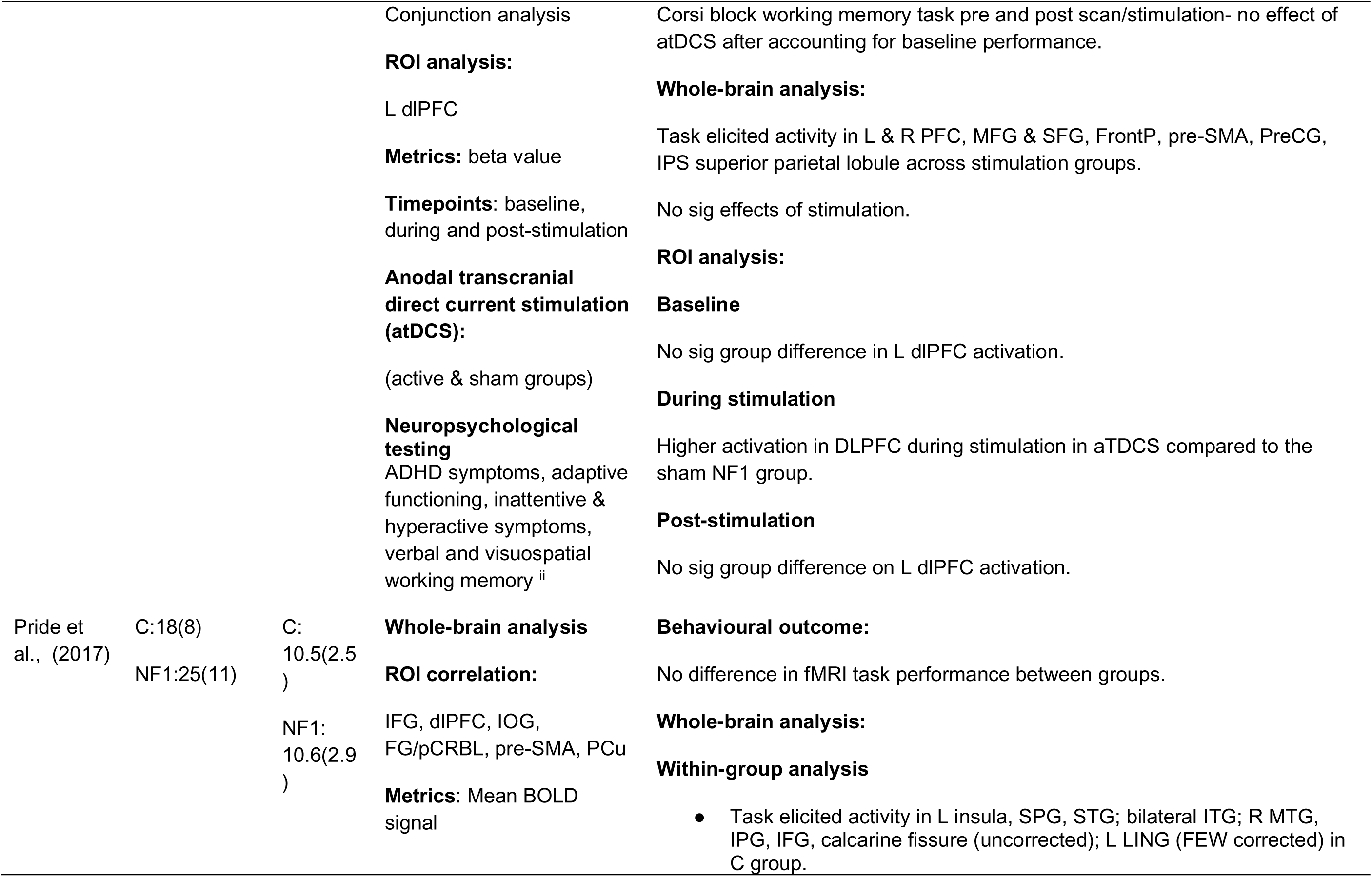
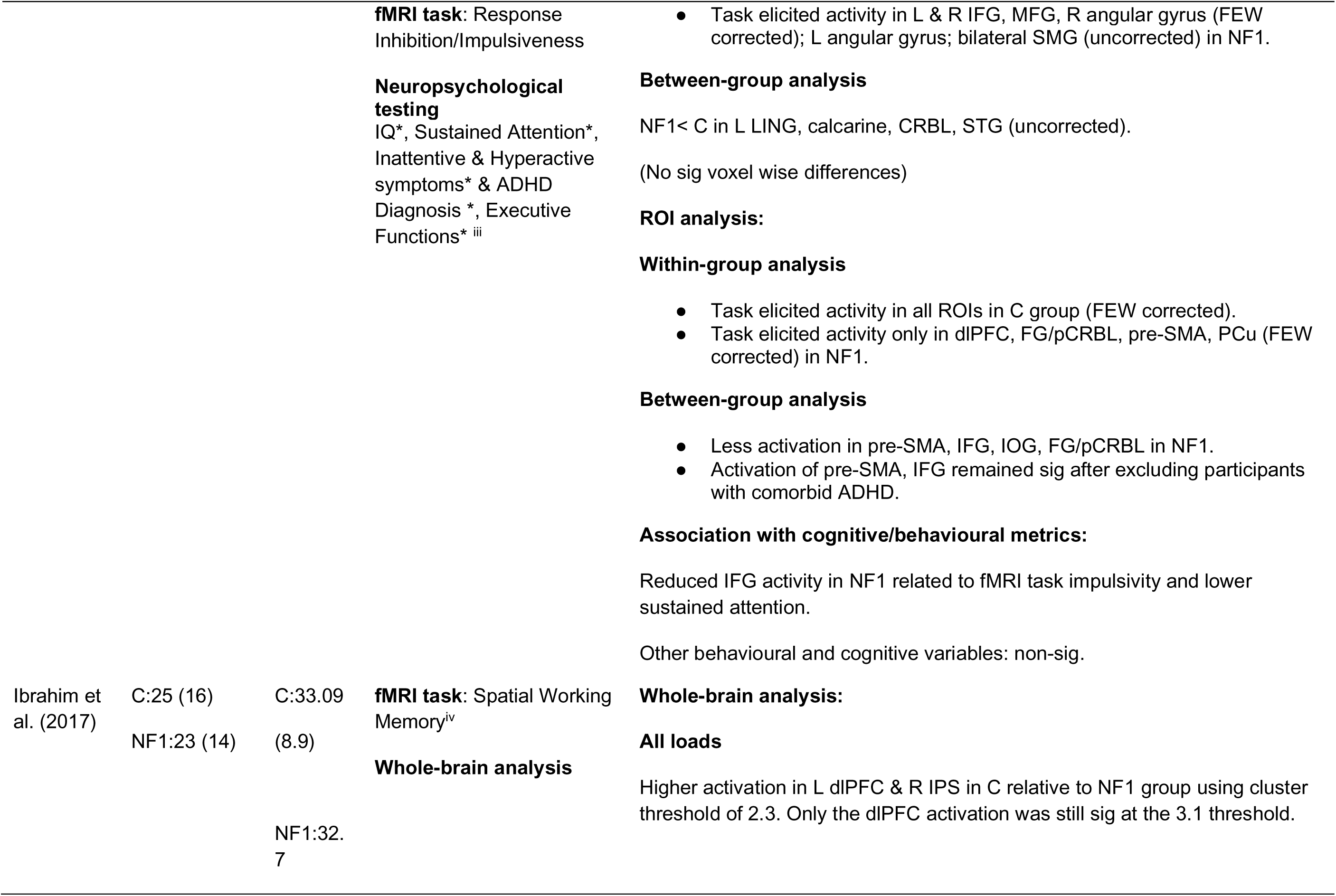
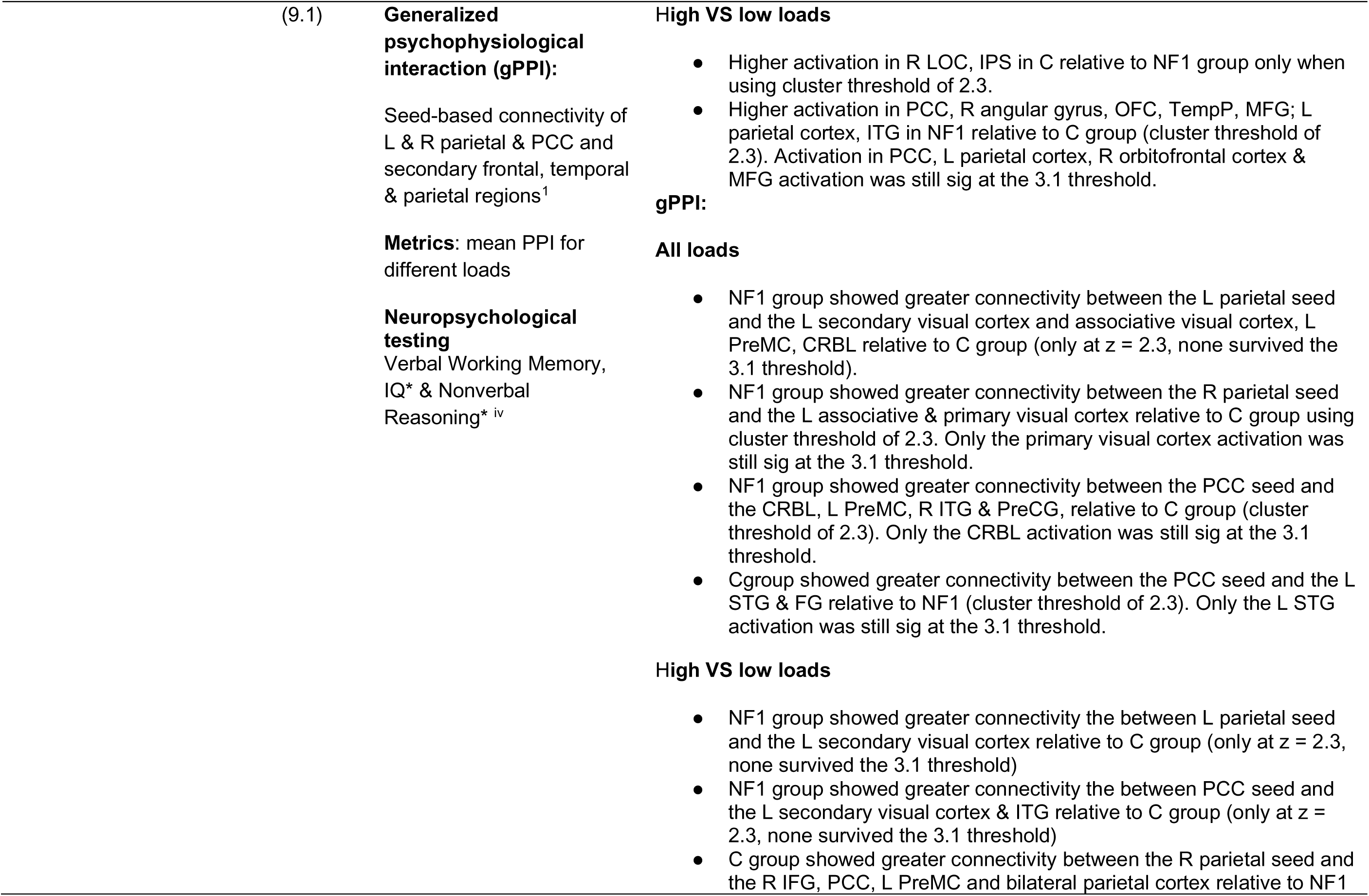
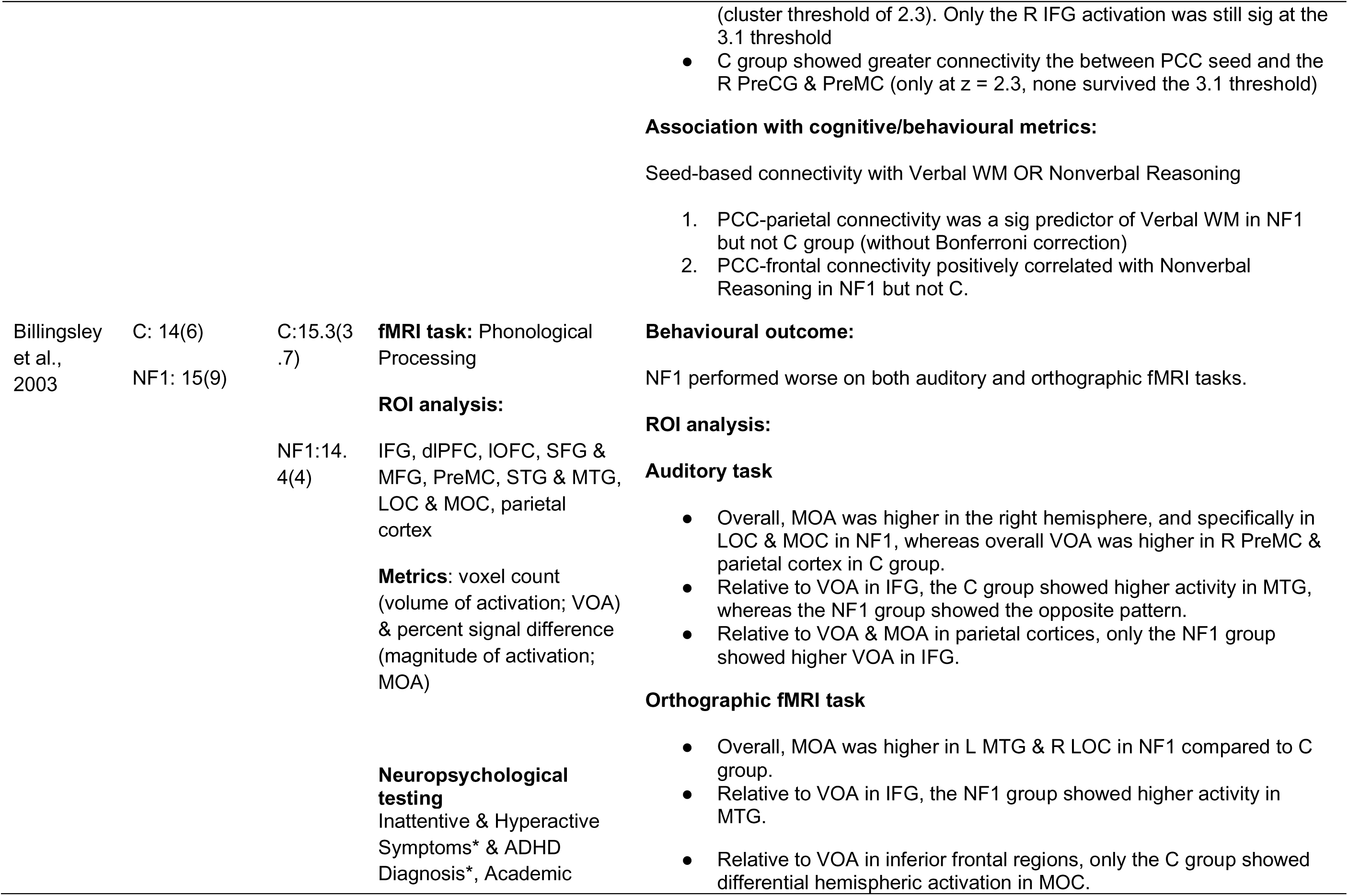
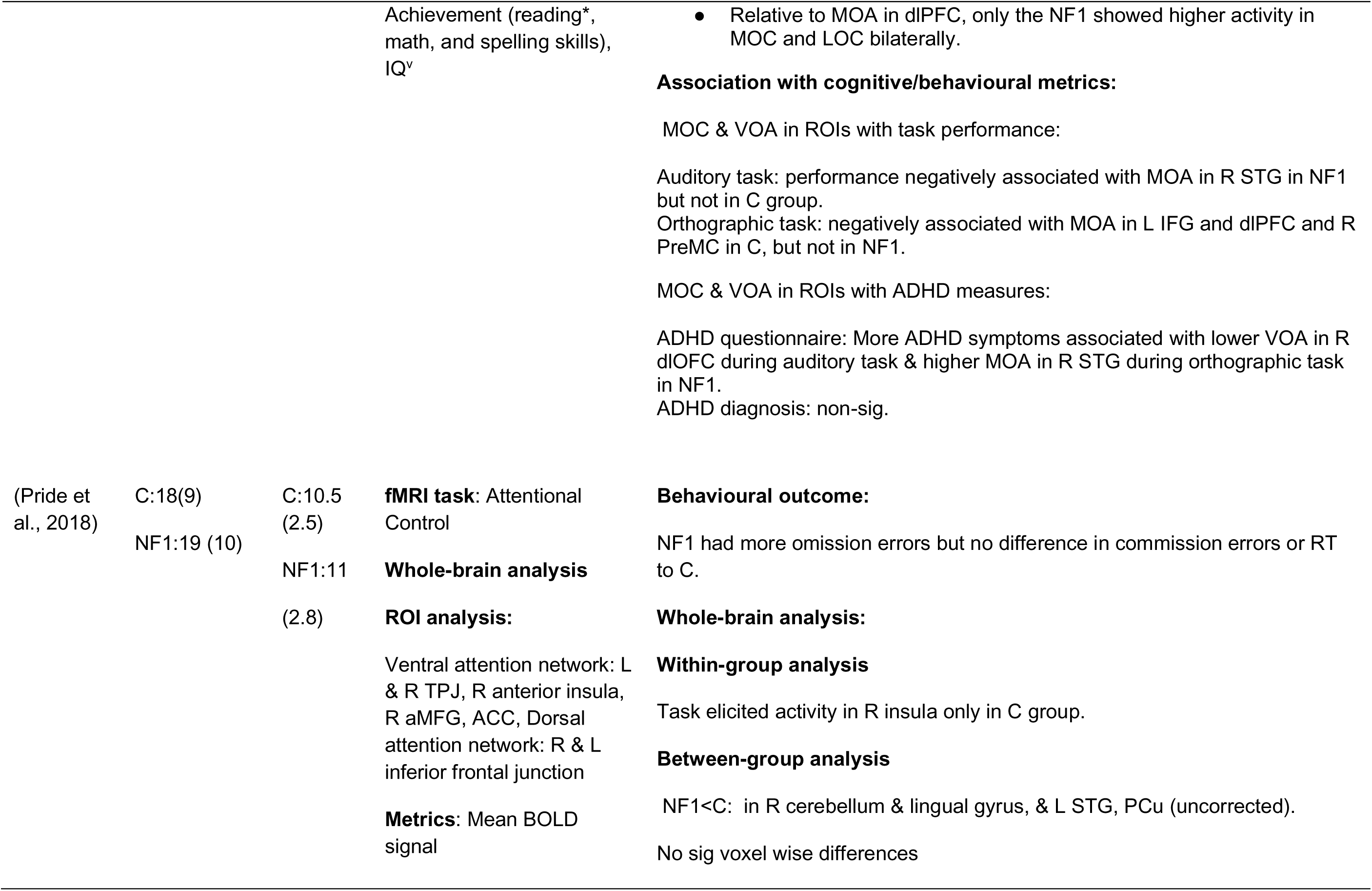
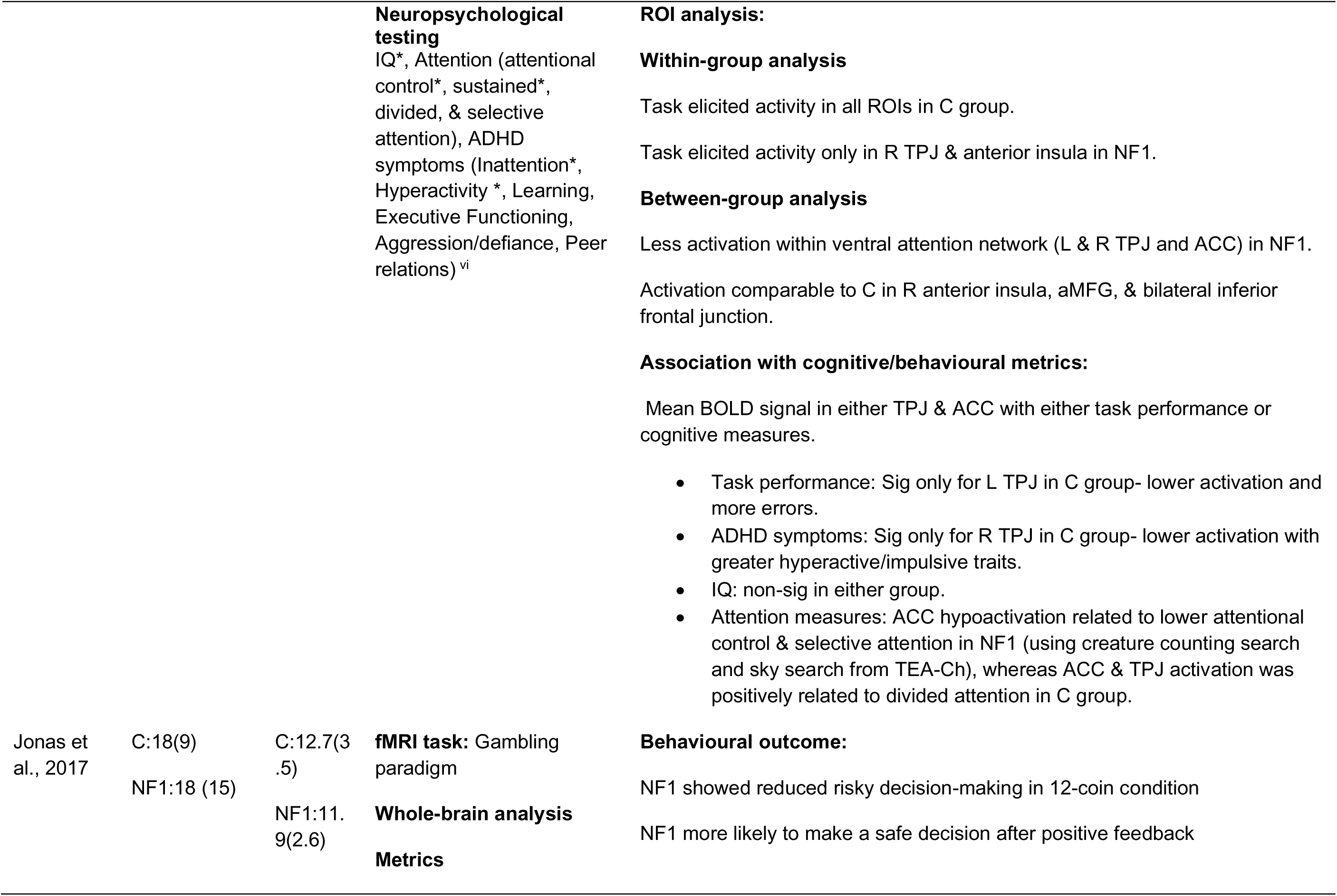
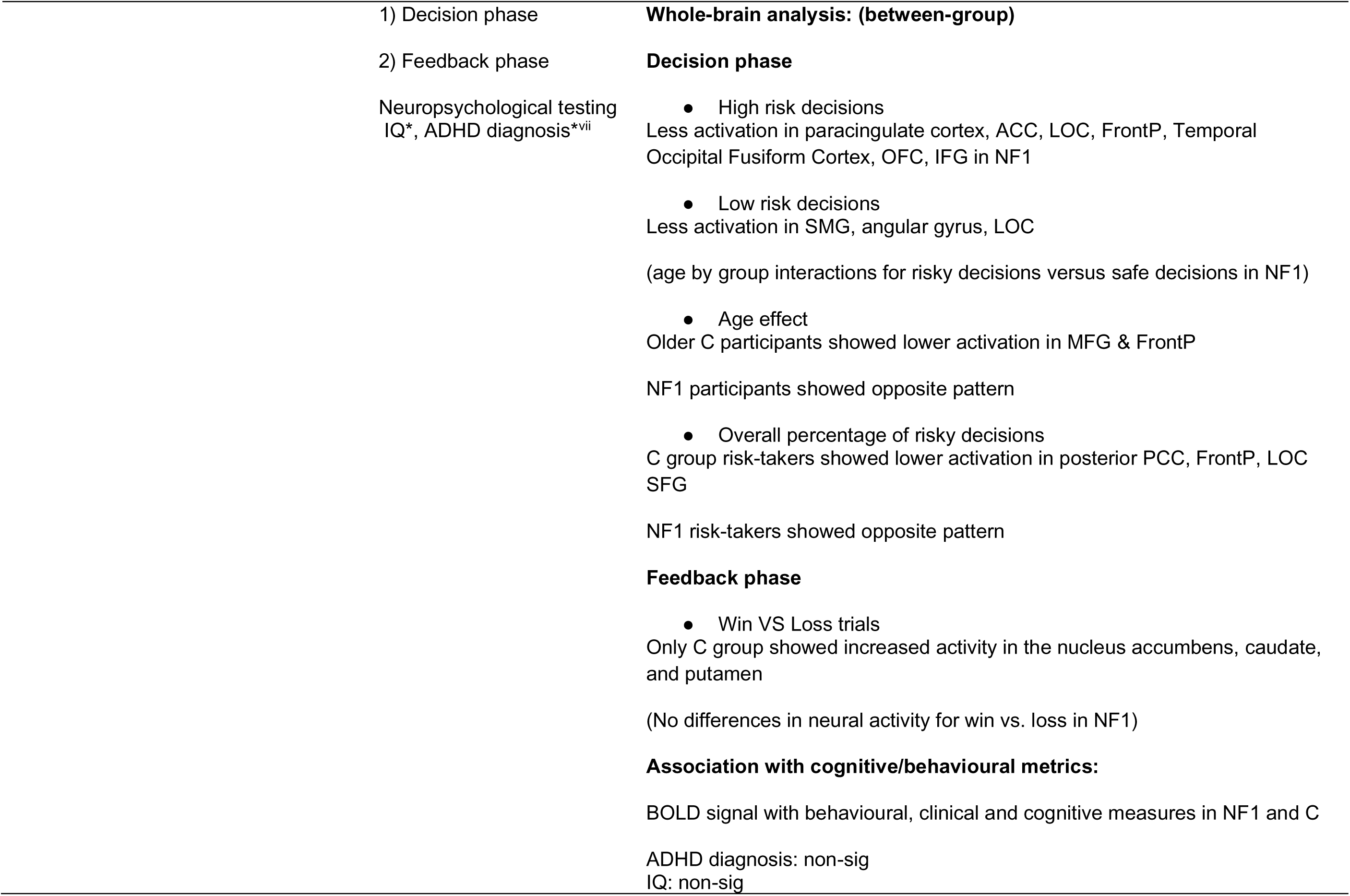
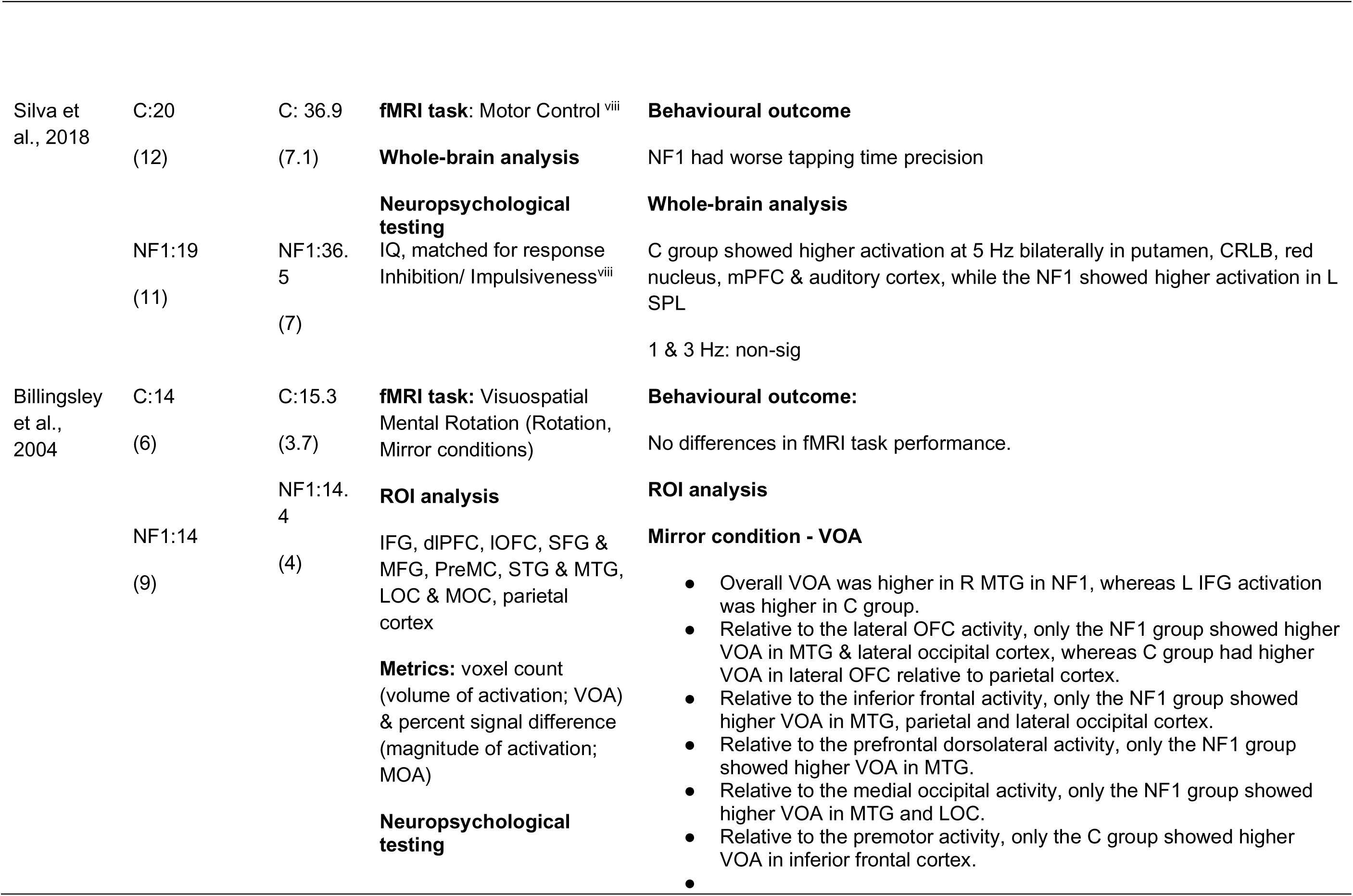
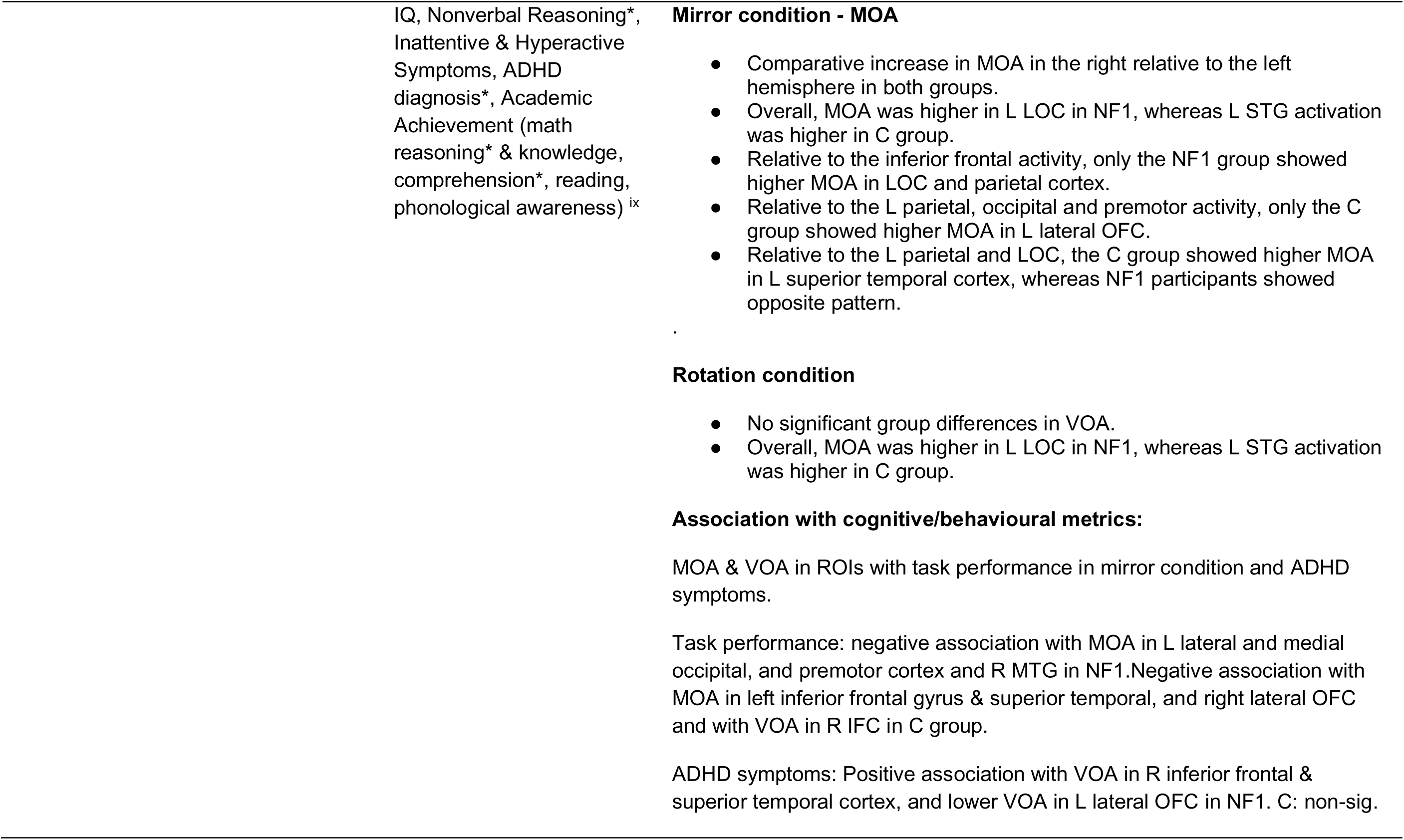
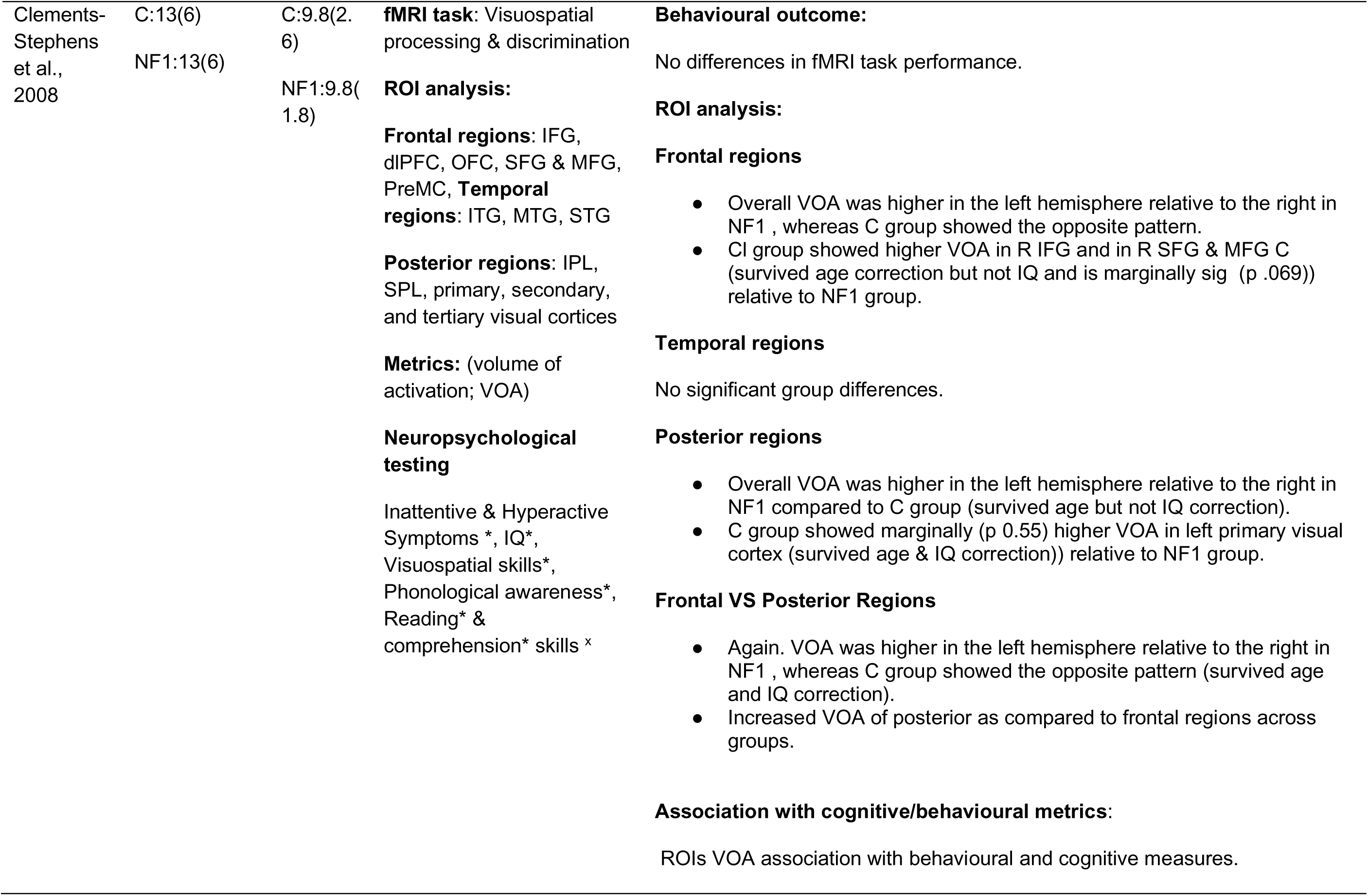
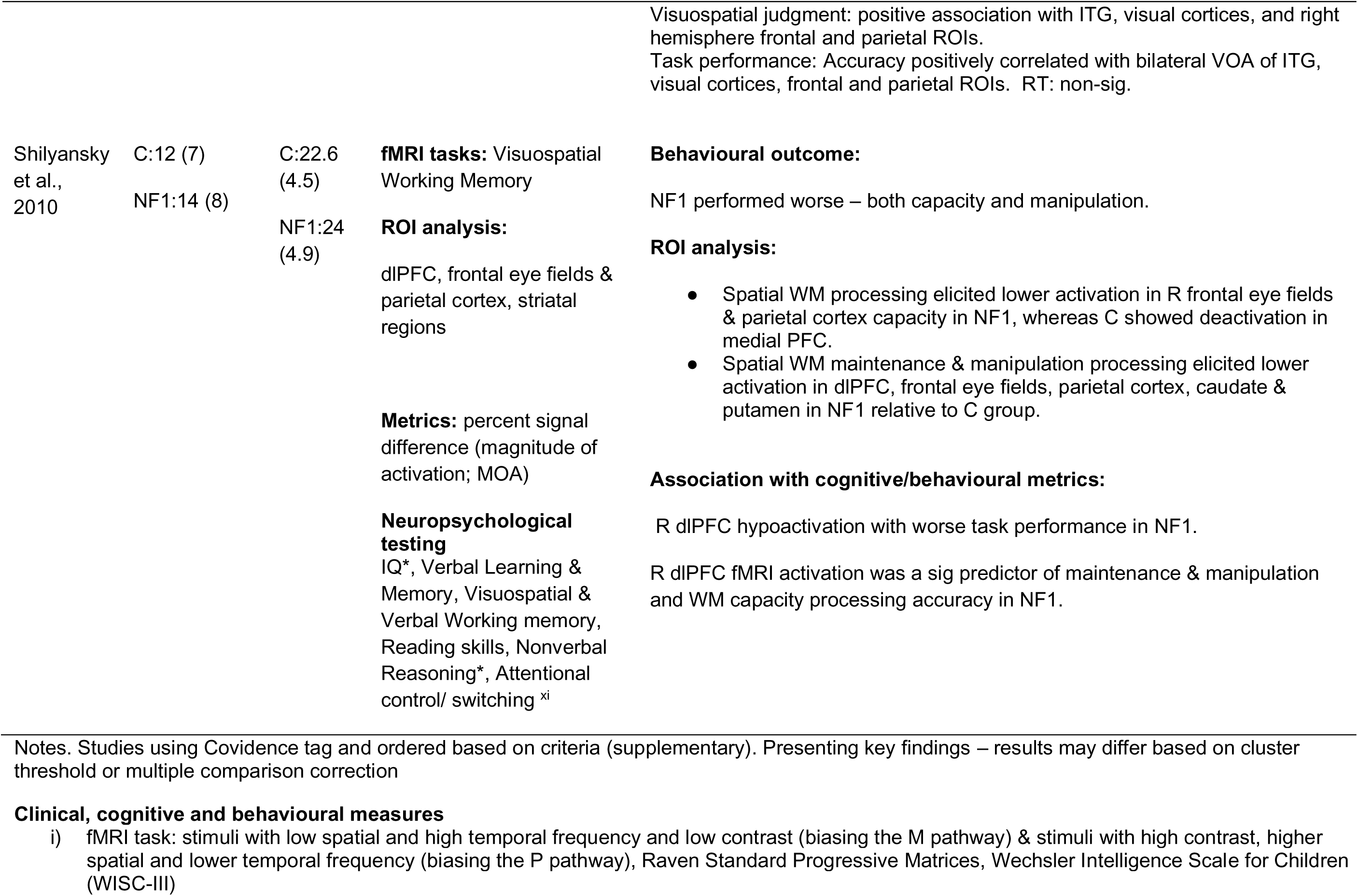
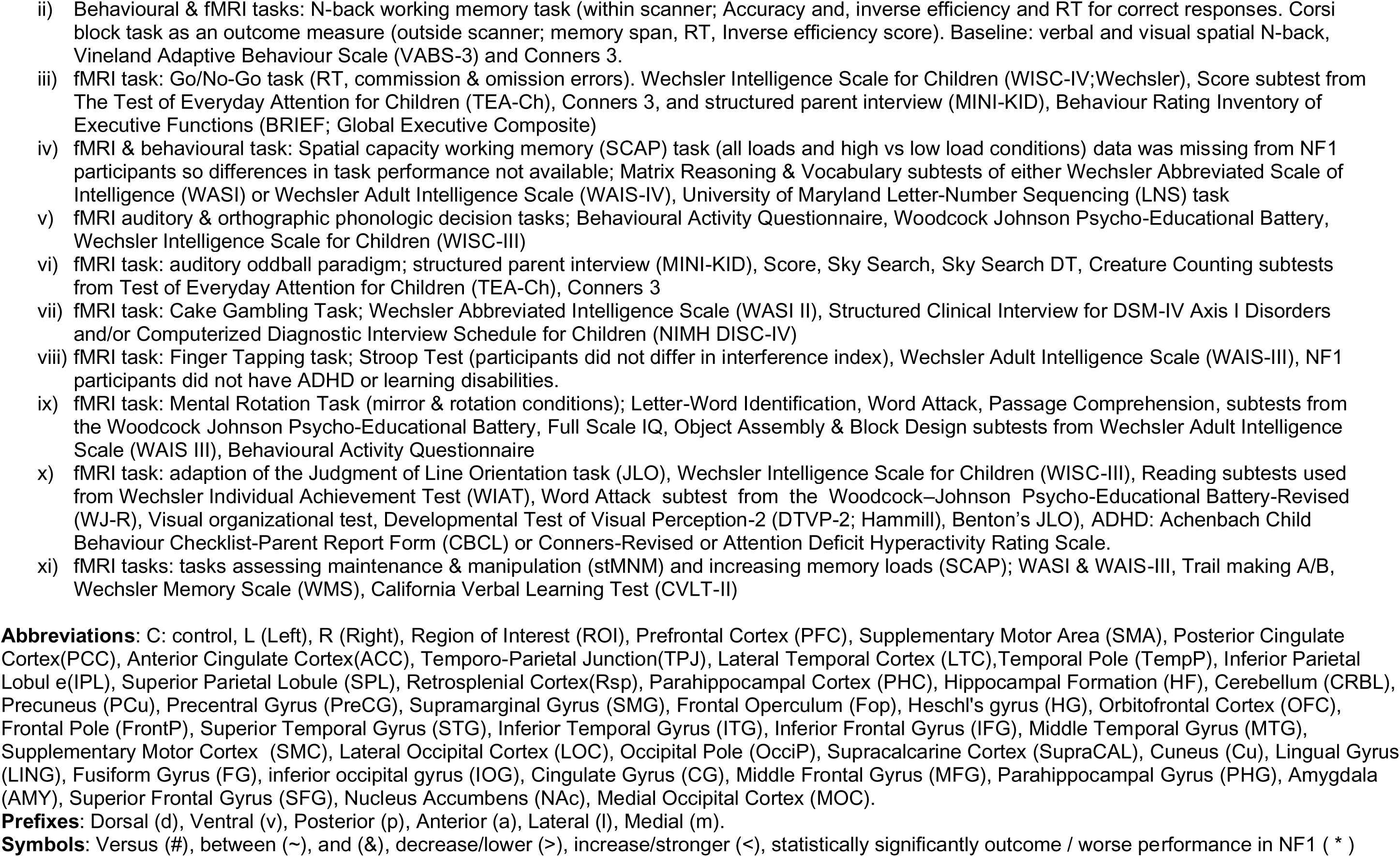
Task-based fMRI studies.

### 3.2 Studies using TMS

**Definition Box 2.** TMS terms used in the text.

|  |  |
| --- | --- |
| Paired pulse transcranial magnetic stimulation (TMS) | Two magnetic pulses are delivered in quick succession (e.g. conditioning pulse and then a test pulse), usually to the primary motor cortex |
| Intermittent theta burst stimulation (iTBS) | High frequency stimulation intended to induce cortical plasticity. |
| Motor evoked potential (MEP) | A magnetic or electrical pulse is applied over the motor cortex, generating a neural impulse that travels through the corticospinal tract to activate the target muscle, resulting in measurable activity. |
| Short interval intracortical inhibition (SICI) | SICI is a TMS-based measure of fast-acting cortical inhibition mediated by GABA<sub>A</sub> receptors. It is assessed using a paired-pulse protocol with a short interval (1-5 ms) between a subthreshold conditioning pulse and a suprathreshold test pulse. SICI reflects the suppression of motor cortical excitability and is associated with transient inhibitory processes. |
| Long interval intracortical inhibition (LICI) | LICI measures sustained cortical inhibition mediated by GABA<sub>B</sub> receptors. It uses a paired-pulse TMS protocol with a longer interval (50-200 ms) between two suprathreshold pulses. The inhibition observed in LICI reflects prolonged suppression of motor cortical excitability and slower-acting inhibitory mechanisms. |
| Intra cortical facilitation (ICF) | ICF serves as a valuable tool for studying cortical excitability and plasticity. Assessed using paired-pulse TMS, when the interval between the subthreshold pulse and the suprathreshold test pulse is 8-30ms, the MEP elicited by the test pulse is |
|  | enhanced. This is thought to reflect excitatory processes in the motor cortex mediated by NMDA receptors. |
| Resting motor threshold (RMT) | Minimum TMS intensity needed to produce a MEP. |
| Cortical silent period (CSP) | After a MEP there is a temporary interruption of muscle activity, known as the CSP, which is a measure of intracortical inhibition. |

(Castricum et al., 2020; Di Lazzaro & Ziemann, 2013; Du et al., 2014; Ziemann et al., 2015)

Four studies in adult NF1 samples using TMS-based measures (short and long interval intracortical inhibition-SICI, late cortical disinhibition or cortical silent period-CSP) of baseline cortical GABAergic inhibition did not find evidence of group differences (Castricum et al., 2020; Germanidis et al., 2021; Lacroix et al., 2022; Zimerman et al., 2015). Only one study by Mainberger et al. (2013) found increased SICI in NF1 compared to controls suggesting increased intracortical inhibition. In addition, Mainberger et al. (2013) carried out a double-blind placebo-controlled trial using 200mg of lovastatin in NF1, which is an inhibitor of 3-hydroxy-3-methylglutaryl coenzyme A (HMG-CoA) and has been identified as an inhibitor of the RAS/MAPK pathway. The drug normalised SICI, such that after a 4-day course, SICI was comparable between patients and controls. The Lovastatin treated NF1 group showed increases in attentional performance as compared to the placebo group, suggesting that normalisation in intracortical inhibition improves focus.

Two studies examined the modulation of intracortical inhibition during motor skills acquisition using a procedural learning task in NF1 patients without underlying cognitive impairments (Germanidis et al., 2021; Zimerman et al., 2015). Zimerman et al. (2015) found NF1 patients with no cognitive impairments showed reduced motor skill acquisition over five consecutive training days compared to controls, predominantly driven by reduced offline effects (between training days) but also reduced fast-online learning (early acquisition phase). The control group showed increased task-specific modulation of SICI, such that there was a reduction in inhibition, and this was associated with better motor learning. This effect was reduced in the NF1 group and associated with worse learning performance. Authors suggest impaired acquisition of skills may be related to alteration of intracortical GABAergic neurotransmission in line with the animal findings in NF1. In contrast, Germanidis et al. (2021) found no group differences in task specific modulation of intracortical inhibition between the NF1 and control groups over a three-day training period. Specifically, the apparent conflicting findings reported in the two studies focusing on procedural learning could be explained by key methodological differences i.e. the differences in training period length, the task, and the study sample size.

Excitability and plasticity were investigated by two studies that used intermittent theta burst stimulation (iTBS) and paired associative stimulation (PAS) protocols (Castricum et al., 2020; Mainberger et al., 2013). Both studies found reduced plasticity in NF1; there was no evident increase in MEP amplitude following PAS-induced plasticity as there was in controls (Mainberger et al., 2013) and overall MEPs were significantly lower and shorter-lasting in the NF1 group compared to controls following iTBS (Castricum et al., 2020). Furthermore, lovastatin normalised this deficit in plasticity, with increases in MEP amplitude observed in NF1 in the treatment group, but not the placebo group, following PAS (Mainberger et al., 2013). Despite Zimerman et al. (2015) findings of a lack of task specific reduction in inhibition associated with worse task performance, other studies investigating intracortical inhibition in the primary motor cortex did not reveal significant associations of TMS-based indices with IQ or behavioural learning (Castricum et al., 2020; Germanidis et al., 2021).

Finally, Doherty et al. (2023) investigated the balance between excitation and inhibition using SICI and ICF in children with NF1, controls and an ADHD group. ICF was lower in NF1 than both controls and ADHD, with reduced excitation consistent with previous findings of reduced plasticity (Castricum et al., 2020; Mainberger et al., 2013), although SICI did not differ from controls or ADHD. However, in NF1, less ICF was also associated with more inhibition (through lower SICI ratios), reinforcing a reduced excitation relative to inhibition ratio in NF1. Moreover, the Physical and Neurological Examination for Soft Signs (PANESS) scale for motor development, which found worse scores in NF1 than both controls and ADHD, was investigated alongside TMS recordings. Interestingly, better scores associated with more inhibition and less excitation in NF1, which unlike Mainberger et al. (2013), suggests that a reduced E/I balance in NF1 may be compensatory rather than maladaptive.

**Table 4.**
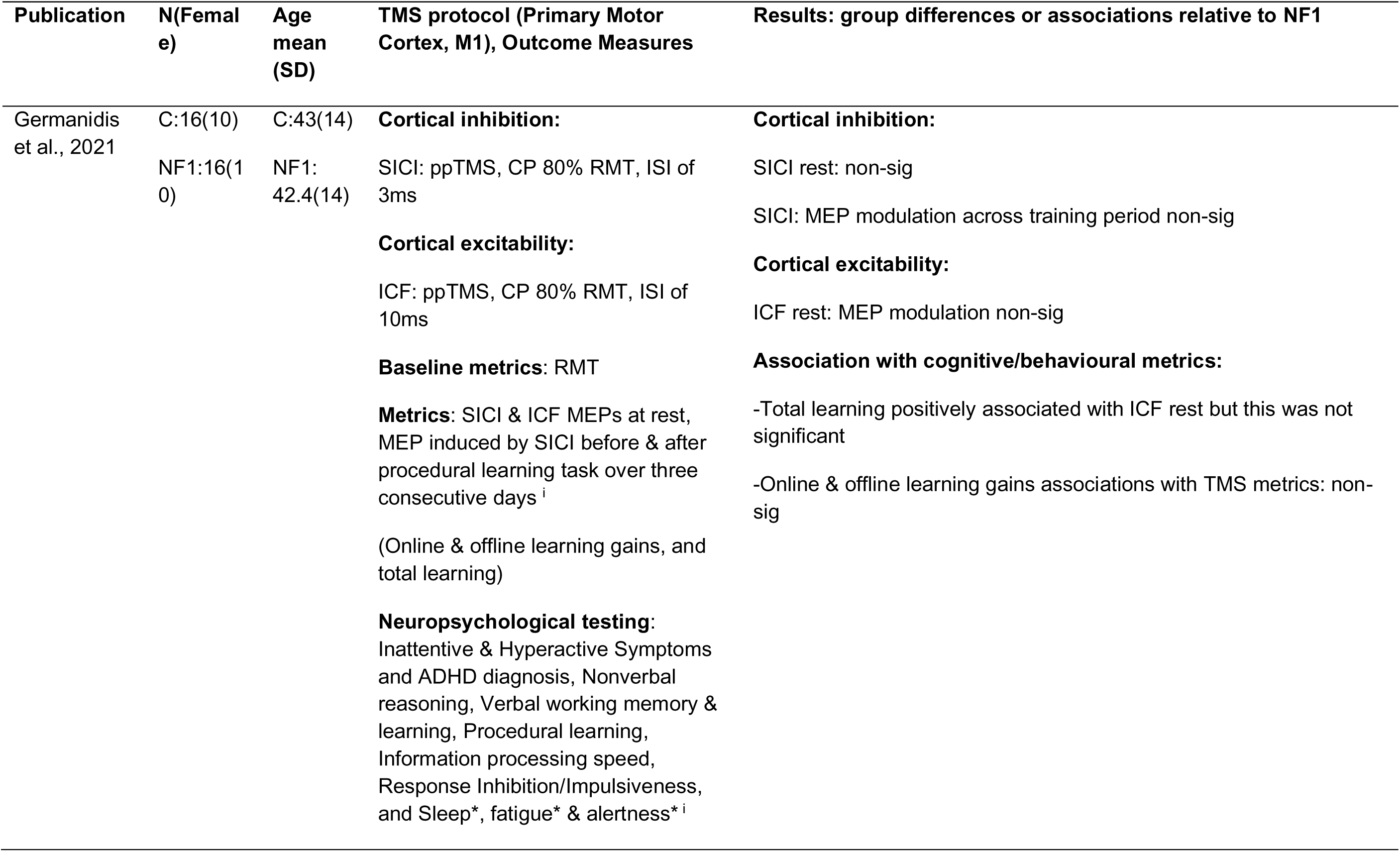
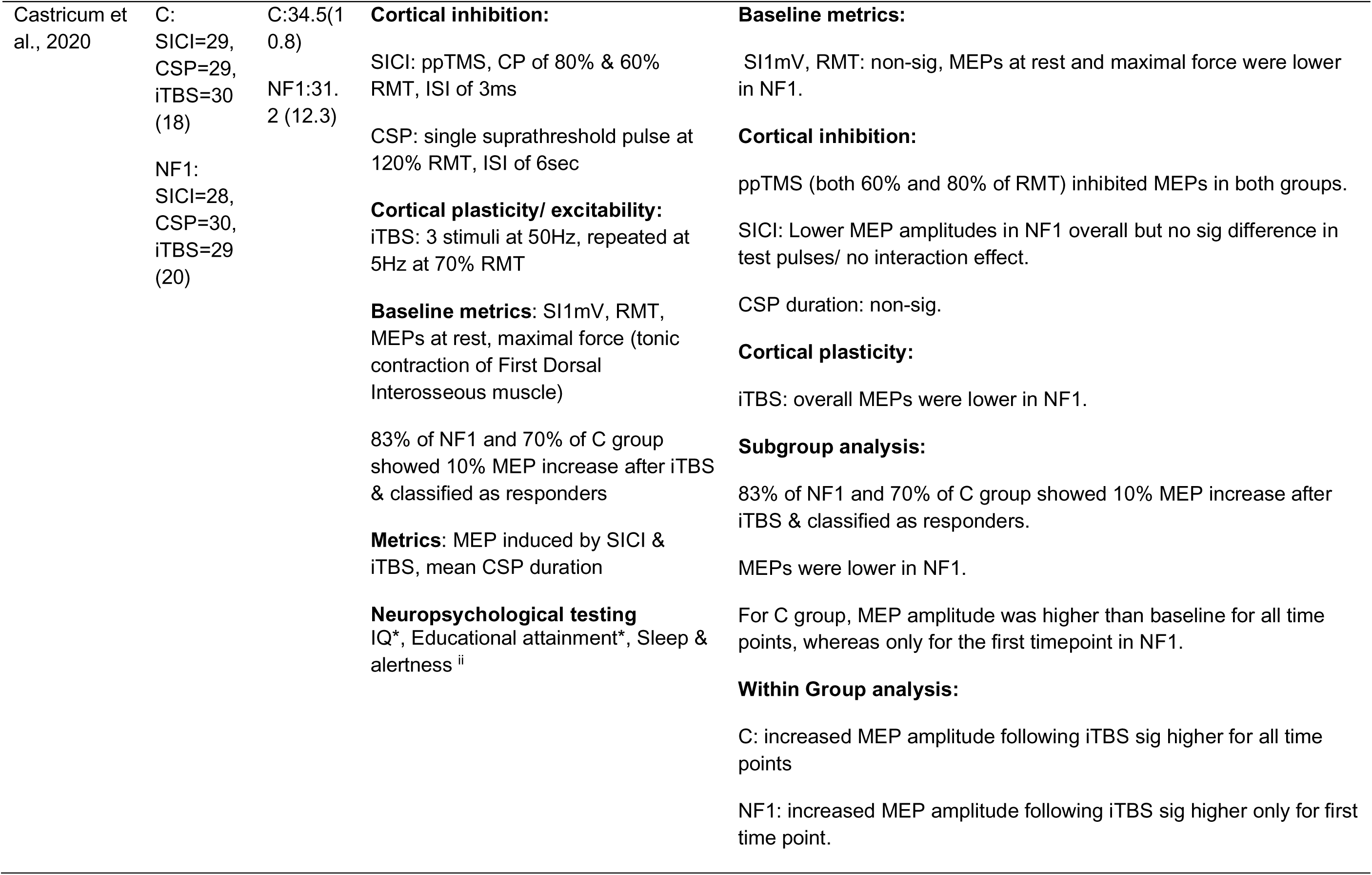
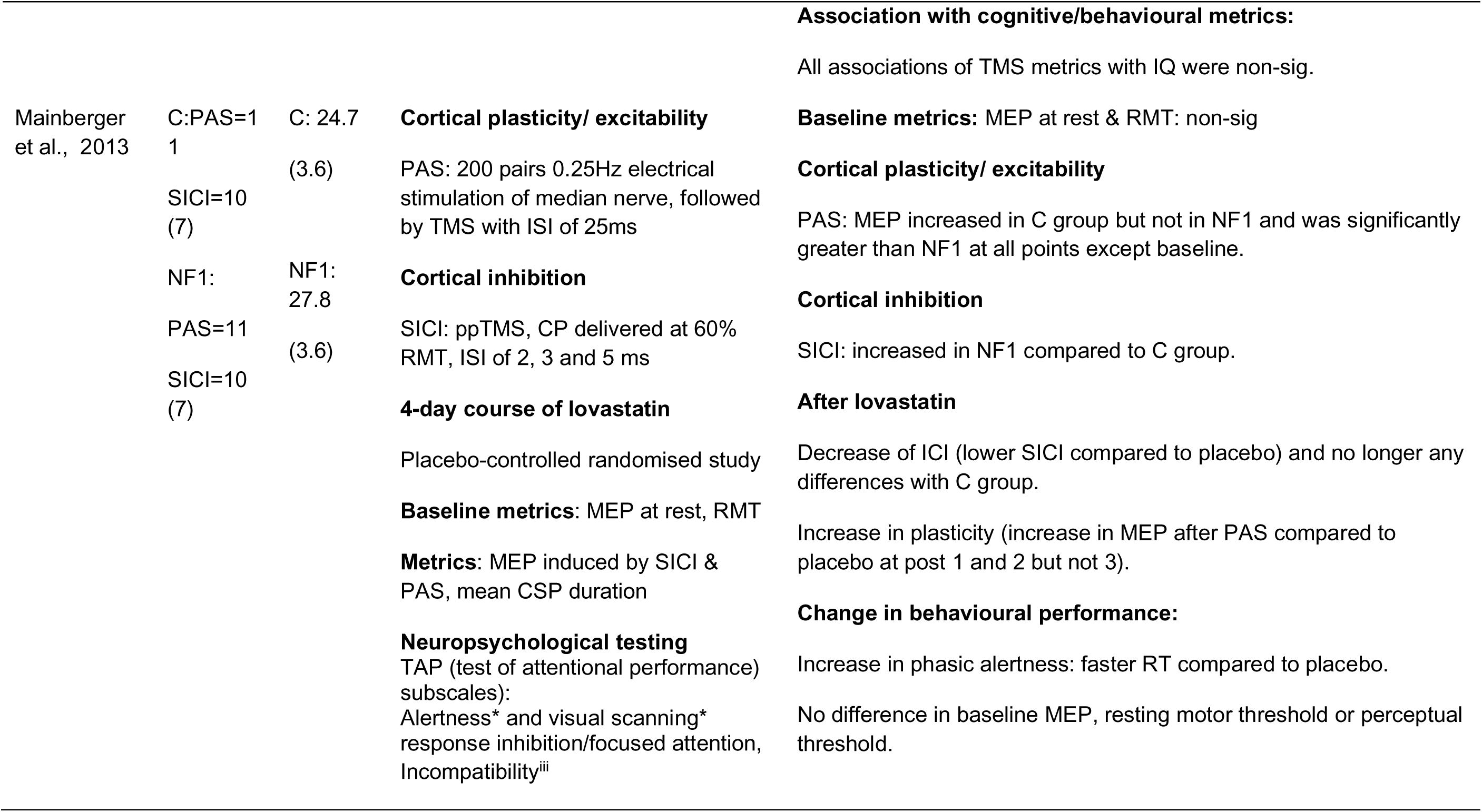
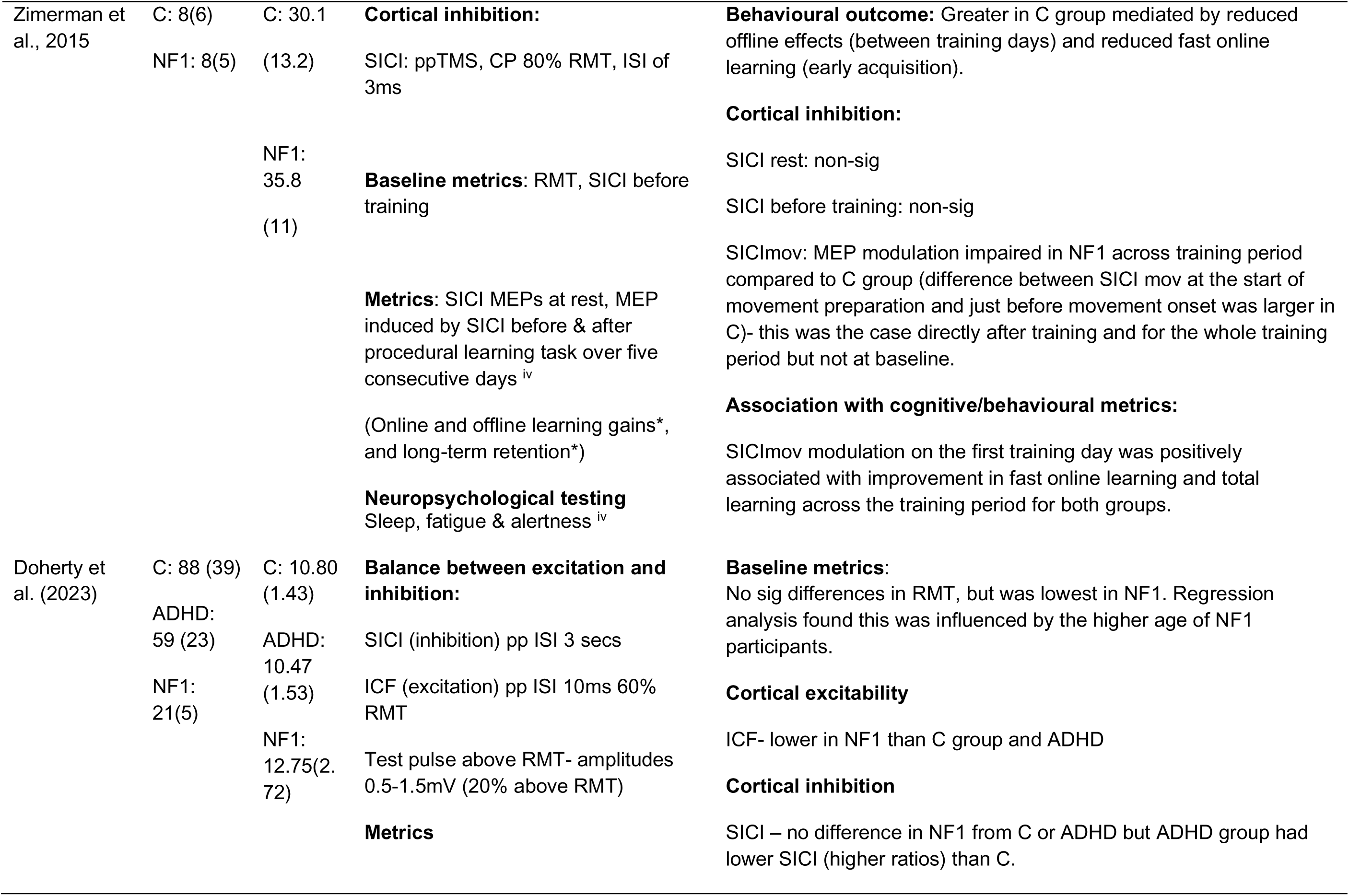
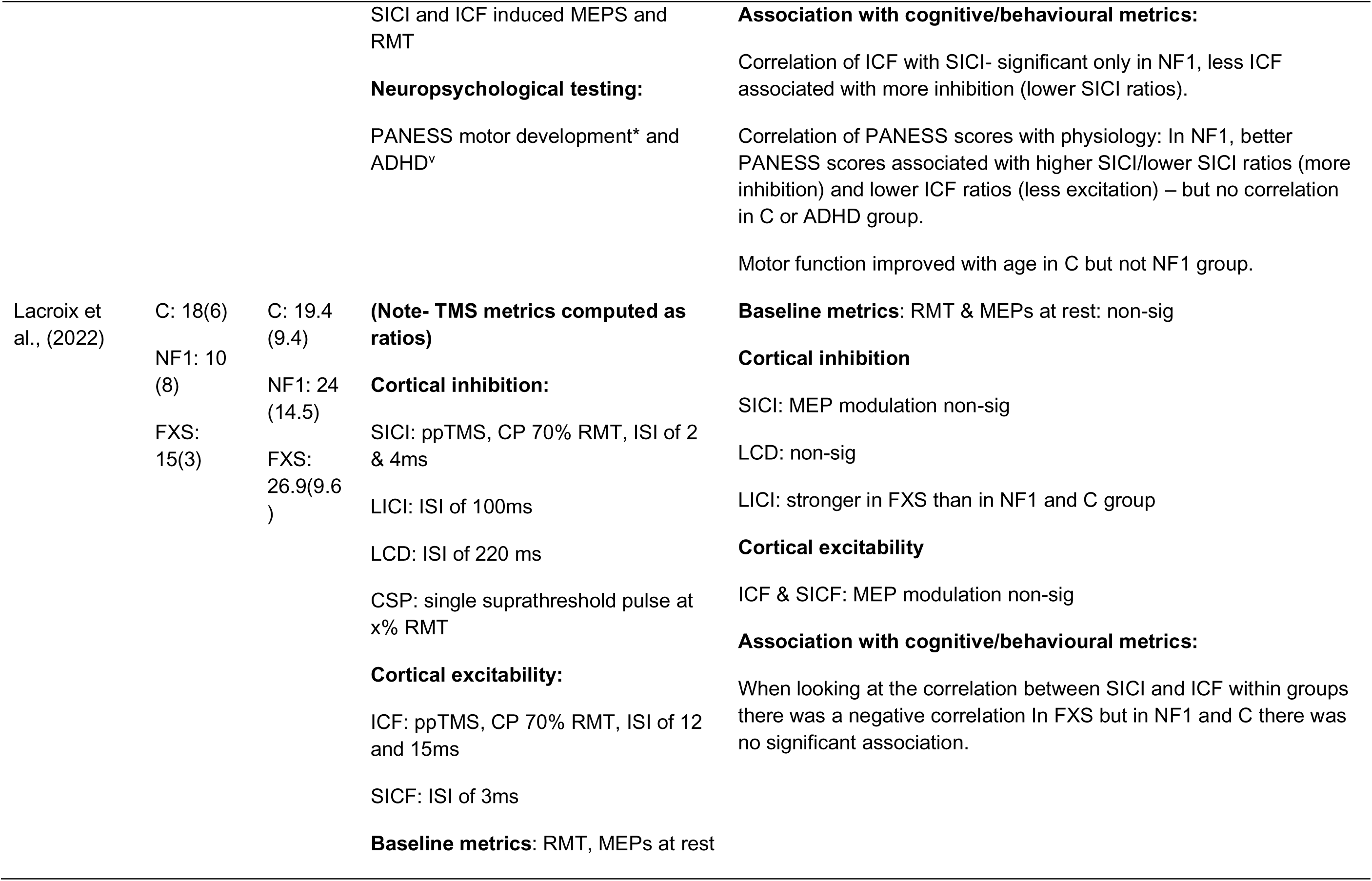
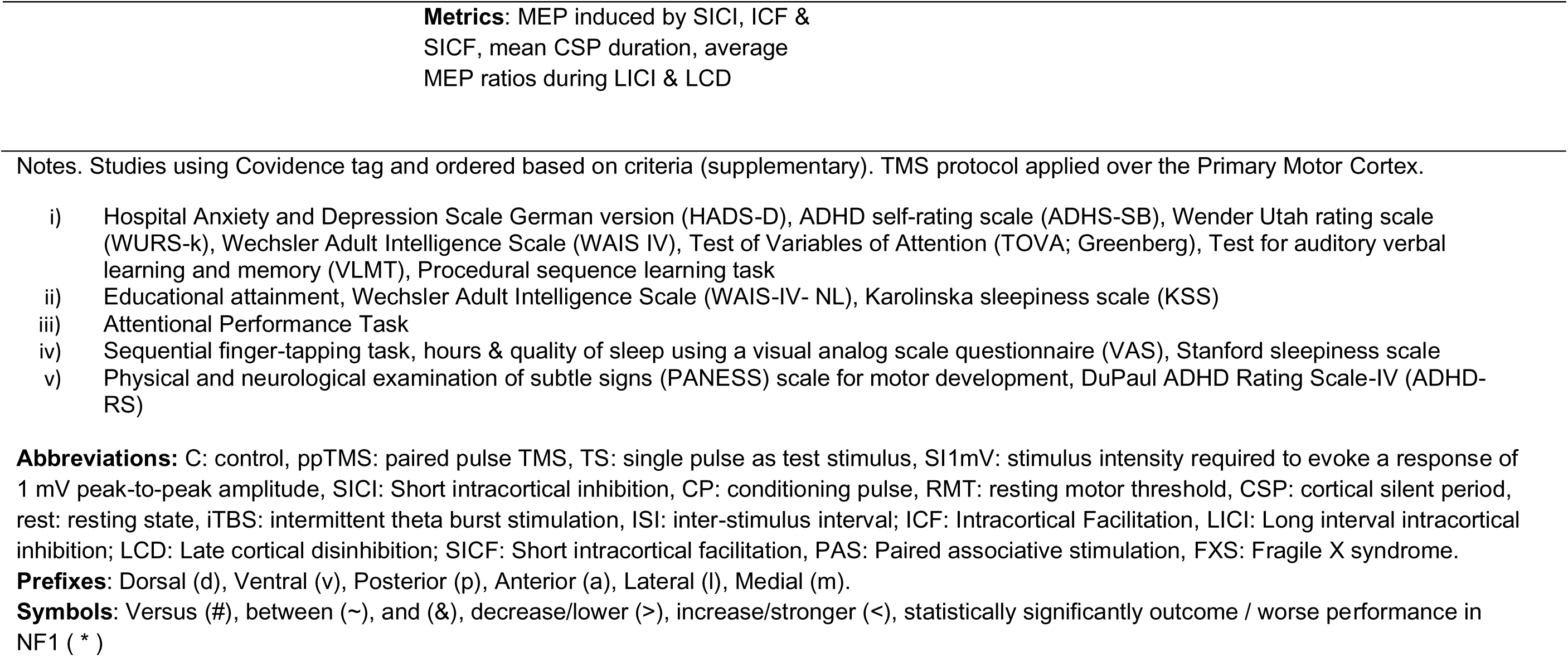
TMS studies.

### 3.3 Studies using EEG

**Definition Box 3.** EEG terms used in the text.

|  |  |
| --- | --- |
| Time domain analysis (Event related potentials (ERPs) and evoked potentials) | Waveform phase locked to stimulus onset containing components associated with a different aspect of cognitive processing, of which the amplitude and latency can be calculated. `Evoked potentials are similarly time locked to a stimulus and refer to response to a specific external sensory stimulus e.g. VEP, rather than higher order cognitive processing that ERPs refer to.<br>P1: 80-130 ms elicited by visual stimuli, most prominent over occipital electrodes.<br>P2: 50-250 ms associated with attention and stimulus evaluation, prominent over central and frontocentral areas.<br>P3: 300-600 ms elicited by infrequent or task-relevant stimuli, prominent over parietal and central electrodes.<br>N1: 80-150 ms elicited by auditory or visual stimuli, most prominent over frontal and central electrodes.<br>N2: 200-350 ms linked to conflict monitoring or mismatch detection, prominent over frontocentral regions.<br>N450: 350-500 ms associated with conflict resolution, most prominent over frontocentral electrodes. |
| Steady state visual evoked potential (SSVEP) | Refers to a type of brain response that is evoked by repetitive visual stimuli, presented at a constant, repetitive frequency. These potentials are recorded from the brain using EEG |
|  | and are analysed in the frequency domain. |
| Alpha waves | 8-12Hz, most prominent when awake but at rest/relaxed often in occipital and parietal regions. Alpha desynchronisation/ reflects active neural processing, whereas higher alpha power occurs when at rest, processing distractors or task irrelevant regions. |
| Theta waves | 2-8Hz, commonly in frontal cortex or hippocampus and is associated with learning and memory. |
| Beta waves | 10-30Hz, most prominent in motor cortex but also in prefrontal regions, higher beta power is linked to active processing and focused attention, may increase when ignoring irrelevant stimuli. A decrease in beta power during movement and increase after movement (inhibition of further movement) is often observed. |

(Foxy & Snyder, 2011; Helfrich & Knight, 2019; Ippolito et al., 2022; Jensen & Mazaheri, 2010; Morales & Bowers, 2022; Zavala et al., 2018)

#### Cognitive control

Ribeiro et al. (2015) used a go/no go task in adolescents with NF1 and age and gender matched controls. Significant group VEP differences observed were lower P1 but not N1 amplitude in the NF1 group suggesting changes in basic visual processing or deficient allocation of spatial attention. The frontal P3 amplitude was reduced in the NF1 group and a negative correlation was found between P3 amplitude and GABA, possibly suggesting abnormal GABA signalling. Likewise, Bluschke et al. (2017a) used a go/no go task, and found reduced P1 amplitude in NF1 but no differences in P3 amplitude. The difference in the findings between the Bluschke et al. (2017a) and Ribeiro et al (2015) study was possibly related to the differences in the cognitive tasks. Furthermore, a different study by Bluschke et al. (2017b) examined the N2 and N450 neural responses to a modified Flanker task in children with NF1 as compared to controls. The N2 amplitude was selectively reduced for the trials in which flanker arrows pointed in the opposite direction in the NF1 group whereas no differences between compatible/incompatible trials were found in the control group. In contrast, in the N450 component, the control group showed a significant main effect of same/opposite direction arrows but not the NF1 group, suggesting a constantly enhanced N450 is used by NF1 group to achieve successful attentional control. The authors suggest that the constantly enhanced N450 leads to general slowing down of the response and can be attributed to dopamine deficiency in NF1.

#### Working Memory

Two studies used EEG to investigate working memory. Booth et al. (2023) investigated oscillatory activity during resting-state and a working memory n-back task in adolescents with NF1. Resting-state delta and theta power was increased whereas peak alpha frequency was decreased in NF1 as compared to controls. An association with age was evident in controls, such that resting delta and theta power decreased with age and peak alpha frequency increased with age, but this relationship was not noted in NF1. During the EEG working memory task, the NF1 group demonstrated higher theta power and theta phase coherence but this was no longer the case when controlling for resting/baseline EEG. Pobric et al. (2022) also used an n-back working memory paradigm, but to investigate differences in P3 amplitude and latency in adolescents with NF1 versus age and sex-matched controls. There were no group differences found in P3 amplitude but P3 latency was shorter in the NF1 group, suggesting differences in neural processing. The topography analyses of P300 demonstrated found imbalance in the distribution of neural activity across the frontal and parietal cortex in the NF1 group, with a weaker left frontal positivity and a stronger left parietal positivity as compared to controls.

#### Visual Processing

Evoked responses to a visual processing task were examined by Ribeiro et al. (2014) in adolescents with NF1 compared to controls. Visual stimulation was chosen to bias activation to three detection mechanisms including achromatic (eliciting steady state VEPS), red-green and blue-yellow (eliciting transient VEPs). Average VEPs across 6 parieto-occipital channels (mean total response amplitude for achromatic stimuli and P1 and P2 for chromatic stimuli) showed a significant effect of stimulus contrast but no group differences were observed. The late component of chromatic VEPs at 300ms showed a significant group difference, with a negative peak in controls, but not in NF1. Frequency-domain analyses revealed group differences elicited by achromatic stimulation in non-phase locked oscillations particularly with higher NF1 amplitude in the alpha band. Similarly, frequency-domain analyses of the EEG elicited by the chromatic visual stimulation showed significant group differences with greater NF1 alpha amplitude. Poorer behavioural performance in the visual detection task was noted in the NF1 group, and alpha amplitude was higher in both NF1 and controls during missed trials compared to correct trials. Furthermore, higher alpha and theta amplitude in NF1 were identified at rest. The authors attributed the difference in the alpha rhythm to difficulties with sustained attention in NF1 and suggest a thalamic dysfunction contributing to this phenotype.

Alpha band oscillations were studied by Silva et al. (2016) in adolescents with NF1 and controls using time-frequency analysis. Posterior alpha and beta oscillations were observed to undergo greater desynchronisation in the NF1 group using a task which required covert attention from participants to perceive changes in a peripherally presented stimulus. Furthermore, significant hemispheric differences in alpha rhythms were observed in the controls but not the NF1 group, such that alpha desynchronisation due to stimulus onset was greater in the contralateral hemisphere. Overall, the authors speculate that strong alpha suppression during visual presentation may suggest a compensatory regulation of exogenous attention. Similarly, Lalancette et al. (2022) also investigated visual processing by measuring steady-state visual evoked potentials (SSVEPs) in children with NF1 and controls (4-13 yrs) during visual stimulation at different frequencies. Signal to noise ratio (SNR) of SSVEPs was compared between NF1 and controls. There was a higher SNR in controls versus NF1 at 15Hz stimulation, although only in NF1 individuals who also had comorbid ADHD. Likewise, the signal strength at 10 and 15hz was negatively associated with inattention symptoms and 15Hz with severe behavioural and emotional problems, but only in the NF1 group. Therefore, the authors suggest that lower SNR of SSVEPs in NF1 may contribute to attentional difficulties.

Finally, Carter Leno et al. (2022) investigated the aperiodic slope component of the EEG power spectrum in infants with NF1 versus controls whilst watching videos designed to create a calm attention. The aperiodic exponent was higher in NF1, suggestive of greater inhibition relative to excitation

#### Auditory Processing

In a longitudinal study Begum-Ali et al. (2021) examined auditory repetition suppression and change detection in infants with NF1 at 5, 10 and 14 months of age. The authors found that the evoked responses decreased with age (5-14 months) in the posterior regions and became more frontally specific in the control group. Such age-related changes were reduced in the NF1 group. The auditory responses at 10 months did not relate to later language acquisition but were related to autistic traits measured at 14 months. Overall, the authors suggest slower neural detection of repetition or change and diminished topographic developmental changes in the NF1 group. Lalancette et al. (2023) reinforced the finding of differences in developmental changes in NF1, using a similar auditory paradigm in children between 4 and 13 years old, measuring alpha, beta and theta power with time frequency analysis. Repetition suppression resulted in comparable spectral power and phase synchronisation between groups, although neural responses to change detection did differ, with increased theta power in NF1. A negative association was observed across all participants between alpha power during change detection and inattention and hyperactivity, such that elevated symptoms occurred with reduced alpha power.

Fontanelli et al. (2023) investigated the effect of an auditory intervention on central auditory processing performance and auditory evoked potentials in NF1. This consisted of 9 weekly 50 minute sessions with auditory stimulation under progressively more difficult listening conditions focusing on figure-ground segregation and binaural integration. Sessions occurred both pre and post intervention as well as a 4 month follow up. The intervention group improved in measures of auditory processing between sessions and were greater than the no-intervention group in both follow up sessions, with comparable performance at baseline. In addition, components of the auditory evoked potential that were absent at baseline (P3 and N2) were evident following the intervention.

**Table 5.**
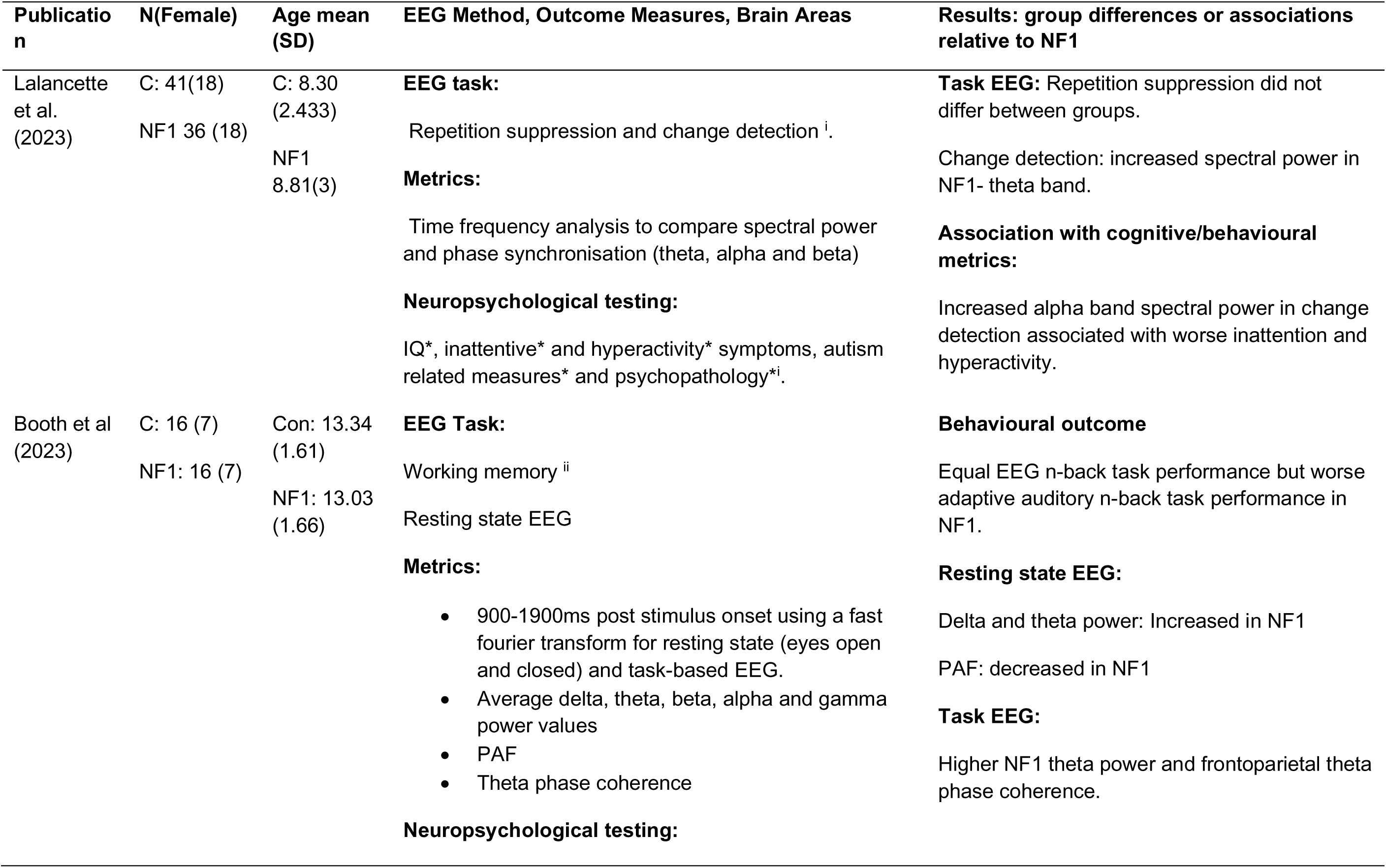
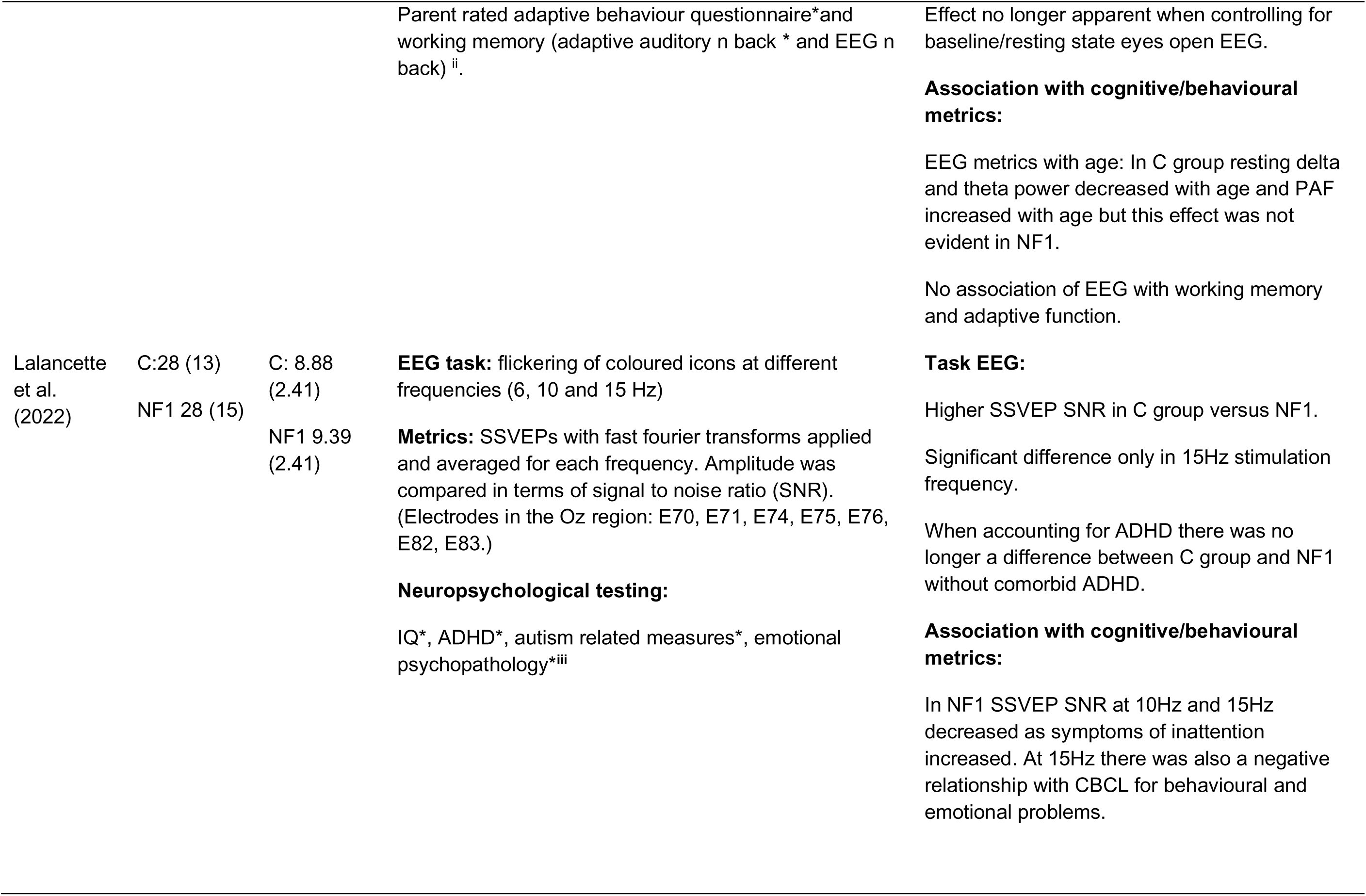
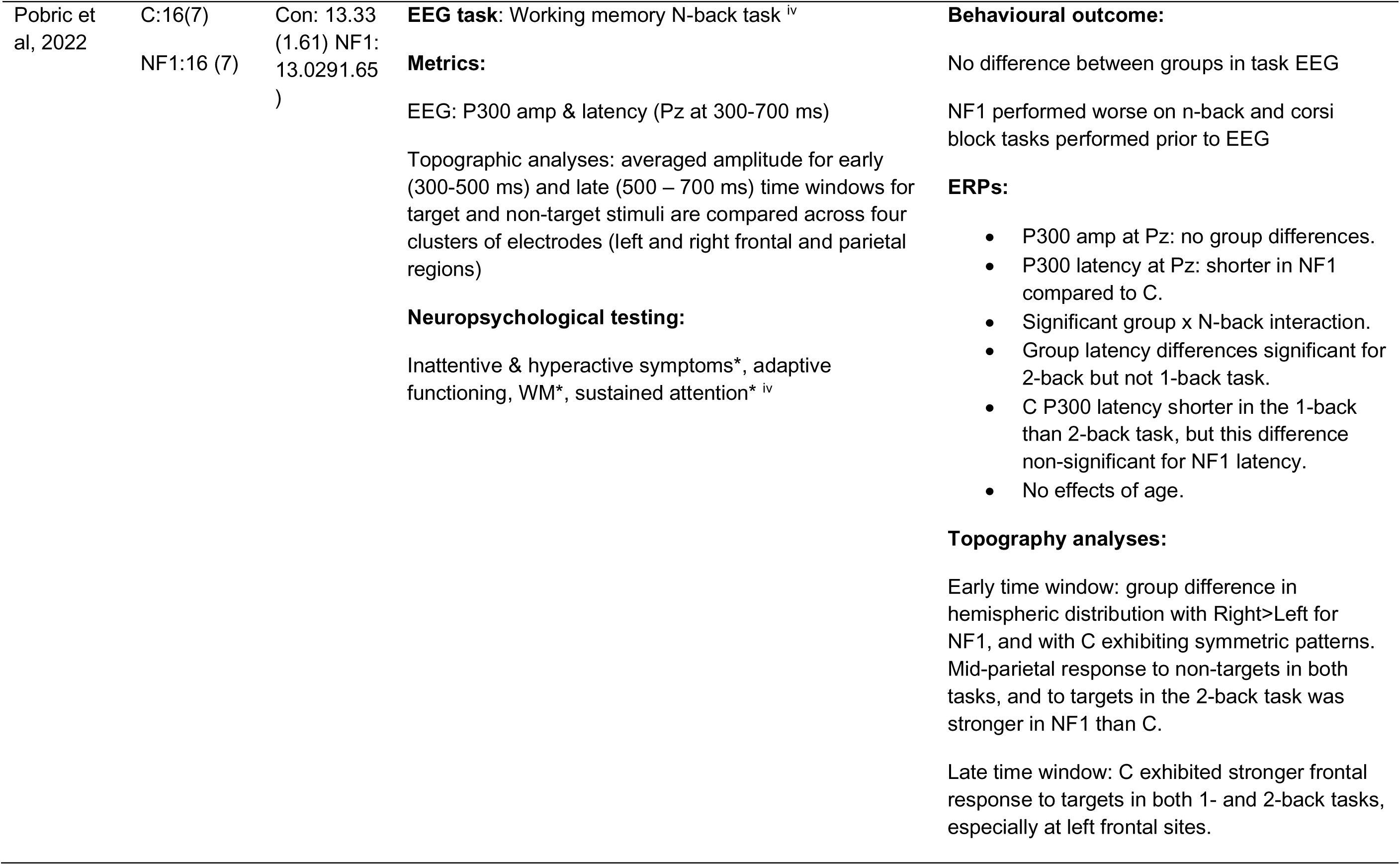
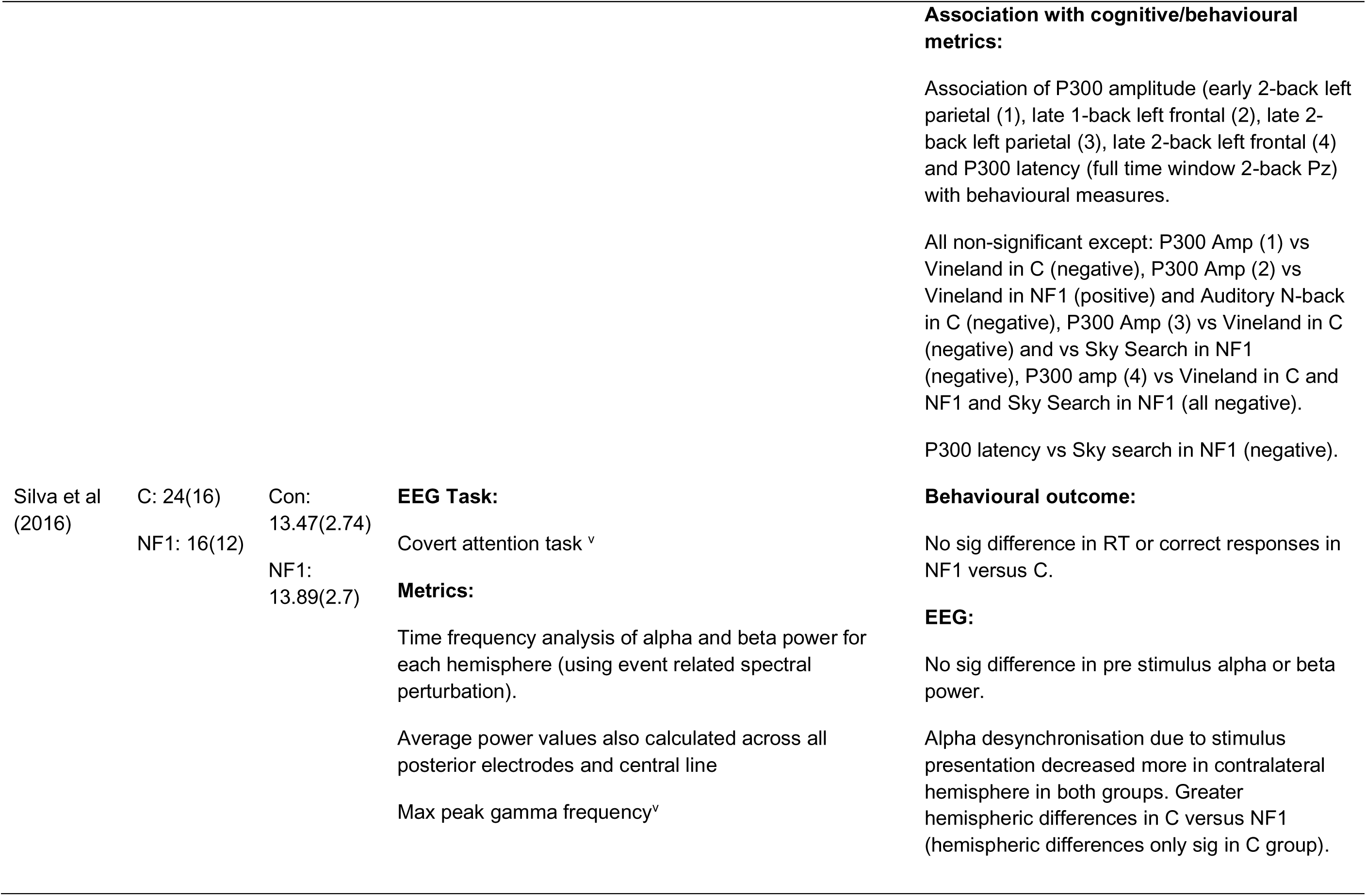
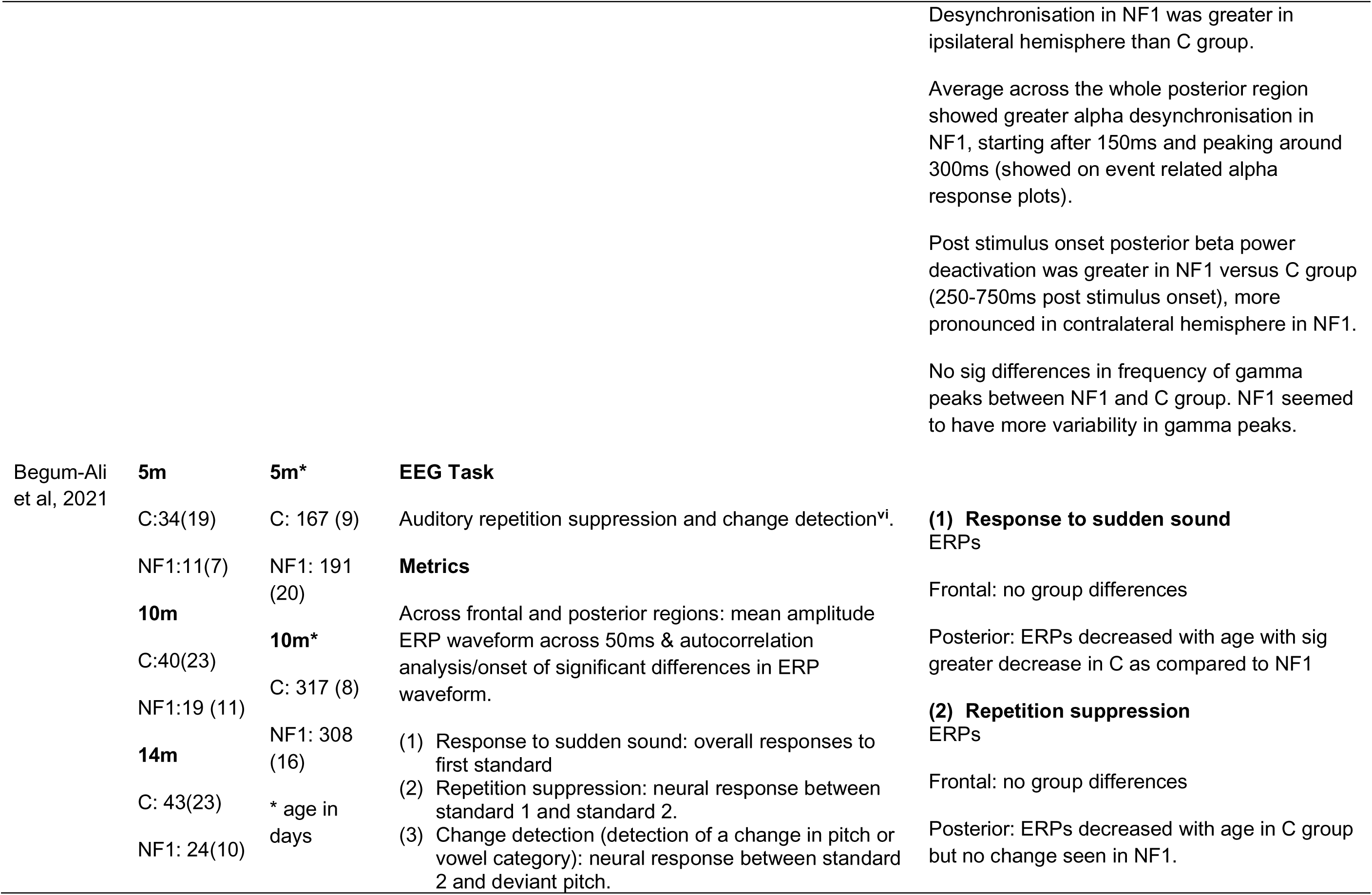
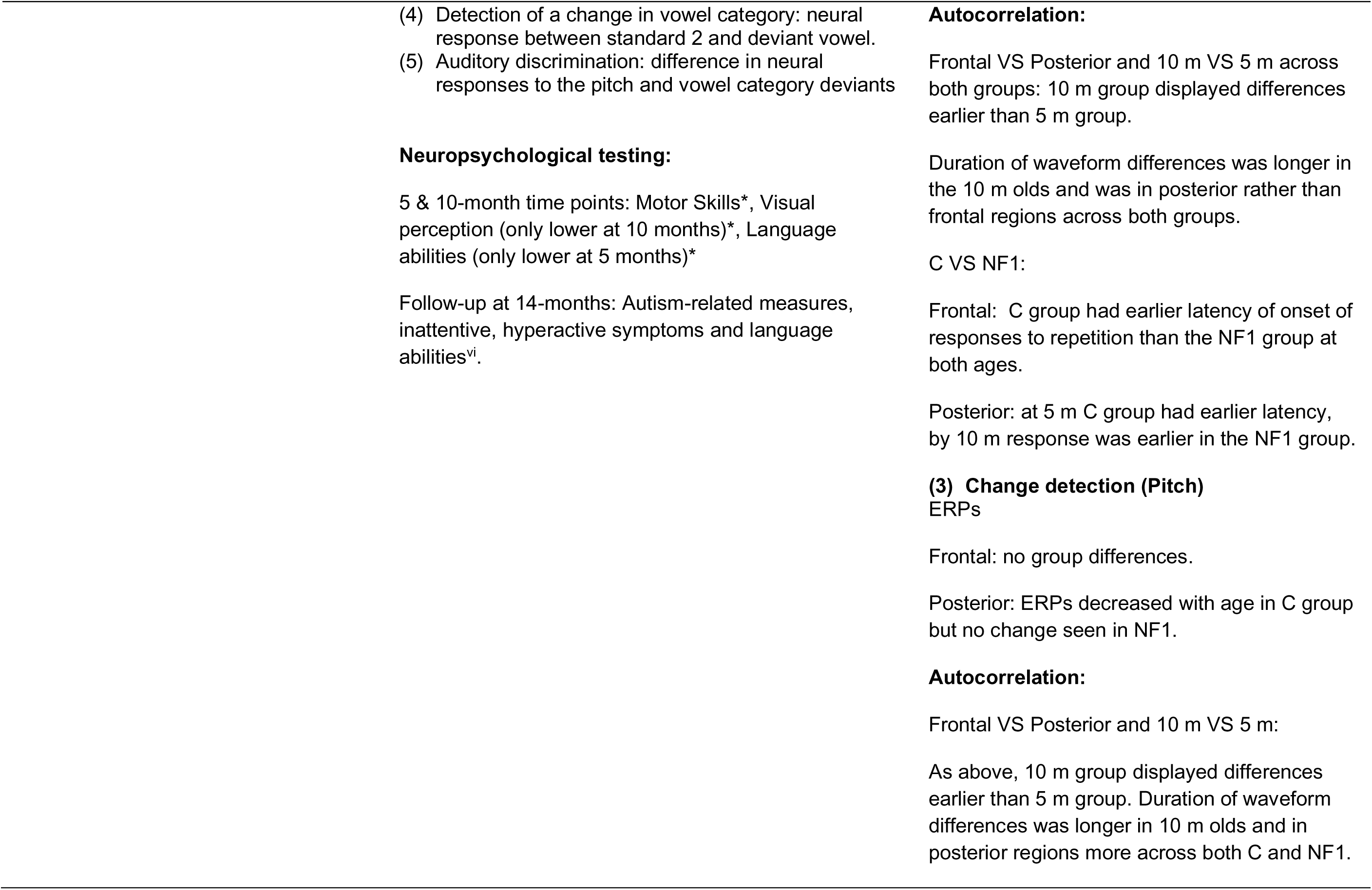
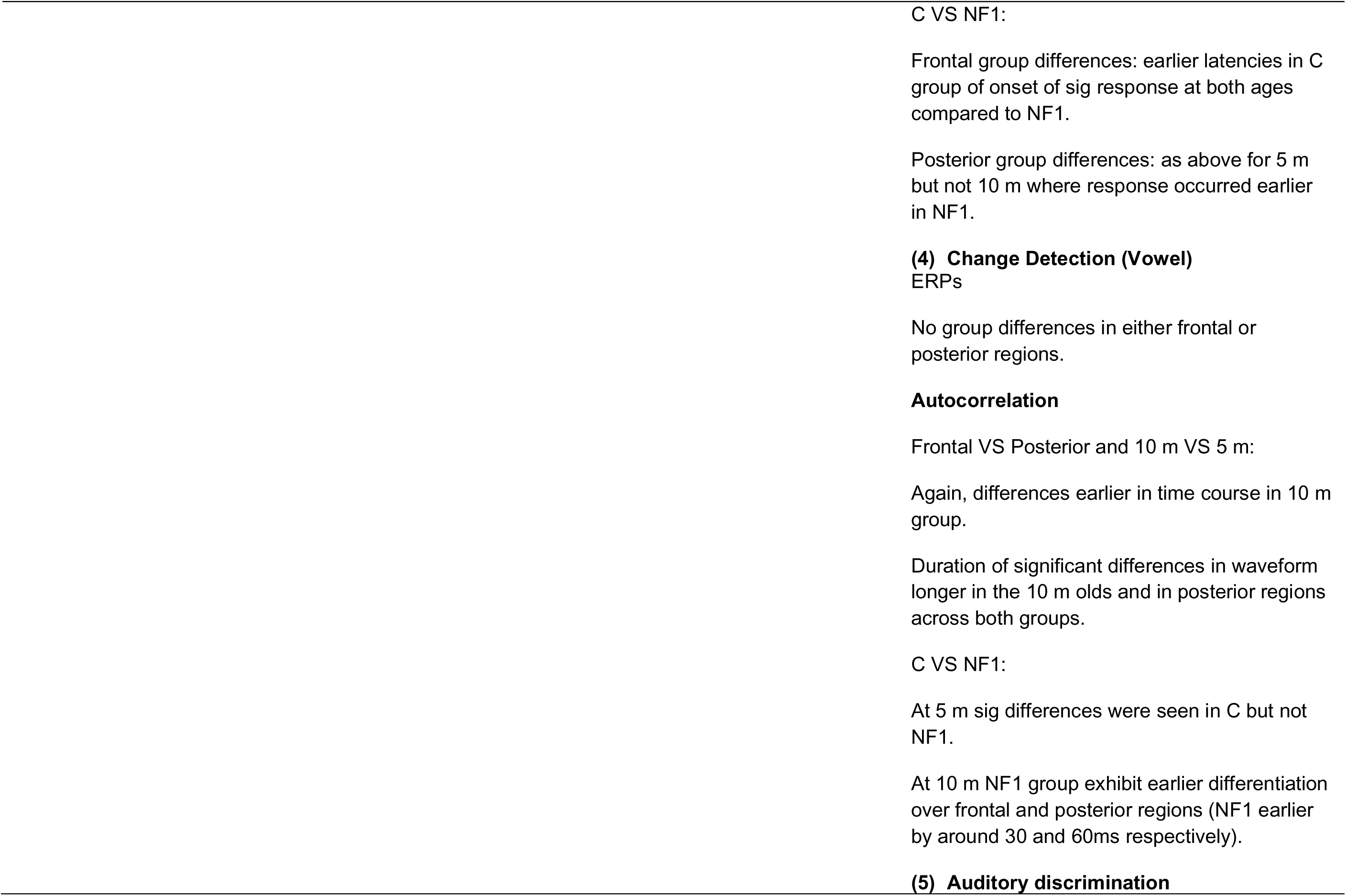
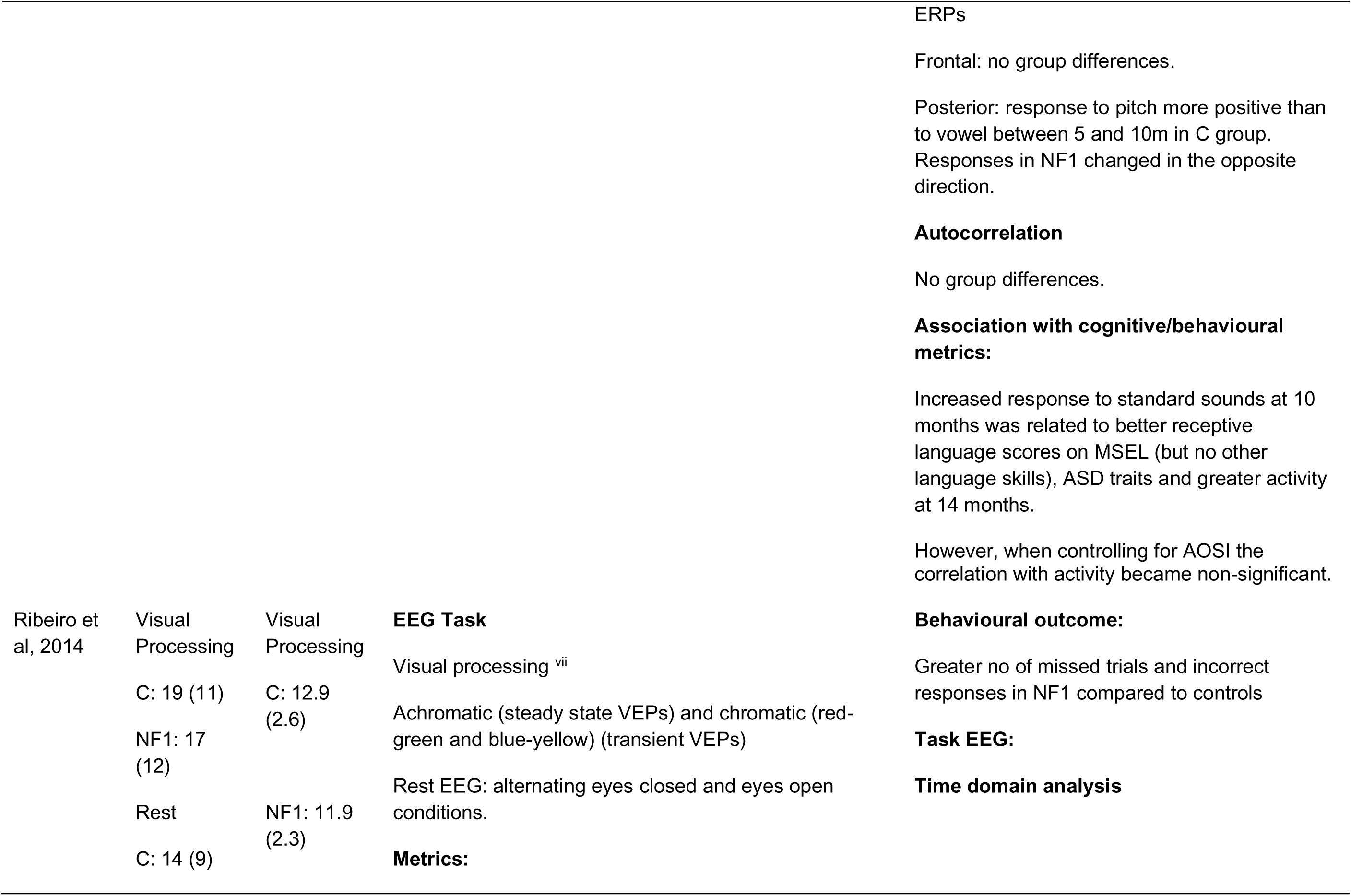
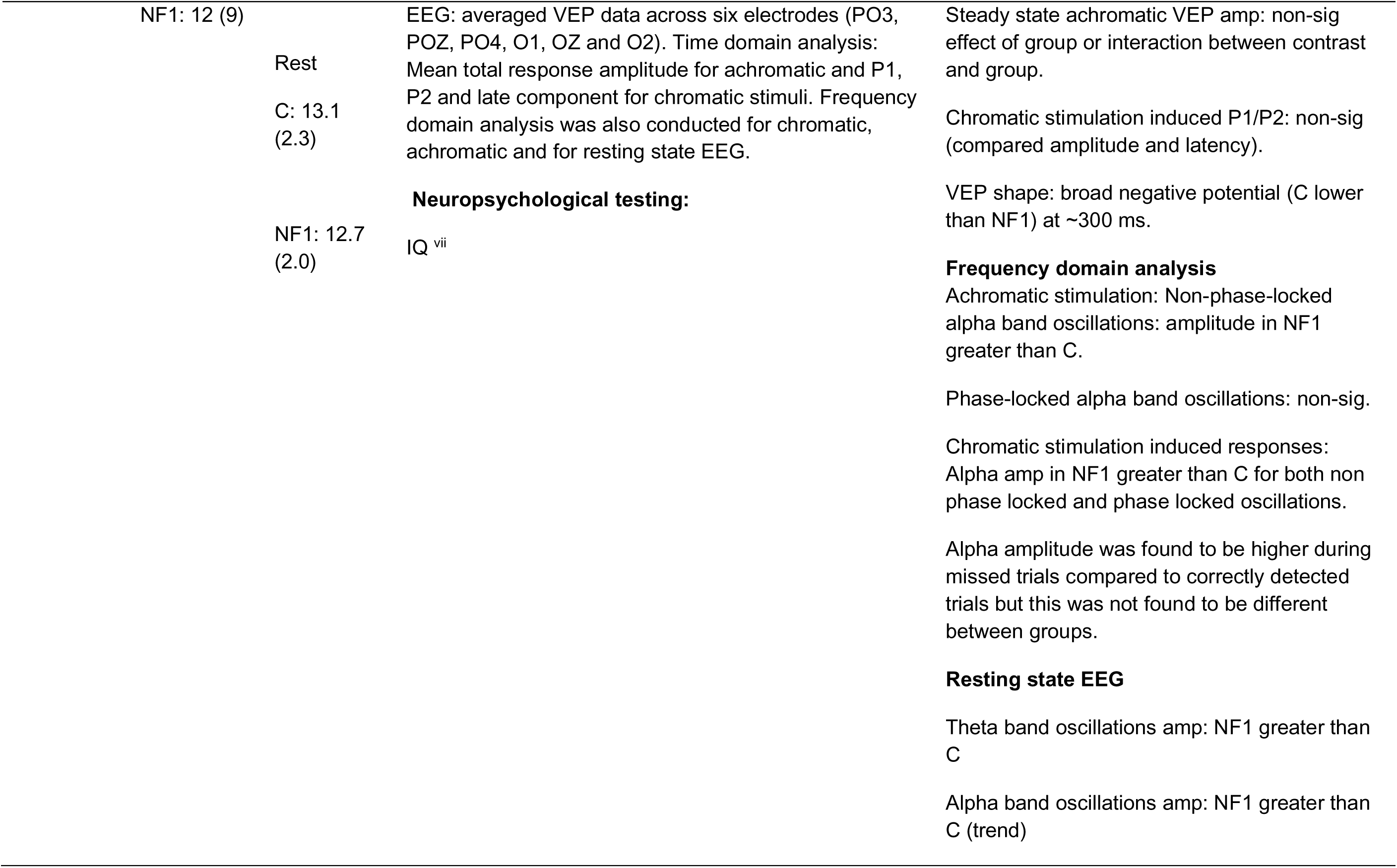
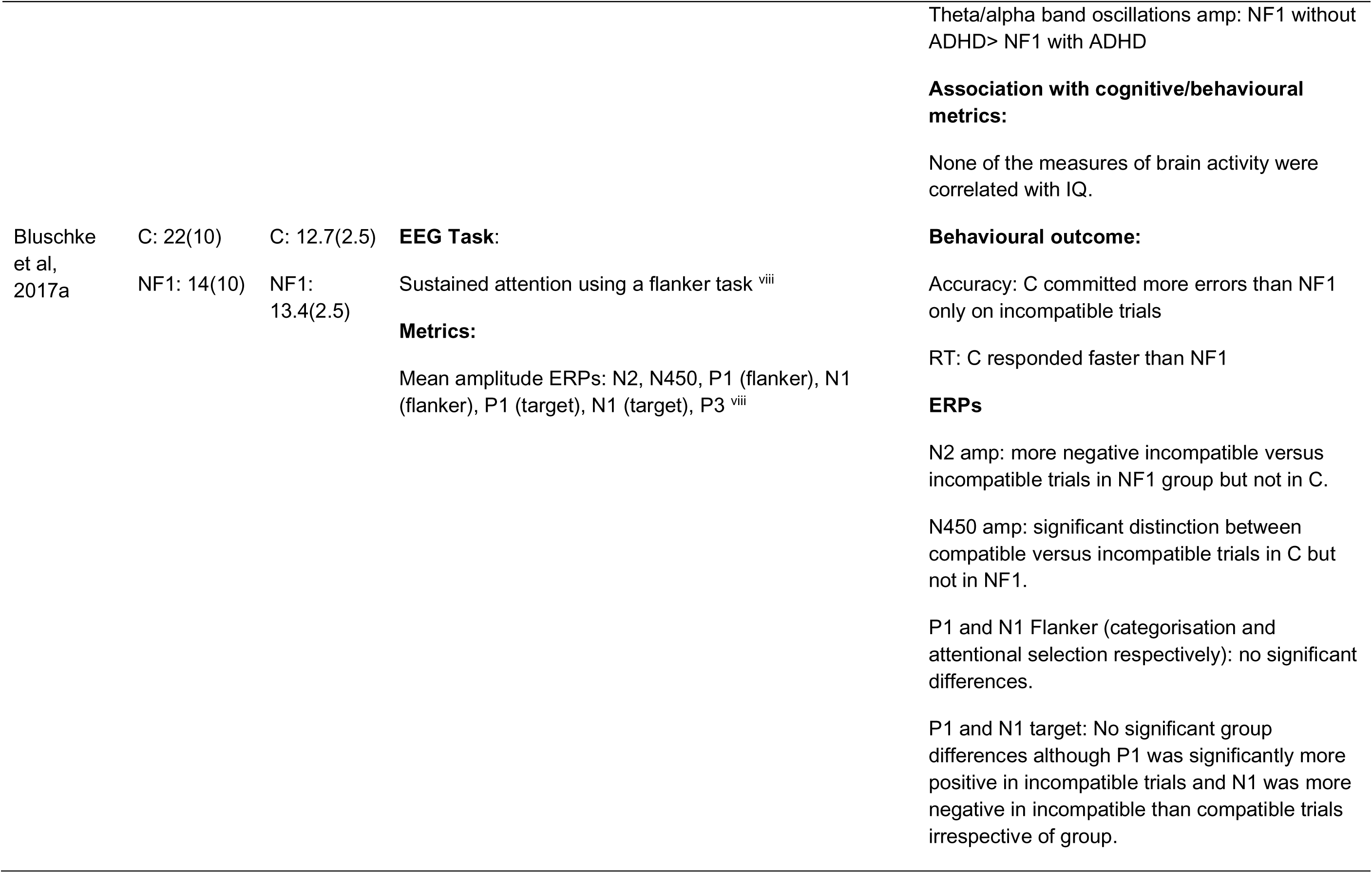
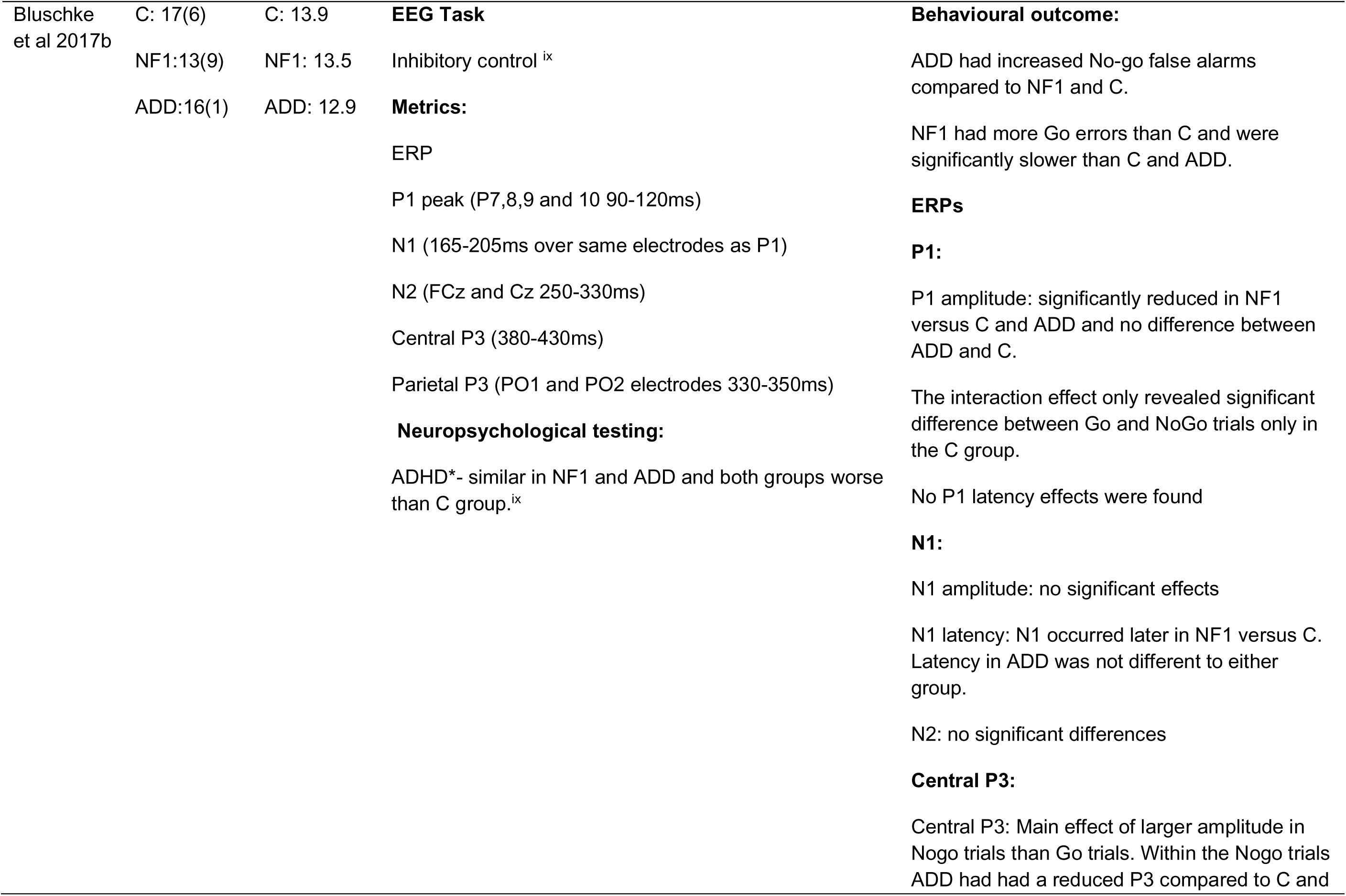
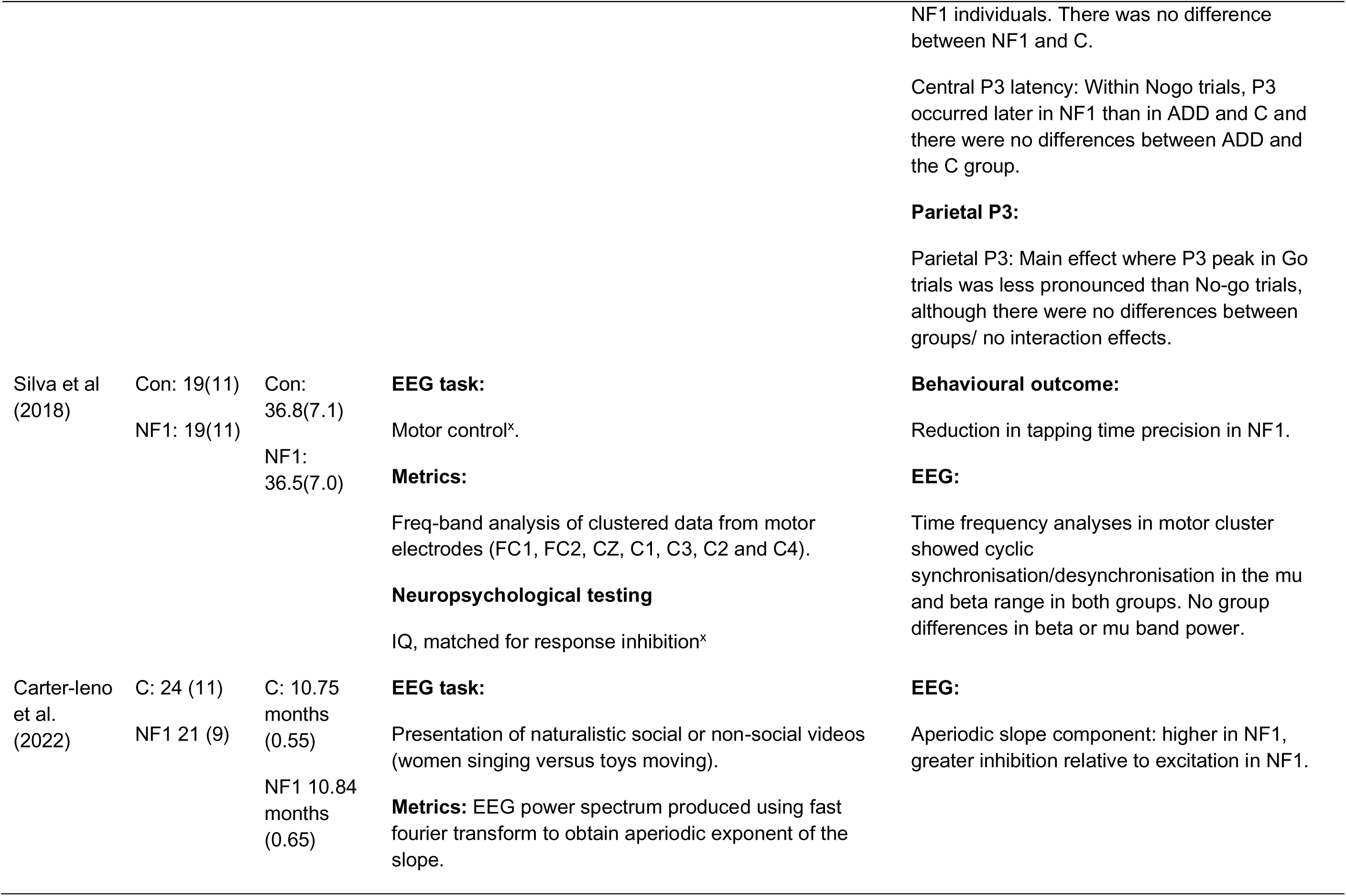
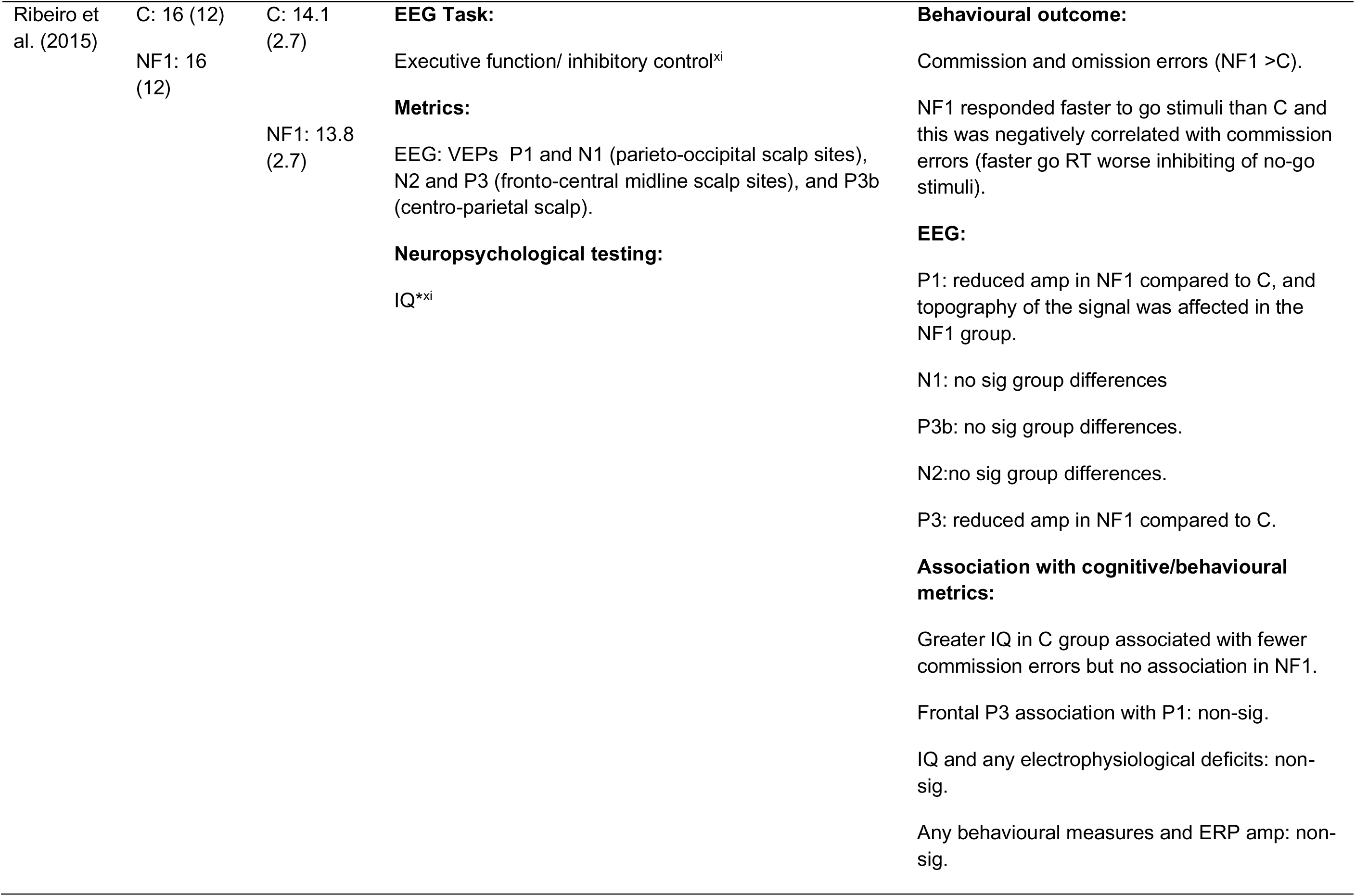
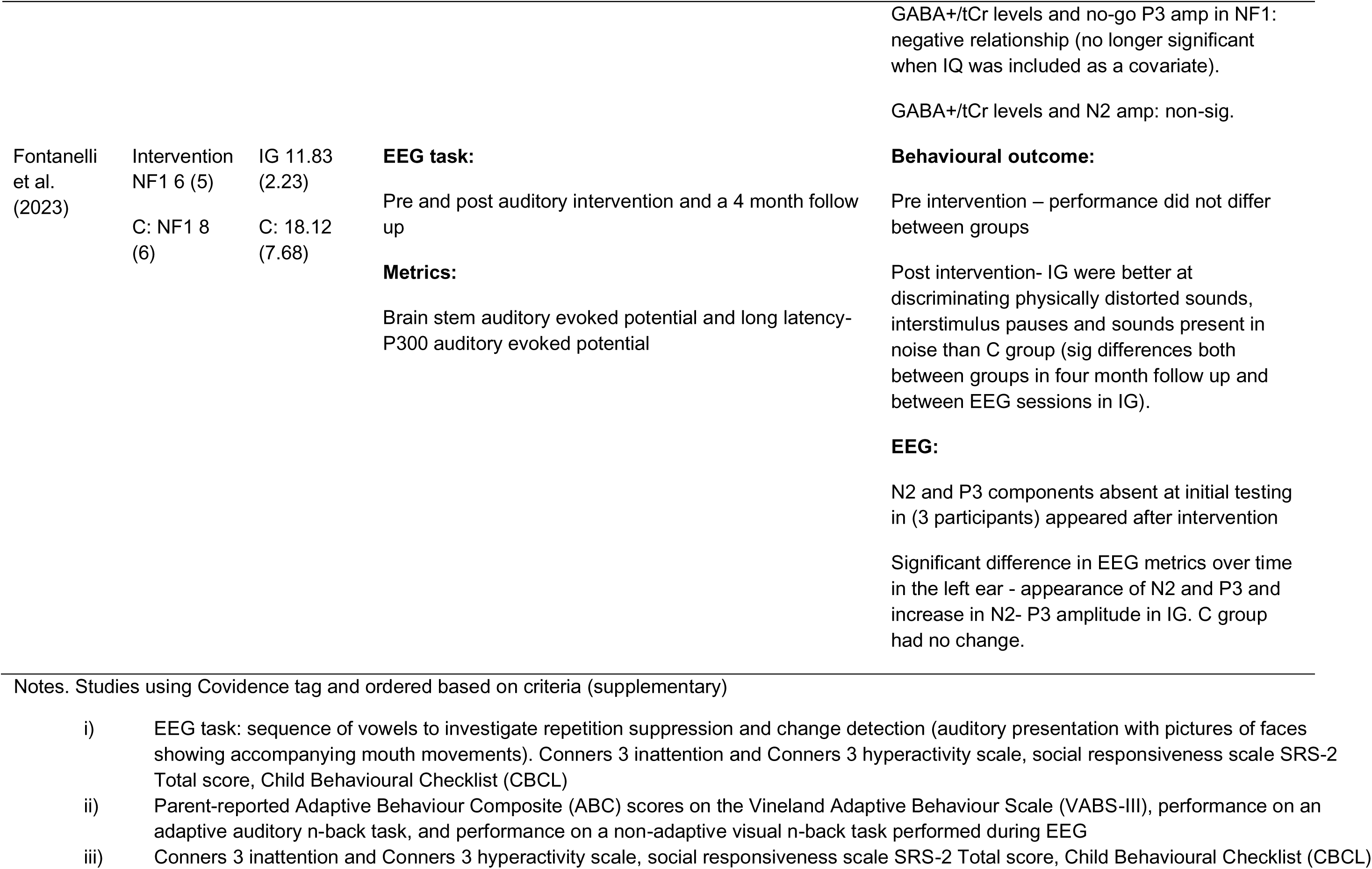
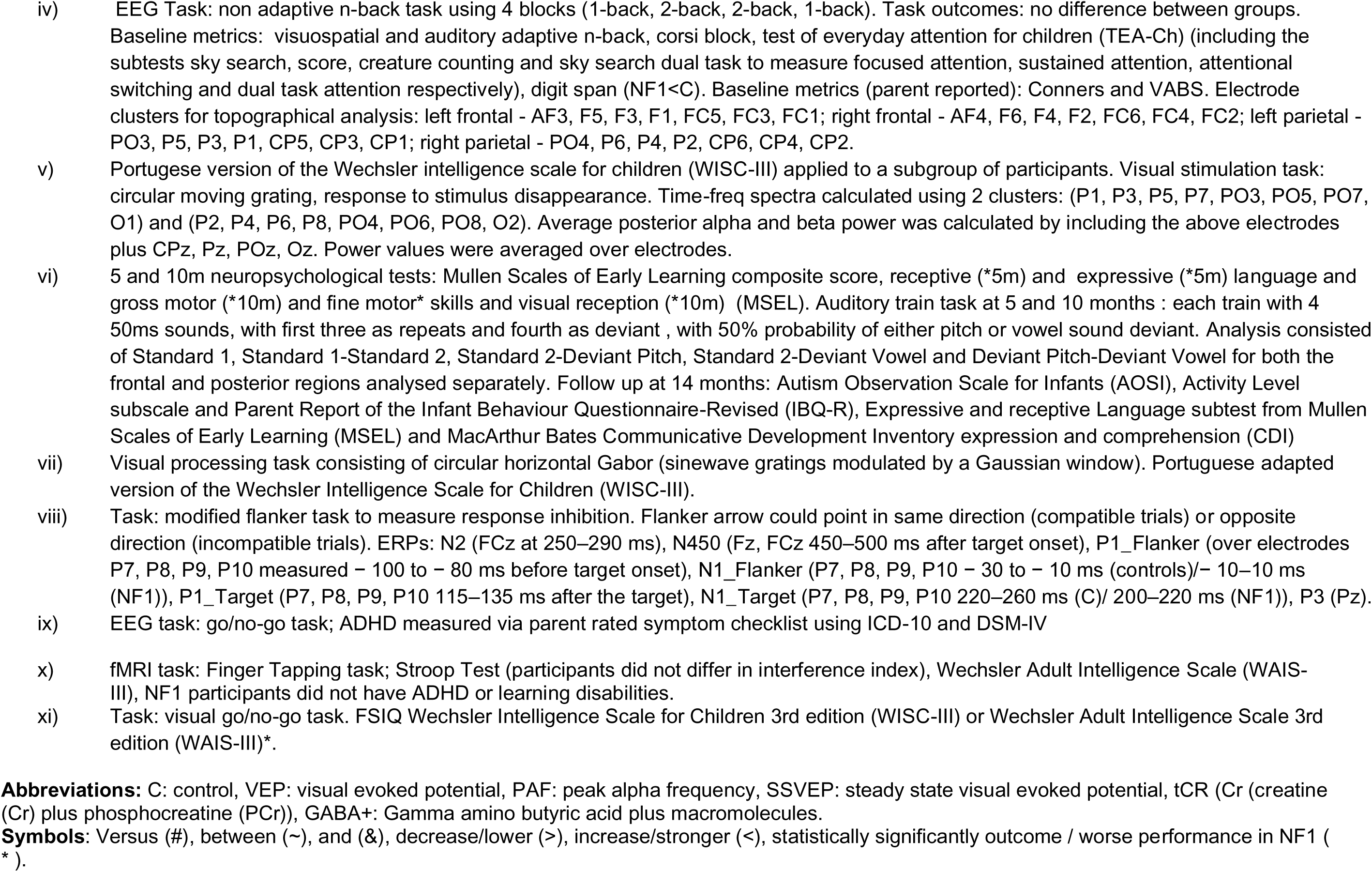
EEG studies.

### 3.4 Studies assessing brain metabolism and perfusion (PET, MRS, ASL)

#### 3.4.1 MRS

**Definition Box 4.** MRS terms used in the text.

|  |  |
| --- | --- |
| Magnetic resonance spectroscopy (MRS) | A method used to quantify chemicals in the brain. |
| GABA+ | Combined measure of GABA and macromolecules. |
| Glx | Combined measure of glutamate and glutamine. |

##### GABA and glutamate

Two studies investigated baseline prefrontal GABA in NF1 and its relationship to cognitive performance. Ribeiro et al. (2015) found reduced GABA+/total creatine (tCr) in the medial frontal cortex (MFC) in children and adolescents with NF1 compared with controls, with no difference in the occipital cortex or for Glx/tCr. MFC GABA correlated with behavioural inhibition, such that higher GABA was associated with faster response time but more commission errors. This relationship was reversed in controls such that higher GABA was associated with a slower response time and less errors. In addition, in NF1 higher GABA was positively associated with general intellectual ability. Similarly, Garg et al. (2022) found higher prefrontal GABA was related to faster response times and better performance (measured by inverse efficiency scores) on a working memory task. Therefore, both studies suggest higher prefrontal GABA in NF1 may confer better task performance and general intellectual function. Ribeiro et al. (2015) also found a reduced frontal P3 amplitude in NF1 from EEG negatively correlated with MFC GABA/tCr on a visual go/no-go task in the patient group only. This is consistent with higher GABA reflecting a more impulsive response style as reduced P3 is associated with poor impulse control. However, this was no longer significant when IQ was included as a covariate.

Three studies investigated GABA levels in the occipital cortex in NF1. In a combined PET-MRS study in adults, Violante et al. (2016) found reduced GABA+ concentrations in the frontal eye fields and occipital cortex in NF1 patients compared to controls. Reduced occipital GABA in NF1 was also noted in two other studies (Lacroix et al., 2022; Violante et al., 2013a). NF1 patients exhibited reduced binding of GABA-A receptors in the parieto-occipital cortex, midbrain, and thalamus compared to controls and subsequently, GABA+ was negatively correlated with the density of GABA-A receptors in the frontal eye fields but not in the occipital cortex in NF1 (Violante et al., 2016). Based on these findings, the authors conclude that reduced GABA concentrations commonly observed in NF1 may reflect compensatory mechanisms of the brain in response to reduced neurofibromin expression. Using a low-level visual attention task, Violante et al. (2013a) found baseline occipital cortex GABA/tCr was significantly correlated with peak BOLD response in both NF1 and controls whereas, Glx/tCr was not. These findings reflect the regional specificity of GABA and Glutamate on cognitive performance.

Studies investigating Glutamate in NF1 show mixed results. For example, Violante et al. (2016) found reduced Glutamate in the occipital cortex in NF1 but no differences in the frontal eye fields. In contrast, Lacroix et al. (2022) found increased ratios of Glx/tCr in NF1 compared to controls in the left occipital cortex. However, two studies found no difference in Glutamate or Glx across occipital and prefrontal regions (Ribeiro et al., 2015; Violante et al., 2013a). Furthermore, when metabolites acquired from the left occipital cortex were compared to TMS measures of inhibitory control, the authors found greater Glx/tCr was associated with more facilitation and less intracortical inhibition (through ICF and SICI) and higher GABA/Glx ratios were associated with higher intracortical inhibition in both NF1 and controls.

##### Other metabolites

Metabolites other than Glutamate and GABA have also been investigated in NF1. Wang et al. (2000) reported reduced levels of N-acetylaspartate (NAA), NAA/choline (Cho), NAA/total creatine in the thalamus, as well as reduced tCr in occipital grey matter in NF1 children 6-19 years old compared to controls regardless of whether patients exhibited T2 hyperintensities in the globus pallidus. Interestingly, NAA seemed to be relatively preserved in younger subjects (<10 years) but reduced in the older subjects (>10 years). A further study in adolescents found that NF1 patients had decreased NAA in the caudate nucleus and globus pallidum compared to controls and that NAA/Cho ratios were reduced in the putamen, caudate nucleus, and thalamus (Nicita et al., 2014). In adults, Violante et al. (2016) found reduced tNAA in the occipital cortex in NF1 compared to controls. However, they found no differences between NF1 and controls in the concentration of GPC (glycerophosphocholine), myo-inositol (mI), tCr, and tNAA in the frontal eye fields, nor in GPC, mI, or total Cr in the occipital cortex.

**Table 6.**
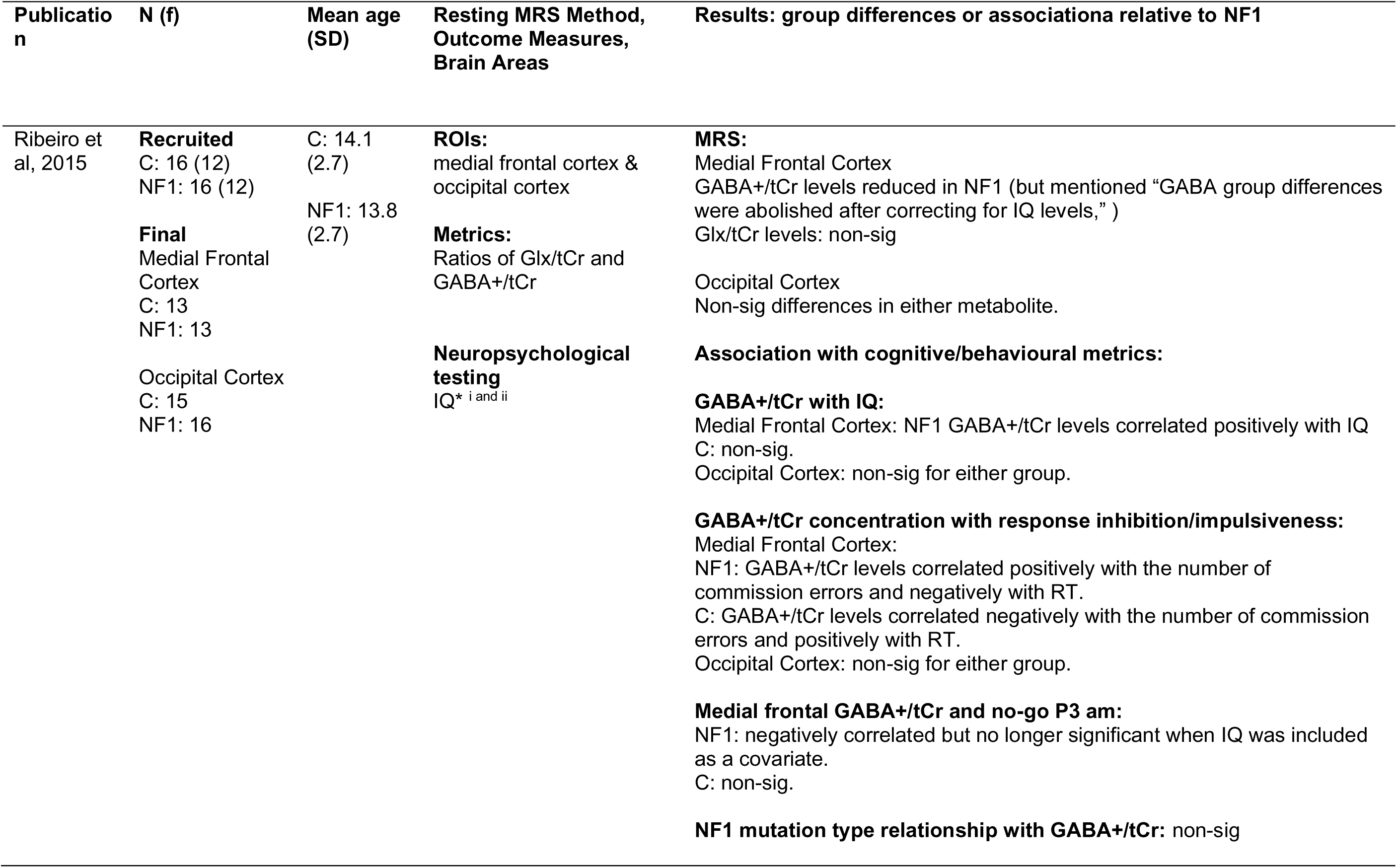
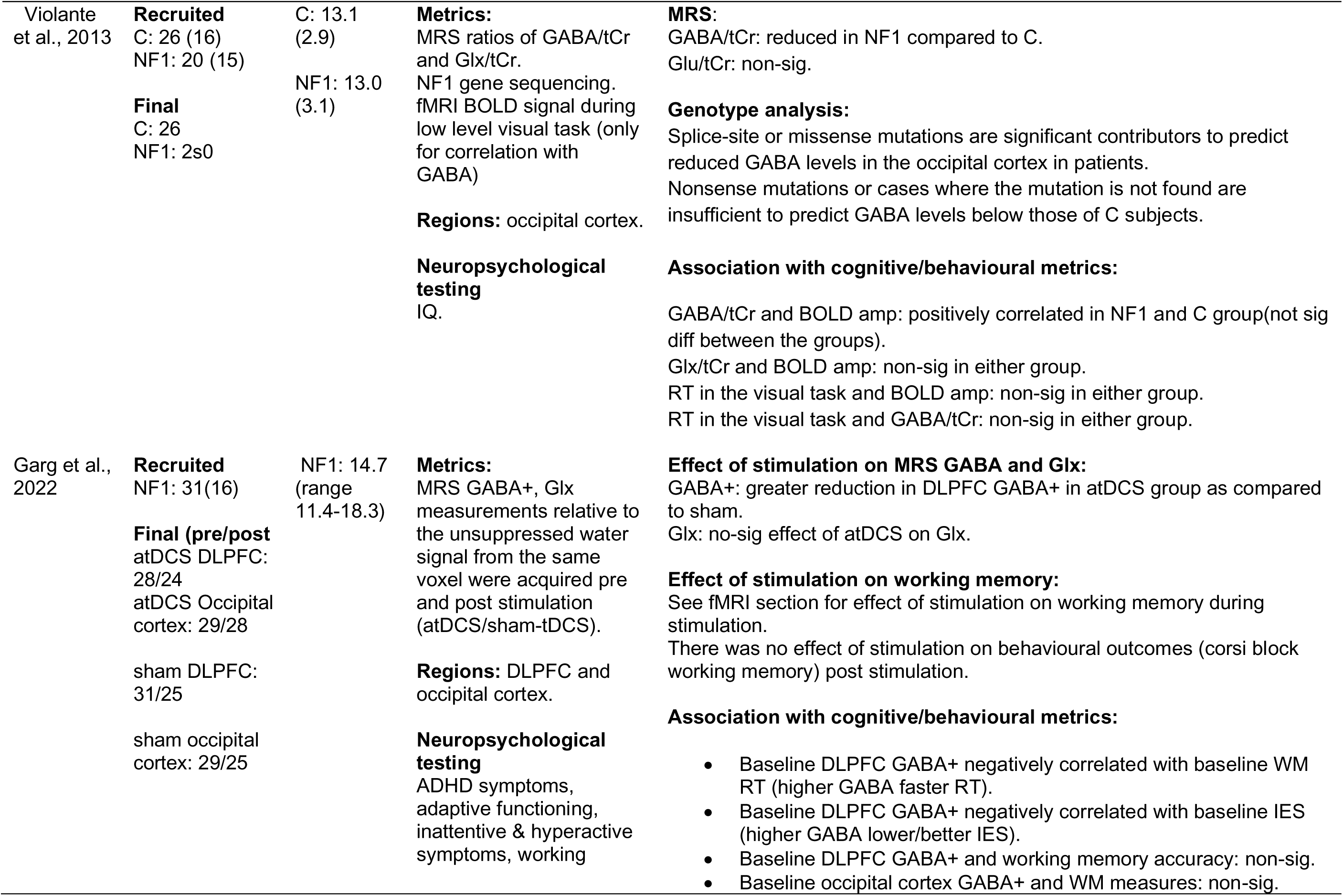
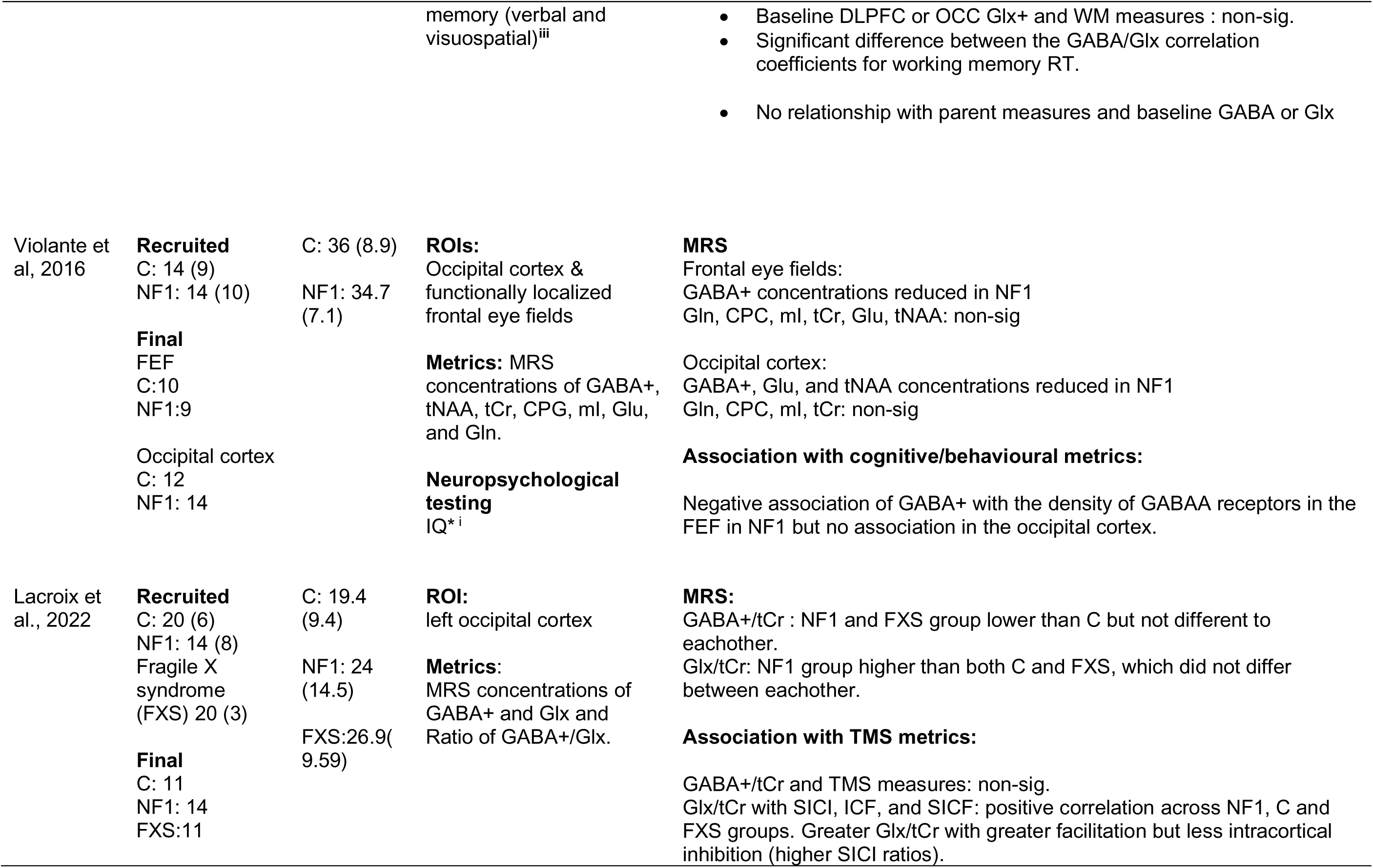
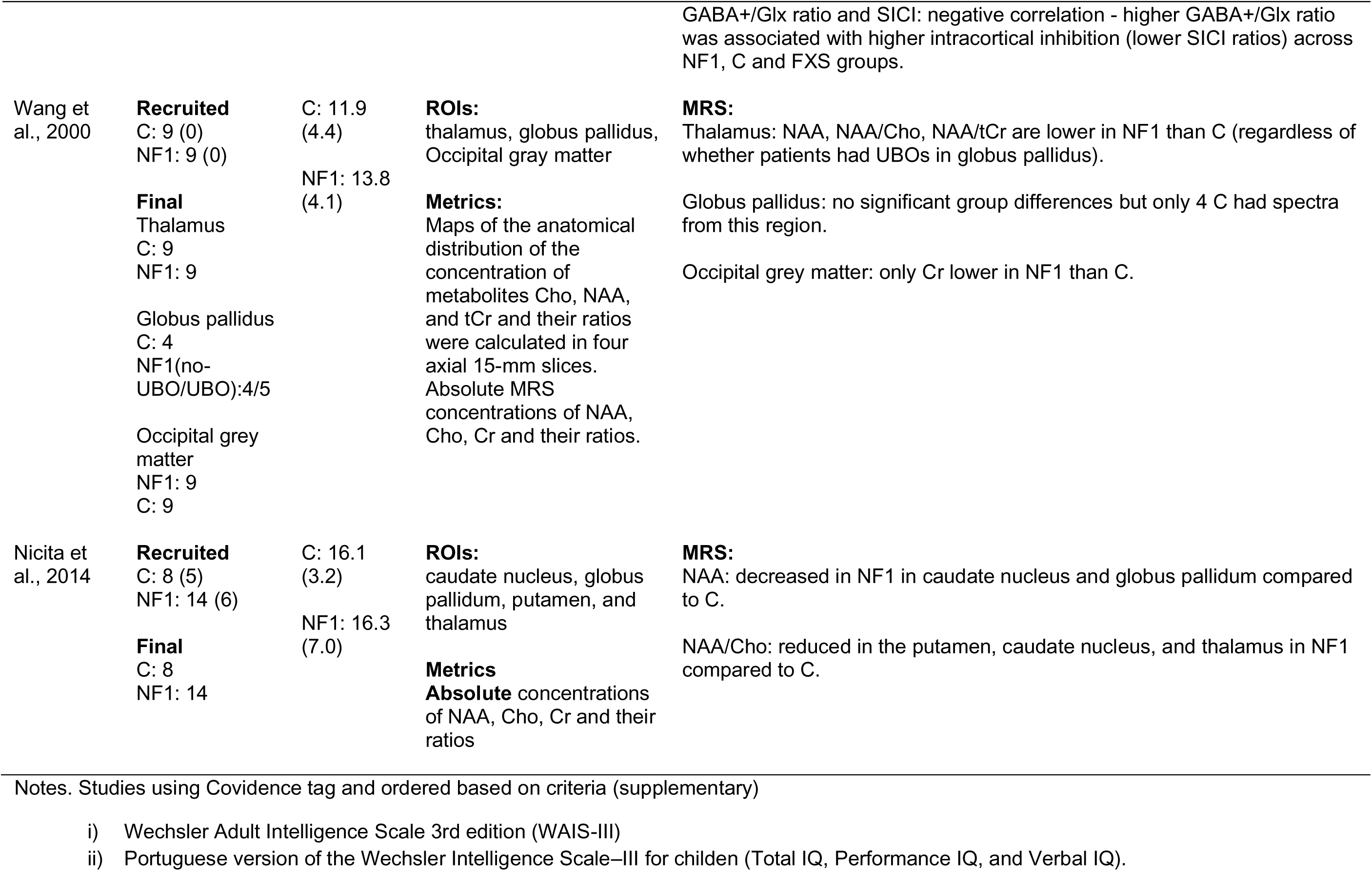
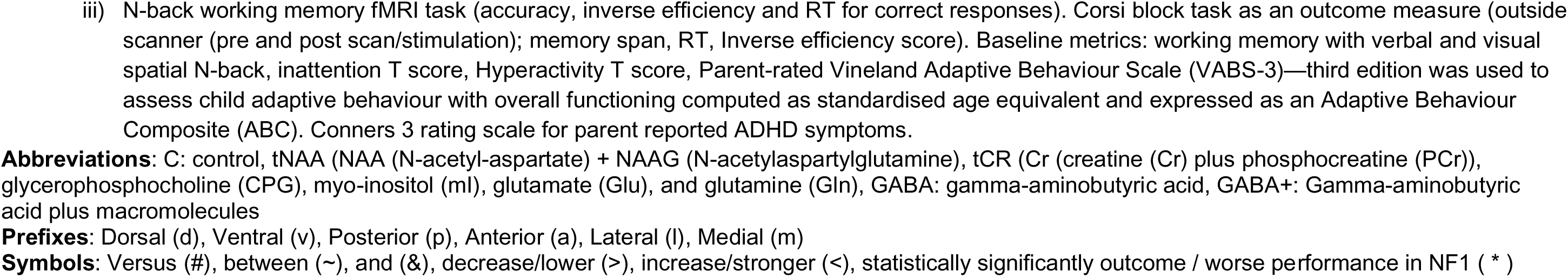
MRS studies.

#### 3.4.2 PET

**Definition Box 5.** PET terms used in the text.

|  |  |
| --- | --- |
| Fluorodeoxyglucose (FDG) PET | Maps glucose metabolism. |
| Flumazenil PET | GABA-A receptor agonist and therefore maps GABA-A receptor density. |

Schütze et al. (2018) examined how resting-state brain metabolism in NF1 was related to cognitive function. They found that resting metabolism measured by using 18F-fluorodeoxyglucose (FDG) PET/CT, was related to IQ, verbal memory, and verbal fluency with both positive and negative relationships globally. Similarly, Buchert et al. (2008) examined cerebral glucose metabolism with FDG PET in adults with NF1 and controls using both a whole-brain voxel-based analysis and ROI-based analysis. Both analyses showed reduced FDG retention in the thalamus in NF1, controlling for differences in the brain uptake period, which suggests reduced metabolic activity. Within group correlation analyses showed that this reduction in the NF1 group was not affected by comorbidities such as learning problems, tumours, or muscle hypotonia.

PET has also been used to investigate neurotransmitter receptor function. Violante et al. (2016) examined regional and whole-brain receptor binding, using [(11)C]-flumazenil PET, in NF1 and age and sex-matched control participants. The study reported that NF1 patients exhibited reduced binding of GABA-A receptors in the parieto-occipital cortex, midbrain, and thalamus compared to controls and the occipital cortex and frontal eye fields displayed reduced GABA+ concentration. Furthermore, GABA+ was negatively correlated with the density of GABA-A receptors in the frontal eye fields (FEF) but not in the occipital cortex in NF1, whereas no such significant association was evident in controls. The authors suggested that the relationship between GABA and GABA receptor expression is therefore not usually tightly linked in the TD brain, explaining why GABA decrease and GABA receptor decrease identified in this study seem to be two separate mechanisms across different regions.

#### 3.4.3 ASL

**Definition Box 6.** ASL description.

|  |  |
| --- | --- |
| Arterial spin labelling (ASL) | Non-invasive perfusion technique to determine cerebral blood flow. |

(Wolters et al., 2017; Yeom et al., 2013)

Yeom et al. (2013) used a magnetic resonance perfusion method to examine regional cerebral blood flow (CBF) differences in children with NF1 and controls. This revealed that the NF1 group showed hypoperfusion in 7 areas relative to controls **(Table 7)**.

**Table 7.**
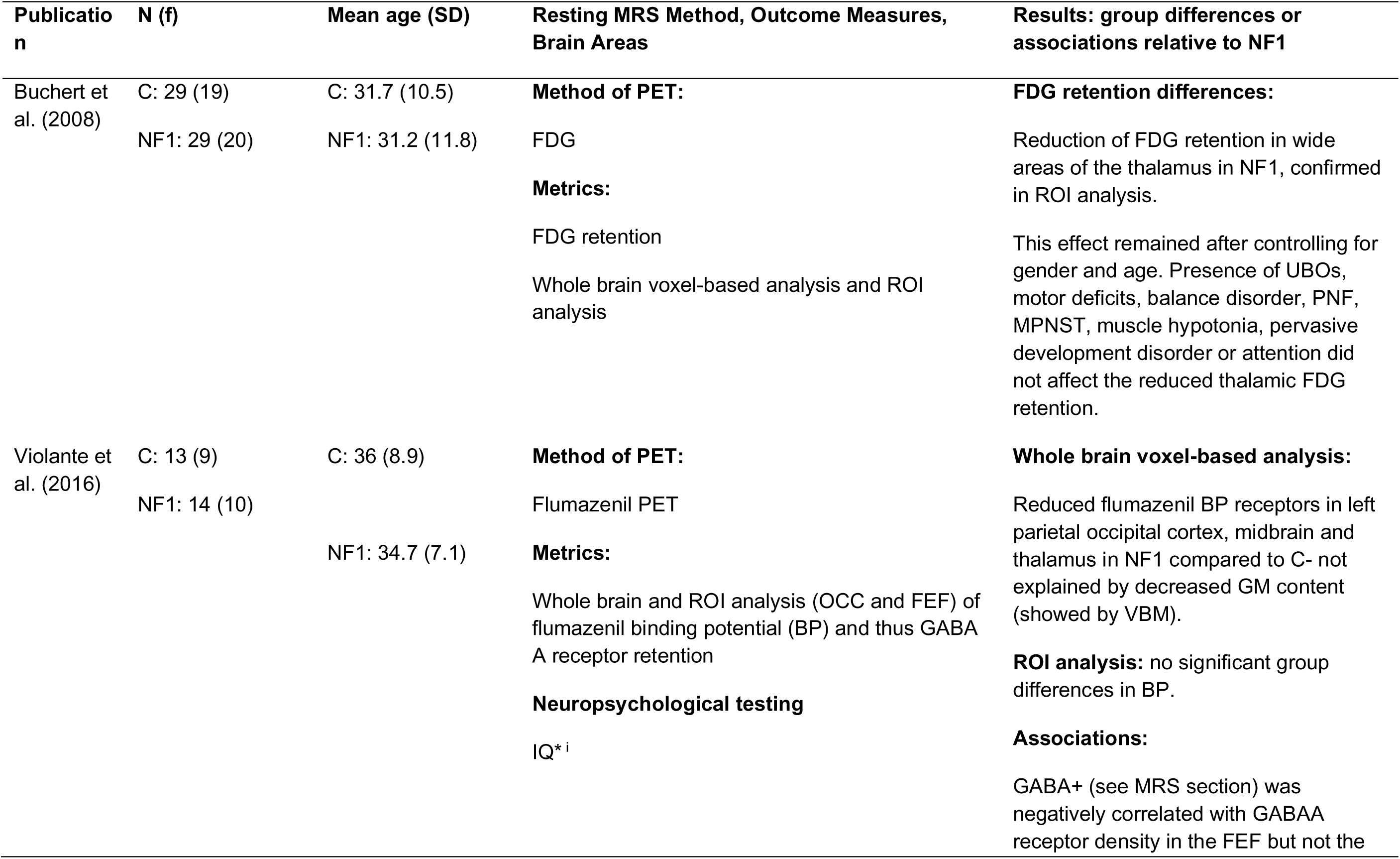
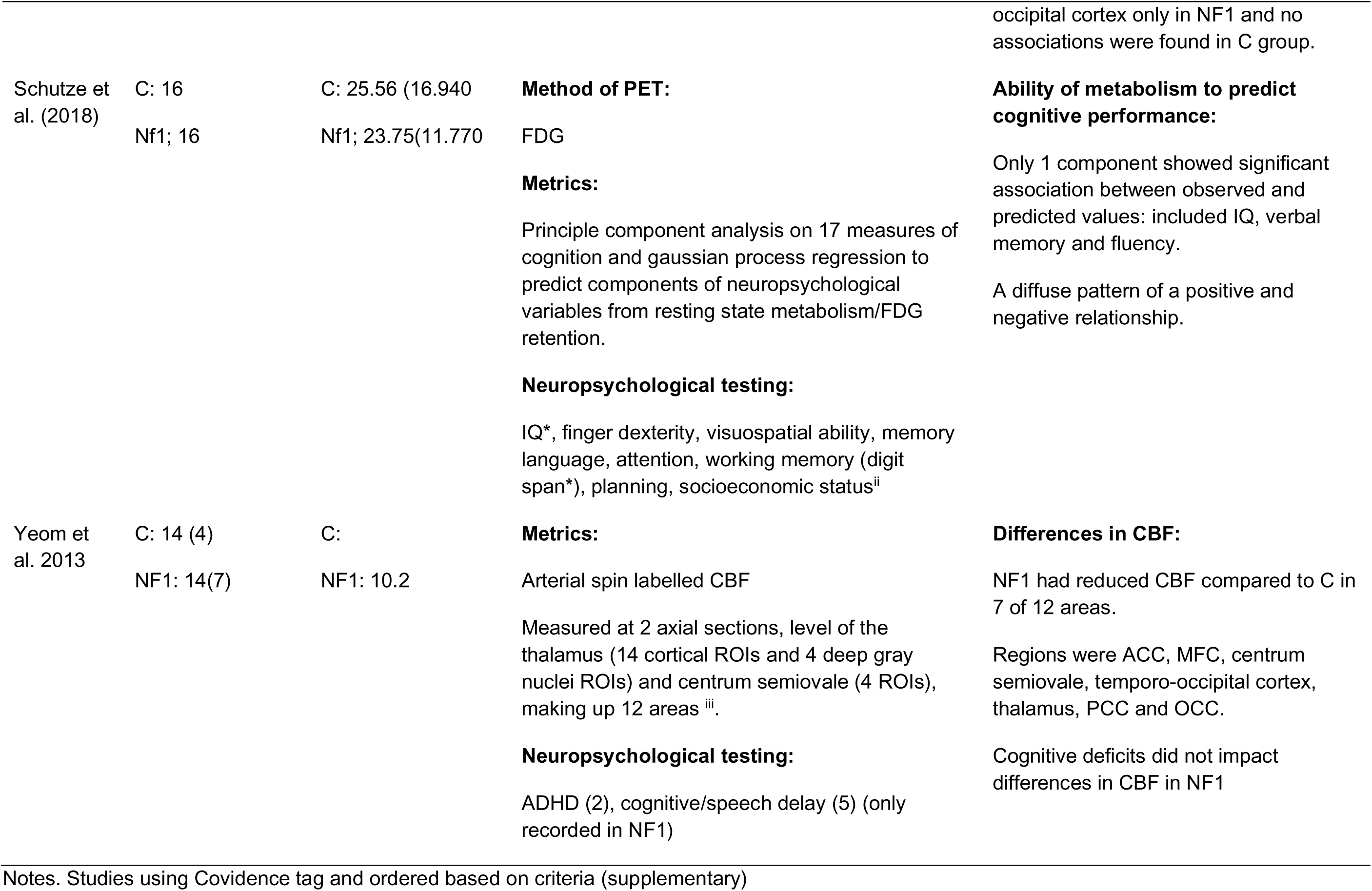
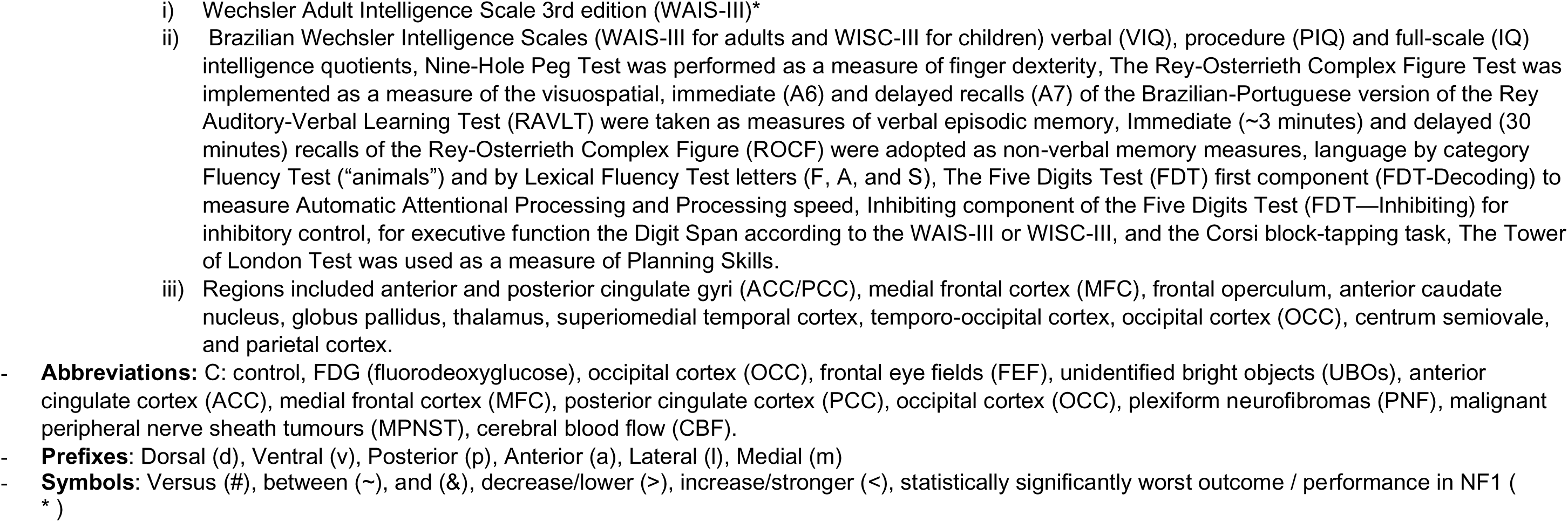
PET and ASL studies.

## 4 Discussion

In this review, we have systematically evaluated results from 44 studies across the lifespan employing functional, metabolic, and neurophysiological research methods to understand the pathophysiological mechanisms underlying cognitive function in NF1. Overall, the results indicate heterogenous and widespread alterations across brain regions, but with a lack of specificity and sensitivity of neuroimaging metrics to domain-specific cognitive functions. Below we discuss the results from the review, synthesising the findings under the broad categories of dysregulated E/I in NF1, changes in brain connectivity and the role of the DLPFC.

### 4.1 Dysregulated excitation/inhibition underlie learning difficulties in NF1

Whilst the extant literature in NF1 suggests excitation/inhibition imbalance, the findings across *Nf1* animal models and in clinical cohorts are inconsistent. Integrating findings from MRS, TMS, EEG and PET research included in this review can help clarify the underlying mechanisms and examine the dysregulated GABAergic inhibition in NF1 clinical cohorts.

The MRS studies in NF1 suggest that GABA levels measured at rest in the frontal and visual cortex are reduced (Ribeiro et al., 2015; Violante et al., 2016; Violante et al., 2013a), and that higher GABA is associated with faster responses on cognitive tasks and better intellectual abilities (Ribeiro et al., 2015). This was also supported by Garg et al. (2022), in an experimental study which involved application of anodal tDCS stimulation to the left frontal lobe in NF1. Application of anodal tDCS was associated with a reduction of GABA, and worse working memory task performance. In contrast, Carter Leno et al. (2022) investigated E/I by examining the aperiodic slope exponent of the EEG power spectrum and found higher inhibition in NF1 infants compared to TD controls. It should be noted that the study by Carter Leno et al. (2022) was in infants as compared to the older cohorts in the MRS studies.

TMS studies included in this review used to investigate GABAergic inhibition report mixed findings. Mainberger et al. (2013) found increased SICI in NF1 as compared to controls suggesting increased intracortical inhibition. Furthermore, the study demonstrated that using lovastatin to target the Ras pathway overactivity in NF1 normalised SICI and was associated with increases in attentional performance. In line with Mainberger et al. (2013), Doherty et al. (2023) found a decreased E/I ratio in NF1 but through lower ICF (suggesting reduced excitation). Importantly, better PANESS scores for motor function in NF1 occurred with a lower E/I ratio, suggesting that more inhibition and less excitation in NF1 may not be maladaptive, which contradicts Mainberger et al. (2013)’s findings. Interestingly, research in other conditions suggests greater SICI (higher inhibition) reflects better motor function and reduced SICI has been reported in ADHD, Alzheimer’s disease, and Parkinson’s disease (Gilbert et al., 2019; Mimura et al., 2021; Ni et al., 2013). Finally, two studies reported reduced plasticity in NF1, reinforcing the idea of dysregulated E/I.

Task specific modulation (i.e reduction in inhibition prior to the start of a cognitive task) is also reduced in NF1. Zimerman et al. (2015) found that whilst the control group exhibited a reduction in intracortical inhibition prior to movement onset, associated with better motor learning, this task-specific modulation was less pronounced in the NF1 group, such that there was not such a decrease in inhibitory activity. Lastly, PET results seem less definitive but in line with widespread disruption to the E/I balance. For instance, FDG retention is reduced in the thalamus in NF1 (Buchert et al., 2008), suggesting reduced excitatory activity and reduced E/I balance. However, reduced GABA receptor binding suggests decreased inhibition (Violante et al., 2016), although this finding and relationship to GABA concentration varies depending on the region and may be a compensatory response to increased GABA release.

Overall, the results across studies in NF1 suggest dysregulation of E/I balance and impairments in plasticity, although the direction of this effect (increased or decreased E/I balance) is less clear. The relationship of E/I with cognition is also unclear; for instance, a lower E/I ratio in NF1 is reported to be associated with worse alertness and motor learning (Mainberger et al., 2013; Zimerman et al., 2015) but also faster RT on working memory tasks, higher IQ and better motor development (Doherty et al., 2023; Garg et al., 2022; Ribeiro et al., 2015). The combined reduction in GABA concentration and GABA receptor expression found in different regions in NF1 may represent compensatory mechanisms for increased synaptic GABA release (Violante et al., 2016). However, this does not explain why, for instance in prefrontal regions, having higher GABA levels seems to be beneficial in NF1 (Garg et al., 2022; Ribeiro et al., 2015). As inhibitory circuits are affected from early development, function is likely distinct from the TD brain, and consequently there is an abnormal and inconsistent E/I relationship with cognitive function. It should be noted that there are key differences across the studies which may impact the results such as the age of the cohort and the brain region being studied. Most of the studies included in the age group are in the adolescent and adult NF1 cohorts apart from one study reported in infants with NF1. Furthermore, TMS studies have investigated motor learning with application of stimulation to the motor cortex whereas the MRS studies report findings in the frontal or occipital cortex. In addition, there are differences based on whether the measures were collected at rest or during performance of a task. MRS GABA reported in studies likely represents a more tonic rather than phasic aspect of inhibition and therefore representing a different aspect of the E/I balance to a measure such as TMS (Dyke et al., 2017).

Animal models of *Nf1* can provide an important tool for causally testing hypotheses about biological processes, especially when using approaches that seek to closely approximate functional alterations observed with brain imaging techniques in patients. Goncalves et al. (2017) tested the E/I hypothesis in NF1 mouse model using MRS techniques similar to those used in human clinical cohorts. As compared to wild-type mice, the study found reduced GABA in the hippocampus but not in the prefrontal cortex or striatum and this was accompanied by a significant increase in GABA-A receptor densities in the hippocampus. Glutamate activity was reduced both in the hippocampus and striatum indicating disrupted E/I in a region-specific manner. Overall, the E/I ratio was reduced in the prefrontal cortex and striatum relative to wild type mice but not significantly different in the hippocampus. Furthermore, a recent study reported sex-specific differences in MRS GABA concentrations in an *Nf1^+/-^* mouse model (Santos et al., 2023). Juvenile female *Nf1^+/-^* mice exhibited increased hippocampal GABA levels alongside better memory and social behaviour whereas no difference was found in male mice and instead they showed decreased GABA receptor expression. Overall, these results suggest that the E/I differences are brain region-specific, and sex should also be considered an important determinant of the NF1 phenotypic expression (Santos et al., 2023). Finally, GABA levels are known to change with age with a recent study demonstrating increase in frontal GABA in childhood, followed by a period of stability during adulthood and a gradual decrease with ageing in a TD population (Porges et al., 2021). Therefore, the age of the population under study should also be an important determinant to consider.

The E/I hypotheses has been implicated in neurodevelopmental conditions like ASD but also in mental illnesses like schizophrenia (Canitano & Pallagrosi, 2017). Technical developments such as functional MRS in humans may help provide a more nuanced understanding of E/I by measuring GABA and Glutamate concentration changes during task engagement (Bell et al., 2023; Pasanta et al., 2023), this could also be complemented by developing use of PET markers of E/I balance, such as both flumazenil PET and ABP688 PET for glutamate receptor availability. Finally, E/I imbalance may be a mechanistic oversimplification as brain disorders can arise from multiple neural circuit alterations. Computational modelling approaches in addition to the empirical studies may build further on our understanding of the neurobiological mechanism affecting cognition in NF1.

### 4.2 Brain connectivity in NF1

Resting-state FC studies have allowed the investigation of activity signals between brain regions in the absence of a task or stimulus (Elorette et al., 2024). The studies included in this paper suggest a pattern of altered connectivity in the NF1 brain. Key findings include reduced anterior-posterior connectivity and increased local connectivity (Billingsley et al., 2004; Chabernaud et al., 2012; Mennigen et al., 2019; Shilyansky et al., 2010; Shofty et al., 2019; Violante et al., 2012). The intra-domain hyperconnectivity was reported in the cognitive domain (Mennigen et al., 2019), within the limbic network (Shofty et al., 2019) and right angular gyrus (Baudou et al., 2022). Interestingly, similar findings of diminished long-range connectivity and local over-connectivity have been observed in autism spectrum conditions and schizophrenia (Courchesne & Pierce, 2005; Lottman et al., 2019), although recent theories suggest local frontal connectivity is not always increased (Liloia et al., 2024; Vissers et al., 2012). The convergence of these findings across conditions may suggest shared downstream circuit level alterations that underpin cognitive functions; for instance, working memory difficulties which are observed across different conditions.

Task-based fMRI studies investigating connectivity have demonstrated reduced deactivation of DMN regions during task performance (Ibrahim et al., 2017; Violante et al., 2012). DMN regions are associated with self-referential thought and deactivate during demanding cognitive tasks. In accordance with this, less deactivation is associated with lapsed attention. Ibrahim et al. (2017) showed reduced connectivity between PCC and temporal regions indicating failure to deactivate the DMN leading to task interference. This DMN interference may contribute to the attentional deficits seen in NF1.

Intracortical myelination plays a significant role in the development of neural circuits and functional networks and has been shown to underlie atypical network connectivity patterns in autism (Chen et al., 2022). Myelination improves the brain’s data processing capacity, increasing neural efficiency and conduction speed of action potentials and synchronises neural signals. Myelin plasticity, both in grey and white matter, is an important regulator of learning and memory (de Blank et al., 2023; McKenzie et al., 2014; Pan et al., 2020). NF1 is associated with white matter abnormalities that can be visualised on T2 weighted scans as focal areas of signal intensity. These T2 hyperintensities are thought to be due to myelin vacuolation or areas of demyelination (de Blank et al., 2023). Although there are no studies directly investigating the relationship between myelination and cognitive function in NF1, it is possible that the abnormal myelination in NF1 contributes to the FC differences seen. Indeed, Nemmi et al. (2019) found that mean diffusivity was the best metric at discriminating NF1 from control group compared to functional metrics.

### 4.3 Differential neural activation patterns in NF1

Task-based fMRI and EEG studies demonstrate widespread differences in neural activation patterns in the NF1 cohort as compared to controls. The differences in experimental protocol across studies makes it difficult to synthesise the results; however, regions that differ between controls and patients with NF1 tend to be associated with attentional control, inhibition and working memory capacity. Taken together, the group differences suggest that the key brain areas including anterior cingulate cortex, temporoparietal junction and inferior frontal gyrus show reduced activation during task performance. Paradigms probing attentional and impulsivity processes reveal that anterior cingulate cortex and inferior frontal gyrus hypoactivation relates to poorer sustained and selective attention and attentional control in NF1 (Pride et al., 2017; Pride et al., 2018).

Hypoactivation of the frontal lobe has been described during reading, visuospatial and working memory tasks (Billingsley et al., 2004, 2003; Ibrahim et al., 2017; Pobric et al., 2022; Shilyansky et al., 2010). Ibrahim et al. (2017) compared neural activation differences in high versus low memory loads and found more diffuse brain activation in the frontal, parietal and temporal regions possibly suggesting a less efficient pattern of neural activity. Similarly, in a topographic analysis of P300 amplitude during a working memory task, Pobric et al. (2022) found imbalance in the distribution of neural activity across the frontal and parietal cortex in the NF1 group, with a weaker left frontal positivity and a stronger left parietal positivity as compared to controls. There was also neural activation differences reported in the cerebral hemisphere recruitment during task performance; Clements-Stephens et al. (2008) demonstrated a predominant left hemispheric recruitment in NF1 as opposed to the right hemisphere in controls. The findings of frontal lobe hypoactivation is perhaps not surprising given that the white matter microstructural alterations in NF1 are reported to be most pronounced in the frontal lobe (Karlsgodt et al., 2012).

Two studies investigated attentional processes associated with alpha rhythms in NF1 adolescents (Ribeiro et al., 2014; Silva et al., 2016). Silva et al. (2016) used a covert attention task and found greater desynchronisation of alpha oscillations in NF1. This is suggestive of compensatory mechanisms to regulate attention in NF1, as alpha suppression is thought to be a sign of active neural processing. In addition, Ribeiro et al. (2014) found enhanced amplitude of alpha oscillations relative to controls during low level visual processing, a characteristic associated with poorer attentional performance. Due to the role of the thalamus in generating alpha rhythms (Hughes & Crunelli, 2007), the authors suggest the abnormal thalamus in NF1 may contribute to this pattern of enhanced alpha rhythms (Ribeiro et al., 2014). Furthermore, both Ribeiro et al. (2015) and Bluschke et al (2017a) used a go-no go task to investigate impulsivity and response inhibition in NF1. Both studies found a reduction in P1, reflecting visual cortex activation, suggested to reflect poor attentional allocation or visual processing and Ribeiro et al. (2015) also found a reduction in frontal P3 amplitude, which is associated with inhibitory control, suggestive of poor self-regulation. Overall, abnormal alpha rhythms and reduced EEG components P1 and frontal P3 suggest deficits in allocating attention and regulating an appropriate response.

Aside from abnormal task-based activation, two studies (Booth et al., 2023; Ribeiro et al., 2014) investigated resting-state oscillatory activity in NF1 and found an increase in resting-state slow wave power in the theta and delta bands. Although the mechanisms underlying increase in slow wave power are not understood, this is a characteristic of conditions impacting myelin such as multiple sclerosis. Animal studies suggest abnormal myelination in multiple sclerosis may promote a compensatory increase in slow wave power (Dubey et al., 2022). As myelin is also suggested to be impacted in NF1, a similar mechanism may be evident (de Blank et al., 2023). Increased resting theta power is also associated with worse cognitive function in the typically developing brain (Tan et al., 2024), consistent with the learning difficulties seen in NF1. A recent review suggests resting slow wave power differences are due to aperiodic activity (Tan et al., 2024), which is also affected in NF1 (Carter leno et al., 2022).

An interesting aspect noted across studies was that the age-associated developmental changes were delayed or atypical in the NF1 group (Jonas et al., 2017). Two studies used auditory habituation and change detection paradigms in infants and young children. Age-related changes were diminished in NF1 as compared to the control group (Begum-Ali et al., 2021). Similarly, Lalancette et al. (2023) found a larger increase in spectral power particularly in the theta band in the NF1 group in response to an auditory change detection task. This is suggestive of an immature response, as increased frontocentral power to change detection is often observed in infants (Deguire et al., 2022). Booth et al. (2023) found the age - Peak alpha frequency correlation disrupted in NF1 possibly suggesting developmental immaturity. The altered age - peak alpha frequency relationship has similarly been reported in other neurodevelopmental conditions like autism (Dickinson et al., 2018).

### 4.4 Frontal dysfunction as an important neural correlate of NF1 cognitive profile

The dorsolateral prefrontal cortex is central to working memory, cognitive flexibility, inhibitory control, decision making and attention (Badre & Wagner, 2004), which all underlie learning difficulties seen in NF1 (Hyman et al., 2005). Due to the prolonged maturation of this region throughout development (Kolk & Rakic, 2022), it is reasonable that disrupted neurobiological processes during development may have affected the function of the DLPFC, which is indeed affected in NF1. Mennigen et al. (2019) found resting-state connectivity within the medial frontal cortex was increased in NF1, even after controlling for possible covariates, with connectivity also reduced between frontal areas and the cerebellum. Similarly, both frontal-frontal and temporo-frontal resting-state FC was increased in NF1 in another study and associated with severity of cognitive deficits (Loitfelder et al., 2015), indicating a maladaptive role of the abnormal frontal network connectivity.

Task fMRI studies similarly show differences in frontal lobe activation during task performance Two studies found hypoactivation of the left and right dorsolateral prefrontal cortex during a visuospatial working memory task (Ibrahim et al., 2017; Shilyansky et al., 2010); right DLPFC hypoactivation also associated with worse task performance (Shilyansky et al., 2010). Importantly, as working memory load is increased, widespread, diffuse activation is observed which perhaps reflects less efficient neural functioning (Ibrahim et al., 2017). Furthermore, measurement of neurochemical concentrations in the prefrontal cortex can contribute to an understanding of its role as a hub in NF1. Ribeiro et al. (2015) found a reduction in baseline GABA levels in the medial frontal cortex, with higher GABA associated with increased IQ whereas this relationship was absent in controls. These results collectively indicating a shift in the functioning of the prefrontal cortex from that in the typically developing brain.

Disrupted frontal GABAergic signalling and thus, disrupted E/I balance essential for brain development may perturb the developmental trajectory of the DLPFC and explain the altered relationship between GABA levels and cognitive function as well as the atypical activation patterns observed. In addition, structural alterations in NF1 may all impact on the connectivity of the DLPFC with other regions, particularly the striatum and the thalamus (Violante et al., 2013b), critical for cognitive control (Jahfari et al., 2011). Further, previous research has reported alterations in the integrity of the white matter tracts connecting the thalamus to the frontal lobes (Karlsgodt et al., 2012). Collectively, disruption of this circuit may place an excessive reliance on the frontal lobes, contributing to the increased resting-state connectivity seen in the region. Likewise, during task engagement, prefrontal regions are less active due to their reduced capacity, and further regions are recruited beyond the typically engaged areas (Ibrahim et al., 2017), emphasising the dysconnectivity and network reorganisation that may have occurred. Ultimately, this would explain why the association of frontal brain metrics, such as prefrontal GABA, with cognitive function in NF1 is distinct from that of controls (Ribeiro et al., 2015). The prefrontal cortex being a hub for cognitive dysfunction has also been identified in other neurodevelopmental conditions such as schizophrenia (Smucny et al., 2022), ADHD (Liston et al., 2011) and autism (Li et al., 2024b; Schubert et al., 2015). Ultimately, this may be a broader finding across neurodevelopmental disorders whereby the pathogenic genetic variants result in delayed maturation of prefrontal areas, implicating white matter development, connectivity and synaptogenesis (Schubert et al., 2015).

### 4.5 Recommendations/implications for further research

This systematic review examined the neural underpinnings of cognitive difficulties seen in NF1. The single-gene aetiology of NF1 makes it an ideal syndromic model for understanding neurobiological underpinnings of conditions with a polygenic risk, such as ADHD (Lidzba et al., 2012) and autism (Garg et al., 2013). Future work needs to consider employing longitudinal study designs to examine the developmental brain changes in NF1 to understand how cognitive processes and underlying brain mechanisms evolve over time. Development of neuroimaging methods in clinical cohorts, analogous to measurements in animal models are also essential for translation of preclinical findings. Furthermore, environmental factors such as maternal education and whether the NF1 variant has been inherited or *de novo,* appear to be an important factor influencing the NF1 phenotype and ultimately the underlying neural mechanisms (Biotteau et al., 2020; Geoffray et al., 2021). Higher socioeconomic status (SES) and having a parent with NF1 are associated with higher IQ, better social skills and fewer symptoms of ADHD (Biotteau et al., 2020; Geoffray et al., 2021). Other factors, such as type of mutation, also influence symptoms. Ottenhoff et al. (2020) found a microdeletion was associated with significantly lower IQ as compared to intragenic variants, although there was no difference within different kinds of intragenic variants.

Finally, it must be remembered that reductionist approaches common in biomedicine are not sufficient to explain the cognitive/behavioural phenotypes in NF1. A systems neuroscience approach to study development at multiple levels from genetic, cellular, brain systems, and behaviour to explain different phenotypic outcomes is important (Johnson et al. (2021).

## Conclusion

Functional neuroimaging studies were synthesised in this review to better understand the neurocognitive phenotype in NF1. Overall, we find widespread heterogeneity between studies but with mechanisms that broadly reflect dysregulated E/I balance, alterations in brain connectivity and neural activation patterns. Ultimately, the neuroconstructivist approach proposed by Karmiloff-Smith (2015) must be applied to understanding the neurocognitive phenotype in NF1 whereby the multidirectional interactions of the NF1 gene on the developing brain and the environment must be considered.

## Data Availability

All data produced in the present work are contained in the manuscript and supplemental files

## Acknowledgements

This research was supported by the NIHR Manchester Biomedical Research Centre (NIHR203308). The views expressed are those of the author(s) and not necessarily those of the NIHR or the Department of Health and Social Care. Anna Wild is funded by the Medical Research Council Doctoral Training Program.

