## Supplementary File 1 for "Examining the cognitive phenotype in Neurofibromatosis 1: a systematic review of functional imaging studies"

### Supplementary file 1: Search queries for each database

| PUBMED query | SCOPUS query | MEDLINE, EMBASE and PsycINFO databases via Ovid query |
| --- | --- | --- |
| **((("Neurofibromatosis Type 1"[Title/Abstract] OR "NF1"[Title/Abstract]) OR ("Neurofibromatosis 1"[Mesh])) AND ((((((((((((((((((("Magnetic Resonance Imaging"[Title/Abstract]) OR (fMRI[Title/Abstract])) OR (connectivity[Title/Abstract])) OR (electroencephalography[Title/Abstract])) OR (EEG[Title/Abstract])) OR (ERP[Title/Abstract])) OR (MEG[Title/Abstract])) OR (magnetoencephalography[Title/Abstract])) OR ("Transcranial Magnetic Stimulation"[Title/Abstract])) OR (TMS[Title/Abstract])) OR ("Positron Emission Tomography"[Title/Abstract])) OR (PET[Title/Abstract])) OR (fNIRS[Title/Abstract])) OR ("Arterial Spin Labeling"[Title/Abstract])) OR (perfusion[Title/Abstract])) OR (ASL[Title/Abstract])) OR (Spectroscopy[Title/Abstract])) OR (MRS[Title/Abstract])) OR ((((((("Magnetic Resonance Imaging"[Mesh]) OR ( "Electroencephalography"[Mesh] OR "Neurofeedback"[Mesh] OR "Electrocorticography"[Mesh] OR "Electroencephalography Phase Synchronization"[Mesh] )) AND "Magnetoencephalography"[Mesh]) OR "Transcranial Magnetic Stimulation"[Mesh]) OR ( "Positron-Emission Tomography"[Mesh] OR "Positron Emission Tomography Computed Tomography"[Mesh] )) OR "Perfusion Imaging"[Mesh]) OR ( "Spectroscopy, Near-Infrared"[Mesh] OR "Magnetic Resonance Spectroscopy"[Mesh] OR "Nuclear Magnetic Resonance, Biomolecular"[Mesh] OR "Spectrum Analysis"[Mesh] OR "Proton Magnetic Resonance Spectroscopy"[Mesh] )))) AND (((("attention"[Title/Abstract] OR "cogniti*"[Title/Abstract] OR "function"[Title/Abstract] OR "adhd"[Title/Abstract] OR "asd"[Title/Abstract] OR "behav*"[Title/Abstract]) OR "performance"[Title/Abstract] OR ((((("Autism Spectrum Disorder"[Mesh]) OR "Autistic Disorder"[Mesh]) OR "Attention Deficit Disorder with Hyperactivity"[Mesh]) OR ( "Cognition"[Mesh] OR "Social Cognition"[Mesh] )) OR "Behavior"[Mesh]) AND (2000/1/1:2022/3/30[pdat]) AND (english[Filter]))))** | **( TITLE-ABS-KEY ( "attention" ) OR TITLE-ABS-KEY ( cogniti* ) OR TITLE-ABS-KEY ( performance) OR TITLE-ABS-KEY ( function ) OR TITLE-ABS-KEY ( adhd ) OR TITLE-ABS-KEY ( asd ) OR TITLE-ABS-KEY ( behav* ) ) AND ( TITLE-ABS-KEY-AUTH ( "Magnetic Resonance Imaging" ) OR TITLE-ABS-KEY ( fmri ) OR TITLE-ABS-KEY ( connectivity ) OR TITLE-ABS-KEY ( electroencephalography ) OR TITLE-ABS-KEY ( eeg ) OR TITLE-ABS-KEY ( erp ) OR TITLE-ABS-KEY ( meg ) OR TITLE-ABS-KEY ( "Transcranial Magnetic Stimulation" ) OR TITLE-ABS-KEY ( tms ) OR TITLE-ABS-KEY ( "Positron Emission Tomography" ) OR TITLE-ABS-KEY ( pet ) OR TITLE-ABS-KEY ( fnirs ) OR TITLE-ABS-KEY ( "Arterial Spin Labeling" ) OR TITLE-ABS-KEY ( asl ) OR TITLE-ABS-KEY ( perfusion ) OR TITLE-ABS-KEY ( spectroscopy ) OR TITLE-ABS-KEY ( mrs ) ) AND ( TITLE-ABS-KEY ( "Neurofibromatosis Type 1" ) OR TITLE-ABS-KEY ( nf1 ) PUBYEAR > 2000 ) AND ( LIMIT-TO ( LANGUAGE,"English" ) )** | **1 NF1.mh. or NF1.ti. or NF1.ab. or NF1.kw.**  **2 limit 1 to english language**  **3 Neurofibromatosis Type1.kw. or "Neurofibromatosis Type1".mh. or "Neurofibromatosis Type1".ti. or "Neurofibromatosis Type1".ab.**  **4 1 or 3**  **5 fmri.kw. or fmri.mh. or fmri.ti. or fmri.ab.**  **6 Magnetic Resonance Imaging.kw. or "Magnetic Resonance Imaging".mh. or "Magnetic Resonance Imaging".ti. or "Magnetic Resonance Imaging".ab.**  **7 connectivity.kw. or connectivity.mh. or connectivity.ti. or connectivity.ab.**  **8 electroencephalography.kw. or electroencephalography.mh. or electroencephalography.ti. or electroencephalography.ab.**  **9 eeg.kw. or eeg.mh. or eeg.ti. or eeg.ab.**  **10 erp.kw. or erp.mh. or erp.ti. or erp.ab.**  **11 meg.kw. or meg.mh. or meg.ti. or meg.ab.**  **12 Transcranial Magnetic Stimulation.kw. or "Transcranial Magnetic Stimulation".mh. or "Transcranial Magnetic Stimulation".ti. or "Transcranial Magnetic Stimulation".ab.**  **13 tms.kw. or tms.mh. or tms.ti. or tms.ab.**  **14 Positron Emission Tomography.kw. or "Positron Emission Tomography".mh. or "Positron Emission Tomography".ti. or "Positron Emission Tomography".ab.**  **15 pet.kw. or pet.mh. or pet.ti. or pet.ab.**  **16 fnirs.kw. or fnirs.mh. or fnirs.ti. or fnirs.ab.**  **17 Arterial Spin Labeling.kw. or "Arterial Spin Labeling".mh. or "Arterial Spin Labeling".ti. or "Arterial Spin Labeling".ab.**  **18 asl.kw. or asl.mh. or asl.ti. or asl.ab.**  **19 perfusion.kw. or perfusion.mh. or perfusion.ti. or perfusion.ab.**  **20 spectroscopy.kw. or spectroscopy.mh. or spectroscopy.ti. or spectroscopy.ab.**  **21 mrs.kw. or mrs.mh. or mrs.ti. or mrs.ab.**  **22 5 or 6 or 7 or 8 or 9 or 10 or 11 or 12 or 13 or 14 or 15 or 16 or 17 or 18 or 19 or 20 or 21**  **23 attention.kw. or attention.mh. or attention.ti. or attention.ab.**  **24 cogniti*.kw. or cogniti*.mh. or cogniti*.ti. or cogniti*.ab. or behav*.kw. or behav*.mh. or behav*.ti. or behav*.ab.**  **25 function.kw. or function.mh. or function.ti. or function.ab. or performance.ab.**  **26 adhd.kw. or adhd.mh. or adhd.ti. or adhd.ab.**  **27 asd.kw. or asd.mh. or asd.ti. or asd.ab.**  **28 23 or 24 or 25 or 26 or 27**  **29 4 and 22 and 28**  **30 limit 29 to yr="2000 -Current"** |
