## Supplementary File 2 for "Examining the cognitive phenotype in Neurofibromatosis 1: a systematic review of functional imaging studies"

**Title**

Title of paper / abstract / report that data are extracted from

**Lead author**

**Country in which the study conducted**

add if other

1. United States
2. UK
3. Canada
4. Australia
5. Other

**Characteristics of included studies**

Data to be collected based on:

Li, T., Higgins, J. P. T., & Deeks, J. J. (2024). Chapter 5: Collecting data. In J. P. T. Higgins, J. Thomas, J. Chandler, M. Cumpston, T. Li, M. J. Page, & V. A. Welch (Eds.), *Cochrane Handbook for Systematic Reviews of Interventions version 6.5*. <www.training.cochrane.org/handbook>.

**Methods**

**Study design**

1. Randomised controlled trial
2. Non-randomised experimental study
3. Cohort study
4. Cross sectional study
5. Other

**2 a) NF1 Diagnostic criteria _ Inclusion Criteria**

** if children specify

1. NIH criteria
2. Aged 18 and above
3. Parent report of NF1
4. Clinical diagnosis
5. Genetic/DNA testing
6. Parents and/or participants fluent in English
7. Not reported
8. Unclear
9. Other

**2 b) Inclusion criteria Controls**

copy paste

i.e. Selection criteria (Parents and/or participants fluent in English), no known medical or de- velopmental condition

**3 b) Exclusion criteria Controls**

copy paste - low SNR, excessive movement, etc.

**3 a) Exclusion Criteria NF1**

* ID (Intelectual disability), SNR (Signal-to-noise ratio)

1. FSIQ > 70
2. IQ > 75
3. No history of ID/ or ID
4. No significant CNS pathology
5. Aged 18 years and above
6. No concurrent genetic disorder
7. No significant hearing or visual problems
8. No significant visual problems
9. No co-existing genetic or neurological disorder
10. No ADHD diagnosis
11. No learning disabilities diagnosis
12. No psychiatric disorder
13. No epilepsy or neuro
14. No neurologic disorder
15. No significant intracranial abnormality
16. Not taking neurotropic medication or simvastatin
17. Not taking anxiety or depression medication
18. No excessive movement
19. No low SNR
20. Other

**Method of recruitment of participants**

Copy paste text if needed

Setting/ Region(s) and country/countries from which study participants were recruited

i.e. which centre? Multicentre (& how many)?

1. Phone
2. Mail
3. Clinic patients
4. Voluntary
5. Other

**4 Participant testing**

1. Random sequence generation
2. Allocation sequence concealment
3. Masking for RCT
4. Used randomized or interleaved participant scanning order (to avoid bias due to scanner changes/upgrades)
5. Other

**5 Participants**

Recruited participants and number of successful scans/tests may be different. Add in brackets the final sample tested, before individual data preprocessing. The final sample used in statistical analysis reflects the total number of participants retained after preprocessing/motion correction etc.

If missing info: enter N/A. If just total number (not NF1/control) is mentioned in text, note that in final column.

|  | **Contacted** | **Recruited (tested/scanned)** | **Tested** | **Excluded after acquisition** | **Included in statistical analysis** | **age (range, mean)** | **gender (M/F, absolute counts or relative frequencies)** | **Handedness (just type Y if reported, or NR for not reported)** |
| --- | --- | --- | --- | --- | --- | --- | --- | --- |
| **Control** |  |  |  |  |  |  |  |  |
| **NF1** |  |  |  |  |  |  |  |  |
| **NF1 post intervention** |  |  |  |  |  |  |  |  |
| **Total** |  |  |  |  |  |  |  |  |

**6 a) Participants Matched for**

note if they compared groups for age, sex, IQ etc.

1. individually matched for age
2. individually matched for sex
3. individually matched for IQ
4. group matched for age
5. group matched for sex
6. group matched for IQ
7. Other

**Intervention**

**Length of intervention - Frequency**

Copy paste (i.e. total scan time)

**Imaging, Neurophysiological or brain stimulation methods**

1. MRI
2. RS fMRI
3. task based fMRI
4. EEG
5. MEG
6. TMS
7. PET
8. fNIRS
9. ASL
10. MRS
11. Other

**significant differences in motion between groups**

1. not reported
2. reported (summaries of subject motion, poor SNR etc were reported in text)
3. not tested
4. participants matched for head motion
5. matched for scan parameters
6. Other

**fMRI - ASL - RS fMRI**

**Participant Preparation**

**7 Mock scanning, special accomodations and experimenter personnel**

Check only if it applies

Checklist Based on OHBM COBIDAS Report and Poldrack et al., 2008

1. a Study clearly reports the protocol used for mock scanning (duration, types of simulated scans and or experiments)
2. b Study reported clearly an acclimatization procedure without the use of a mock scanner
3. c Study reported clearly any special accommodations (i.e. presence of parent/guardian in the room for pediatric scanning)
4. d If participants required to undergo deep sedation or general anesthesia, the process was described clearly and in detail (if natural sleep techniques were used, or only part of the sample underwent the aforementioned processed, the report clearly stated the number/percentage of awake participants)
5. e Study did not report clearly the sedation and or preparation protocol used for scanning
6. f Study reported using the same sedation and or preparation protocol for scanning controls and cases
7. Other

**MRI system description & acquisition**

**8 MRI system description & acquisition reporting**

Check only if it applies

Checklist Based on OHBM COBIDAS Report and Poldrack et al., 2008

For ASL:

ASL Labelling method (e.g. continuous ASL (CASL), pseudo?continuous ASL (PCASL), Pulsed ALS (PASL), velocity selective ASL (VSASL)).

● Use of background suppression pulses and their timing.

● For either PCASL or CASL report: ○ Label Duration. ○ Postlabeling delay (PLD). ○ Location of the labeling plane.

● For PCASL also report: ○ Average labeling gradient. ○ Slice-selective labeling gradient. ○ Flip angle of B1 pulses. ○ Assessment of inversion efficiency; QC used to ensure off-resonance artifacts not problematic, signal obtained over whole brain.

● For CASL also report:○ Use of a separate labeling coil. ○ Control scan/pulse used. ○ B1 amplitude.

● For PASL report ○ TI. ○ Labeling slab thickness. ○ Use of QUIPSS pulses and their timing.

● For VSASL ○ TI. ○ Choice of velocity selection cutoff (“VENC”).

1. Model, make and field strength in tesla was stated
2. Receive and transmit coil, and gradient system were sufficiently described
3. Pulse sequence type (i.e. spin echo), imaging type (i.e. EPI, spiral, 3D), number of shots (if multi-shot) and partial Fourier scheme & reconstruction method (if used) were clearly described
4. Essential sequence & imaging parameters were stated (i.e. echo time, repetition time, flip angle, duration, number of volumes per session)
5. Imaging and phase encoding parameters were stated (i.e. field of view, in-plane matrix size, slice thickness/orientation, interslice skip,3D matrix size)
6. Reported parallel imaging method & parameters (e.g. SENSE, GRAPPA or other parallel imaging method, and acceleration factor, Matrix coil mode, and coil combining method (if non­standard)
7. Reported weather fat suppression was used or not for the anatomical scans
8. Reported using specialized shimming procedures
9. Reported slice order and timing (i.e. interleaved vs sequential ordering and direction - ascending/descending-, location of 1st slice)
10. Reported whether coverage was whole-brain, and whether cerebellum and brainstem were included ( If not, noted the nature of the partial area of coverage. If axial and co­planar with AC­PC line, the volume coverage in terms of Z in mm)
11. The scanner-side preprocessing was clearly described (i.e. Reconstruction matrix size differing from acquisition matrix size, Prospective-motion correction process, Signal inhomogeneity correction, Distortion-correction)
12. Number of initial "dummy" scans acquired and then discarded by scanner was stated (T1 stabilization)
13. ASL Labeling method was clearly described
14. Other

**9 Preliminary Quality Control**

Checklist Based on OHBM COBIDAS Report and Poldrack et al., 2008

1. Reported performing visual or quantitative checks for severe motion

**10 Design Specification**

Check only if it applies

Checklist Based on OHBM COBIDAS Report and Poldrack et al., 2008

1. There was a clear description of conditions and or stimuli used (i.e. baseline may be a blank white/black screen, or a fixation cross)
2. The number of blocks, trials or experimental units was clearly described (specified per session, and if differing by subject, summary statistics of such counts)
3. The length of each trial or block and interval between trials was reported
4. The total length of the scanning session was reported, as well as the duration of each run, and or trial and or block, and interval in between them.
5. Study reports whether and how the design was optimized for efficiency
6. The presentation software was described clearly (name, version, operating system or code used to drive the experiment)
7. Other

**11 Task specification**

Check only if it applies

Checklist Based on OHBM COBIDAS Report and Poldrack et al., 2008

1. Conditions were clearly described (psychological construct and specific modeled effect are clearly described)
2. Instructions to participants were clearly described (i.e. for resting-state they indicated whether it was eyes-closed, eyes-open, any fixation. )
3. Stimuli were clearly described for each run (the unique number of used stimuli was reported, in addition to whether/how stimuli were repeated over trials or conditions)
4. Randomization procedures were appropriate and clearly described (block or event ordering was described as deterministic, or reported manner of randomization in terms of order and timing. if pseudo-randomized, the criteria used to constrain the orders/timing are described)
5. The presentation and response collection hardware were clearly described, in addition to how it was synced to the scanner (e.g. scanner TTL, or manual sync.)
6. The order in which tasks runs were conducted in the scanner was stated
7. Other

**12 Power analysis**

Check only if it applies

Checklist Based on OHBM COBIDAS Report and Poldrack et al., 2008

1. The report specified the type of outcome used as basis of power computations (e.g. signal in a pre-specified ROI, or whole image voxelwise, or cluster-wise, or peak-wise)
2. The power parameters were clearly described (e.g. effect size or magnitude and SD, source of predicted effect size - previous literature citation-, significance level, target power - typically 80%- etc.)
3. Other

**13 Behavioural Performance**

Check only if it applies

Checklist Based on OHBM COBIDAS Report and Poldrack et al., 2008

1. The number of variables recorded was stated (e.g. correct button press, RT)
2. Summaries of behavior sufficient to establish that the subjects were performing the task as expected were provided (i.e. mean, range, SD of correct response rates and/or RTs summarized over subjects)
3. Other

**Data Preprocessing**

copy paste i.e. software used, template for registration, smoothing, parcellation ; steps: artifact correction process, bandpass filter cutoff frequencies/epoch rejection threshold, or movement defined in mm translation/rotation AND number of trials per condition left OR time-series, time-points left per subject

Checklist Based on OHBM COBIDAS Report and Poldrack et al., 2008

**14 Preprocessing reporting**

Check only if it applies

If slice time correction performed, report: ● Name of software/method. ● Whether performed after or before motion correction. ● Reference slice. ● Interpolation type and order (e.g., 3 rd

order spline or sinc)

If motion correction performed, :

Report: ● Name of software/method. ● Use of non?rigid registration, and if so the type of transformation. ● Use of motion susceptibility correction (fieldmap-based unwarping), as well as the particular software/method. ● Reference scan (e.g. 1 st

scan or middle scan).

● Image similarity metric (e.g. normalized correlation, mutual information, etc).● Interpolation type (e.g., spline, sinc), and whether image transformations are combined to allow a single interpolation.

● Use of any slice-to-volume registration methods, or integrated with slice time correction.

For the modelling/post-processing scheme ASL, Report: ● For subtraction, specify whether simple subtraction, running, sinc-subtraction, etc.

● For quantitative model, specify model used, number of free parameters.

Checklist Based on OHBM COBIDAS Report and Poldrack et al., 2008

1. Software used, and version & revision number was stated
2. Number of initial “dummy” scans discarded as part of preprocessing was stated (if not already performed by scanner)
3. Brain extraction was clearly described (i.e. name of software/method, parameter choices, and/or manual editing applied to masks)
4. Name or method of motion correction was clearly described
5. Slice time correction was clearly described
6. For structural images, the method used to extract tissue classes was described
7. Gradient distortion correction was reported
8. The modelling/post-processing scheme was clearly described
9. Other

**15 Intra and intersubject registration**

Check only if it applies

For intra-subject function-structure coregistration, Report:

Report: ● Name of software/method (e.g., FSL flirt followed by fnirt, FreeSurfer, Caret, Workbench, etc)

● Whether volume and/or surface based registration is used (if not already clearly implied).

● Image types registered (e.g. T2* or T1). ● Any preprocessing to images; e.g. for T1, bias field correction, or segmentation of gray matter; for T2*, single image (specify image) or mean image.● Interpolation type (e.g., spline, linear); if projection from volume to surface space, how were voxels sampled from the volume (e.g., trilinear; nearest neighbor; ribbon?constrained specifying inner and outer surface used).

● Cost function (e.g., correlation ratio, mutual information, SSD). ● Use of cost?function masking.

Checklist Based on OHBM COBIDAS Report and Poldrack et al., 2008

1. Name or method used for function-structure coregistration was clearly described(intra-subject)
2. Type of transformation (rigid, nonlinear); if nonlinear, type of transformation was stated (intra-subject) and Cost function (e.g., correlation ratio, mutual information, boundary­-based registration, etc) and Interpolation method (e.g., spline, linear) were stated (intra-subject)
3. Name or method used for inter-subject registration was clearly described (Transformation model (linear/affine, nonlinear), type of any non-linear transformations (polynomial, discrete cosine basis), number of parameters (e.g., 12 parameter affine, 3×2×3 DCT basis), regularization, image-similarity metric, and interpolation method)
4. Image types registered (e.g. T2* or T1) and any preprocessing to images was stated (e.g. for T1, bias field correction, or segmentation of gray matter; for T2*, single image (specify image) or mean image)
5. Template space (e.g., MNI, Talairach, fsaverage, FS_LR), modality (e.g., T1, T2*), resolution (e.g., 2mm, fsaverage5, 32k_FS_LR), and the specific name of template image used were clearly stated
6. Intensity correction was clearly stated (Bias field corrections for structural MRI, but also correction of odd versus even slice intensity differences attributable to interleaved EPI acquisition without gaps)
7. Other

**16 Spatial smoothing**

Check only if it applies

Checklist Based on OHBM COBIDAS Report and Poldrack et al., 2008

1. If spatial smoothing smoothing was performed, the study stated the following: 1) Name of software/method, 2) Size & type of kernel, 3) Filtering approach, e.g., fixed kernel or iterative smoothing until fixed FWHM, 4) Space in which smoothing is performed (i.e. native volume, native surface, MNI volume, template surface)
2. Other

**17 Artifact and structured noise removal**

Check only if it applies

● Interpolation type (e.g., spline, linear); if projection from volume to surface space, how were voxels sampled from the volume (e.g., trilinear; nearest neighbor; ribbon?constrained specifying inner and outer surface used).

● Cost function (e.g., correlation ratio, mutual information, SSD). ● Use of cost?function masking.

Bias field corrections for structural MRI, but also correction of odd versus even slice intensity differences attributable to interleaved EPI acquisition without gaps.

Scan?by?scan or run?wide scaling of image intensities before statistical modelling. E.g. SPM scales each run such that the mean image will have mean intracerebral intensity of 100; FSL scales each run such that the mean image will have an intracerebral mode of 10,000.

Use of physiological noise correction method. Report: ● Name of software/method used (e.g. CompCor, ICA?FIX, ICA?AROMA, etc.).

● If using a nuisance regression method, specify regressors used; for each type, include key details, as follows: ○ Motion parameters. ■ Expansion basis and order (e.g. 1st temporal derivatives; Volterra kernel expansion)

○ Tissue signals. ■ Tissue type (e.g., whole brain, gray matter, white matter, ventricles).

■ Tissue definition (e.g., a priori seed, automatic segmentation, spatial regression).

■ Signal definition (e.g., mean of voxels, first singular vector, etc.).

○ Physiological signals ■ e.g., heart rate variability, respiration ■ Modeling choices (e.g. RETROICOR, cardiac and/or respiratory response functions) and number of computed regressors.

For “scrubbing” or “despiking”. Report: ● Name of software/method. ● Criteria (e.g., frame-by-frame displacement threshold, percentage BOLD change).

● Use of censoring or interpolation; if interpolation, method used (e.g., spline, spectral estimation).

Checklist Based on OHBM COBIDAS Report and Poldrack et al., 2008

1. Name of software/method used for noise correction was stated
2. For nuisance regression method, the report specified regressors used and key details (i.e. for motion parameters: expansion basis and order; for tissue signals: the type, and definition method; for physiological signals: the modeling choices and number of computed regressors)
3. For “scrubbing” or “de­spiking”, the report clearly stated the name of software/method, the criteria (e.g., frame-by-frame displacement threshold, percentage BOLD change), and the use of censoring or interpolation; if interpolation, method used (e.g., spline, spectral estimation)
4. Other

**18 Resting state fMRI: Creation of summary measure like ALFF, fALFF, ReHo**

Check only if it applies

For Creation of summary measure like ALFF, fALFF, ReHo.

For ALFF, fALFF report: ● Lower and upper band pass frequencies.

For ReHo, report: ● Neighborhood size used to compute local similarity measures (e.g. 6, 18 or 26).

● Similarity measure (e.g. Kendall’s coefficient of concordance).

Checklist Based on OHBM COBIDAS Report and Poldrack et al., 2008

1. For ALFF, fALFF reported lower and upper band pass frequencies
2. For ReHo, reported neighborhood size used to compute local similarity measures (e.g. 6, 18 or 26) and similarity measure (e.g. Kendall’s coefficient of concordance)
3. Other

**Statistical modeling & inference, Results**

**19 a) Mass univariate analyses**

Check only if it applies

see OHBM COBIDAS report for more details

1. Reported the number of time points, number of subjects; specify exclusions of time points / subjects, if not already specified in experimental design
2. If not “Full brain”, the study specified an anatomically or functionally defined mask
3. The independent variables, model type and settings were clearly described
4. The contrast/effect and search region was clearly defined and described
5. The statistic type was clearly described (i.e. voxel-wise, or cluster size, mass, Threshold-free Cluster Enhancement, cluster-forming threshold etc)
6. The multiple testing correction was clearly described
7. The drift, movement or any other nuisance regressors were clearly described (i.e. whether they were entered as interactions, whether or not covariates are split by group)
8. The report clearly stated if the search region was whole brain or “small volume” for each contrast
9. Provided a complete list of tested and omitted effects
10. The extracted data, tables of coordinates, thresholded/unthresholded maps were described well
11. Other

**19 b) Functional connectivity**

Check only if it applies

see OHBM COBIDAS report for more details

for Multivariate method: Independent Component Analysis, Report:● Algorithm to estimate components. ● Number of components (if fixed), or algorithm for estimating number of components.

● If used, method to synthesize multiple runs. ● Sorting method of IC’s, if any. ● Detailed description of how components were chosen for further analysis

Dependent variable definition: For seed?based analyses report: ● Definition of the seed region(s). ● Rationale for choosing these regions.

For region?based analyses report: ● Number of ROIs. ● How the ROI’s are defined (e.g. citable anatomical atlas; auxiliary fMRI experiments); note if ROIs overlap.

● Assignment of signals to regions (i.e. how a time series is obtained from each region, e.g. averaging or first singular vector) ● Note if considering only bilateral (L+R) merged regions

Functional connectivity measure/ modelReport: ● Measure of dependence used, e.g. Pearson’s (full) correlation, partial correlation, mutual information, etc; also specify: ○ Use of Fisher’s Z-transform (Yes/No) and, if standardised, effective N is used to compute standard error (to account for any filtering operations on the data).

○ Estimator used for partial correlation. ○ Estimator used for mutual information.

● Regression model used to remove confounding effects (Pearson or partial correlation)

Effectivity connectivity Report: ● Model. ● Algorithm used to fit model. ● If per?subject model, method used to generalize inferences to population. ● Itemize models considered, and method used for model comparison

Graph analysis

Report the ‘dependent variable’ and ‘functional connectivity measure’ used (see above).

Specify either: ● Weighted graph analysis or, ● Binarized graph analysis is used, clarifying the method used for thresholding (e.g. a 10% density threshold, or a statistically?defined threshold); consider the sensitivity of your findings to the particular choice of threshold used.

Itemise the graph summaries used (e.g. clustering coefficient, efficiency, etc), whether these are global or per-node/per-edge summaries. In particular with fMRI or EEG, clarify if measures applied to individual subject networks or group networks

1. Method for detecting movement artifacts, movement-related variation, and remediation (e.g. ‘scrubbing’, ‘despiking’, etc) was described clearly
2. Report described the use of global signal regression, exact type of global signal used and how it was computed
3. Report described whether a high or low-pass temporal filtering is applied to data, and at which point in the analysis pipeline (Note, any temporal regression model using filtered data should have it’s regressors likewise filtered)
4. Method used for functional connectivity was described well and in a good level of detail (i.e. independent component analysis, dependent variable definition, functional connectivity model, effective connectivity, and graph analysis)
5. Other

**19 c) Multivariate modelling & predictive analysis**

Check only if it applies

see OHBM COBIDAS report for more details

For traditional multivariate analyses, report: ● Type of model, e.g. MANOVA. ● Assumptions made on the covariance structure, e.g. independence, or a common arbitrary covariance between groups.

● Statistic used to assess significance, e.g. Wilk’s lambda, Hotelling?Lawley trace, etc.

For predictive models, report: ● Type of model, e.g. Linear discriminant analysis, support vector machines, logistic regression, etc.

● For kernel?based methods (i.e. SVM) report type of kernel used, type and number of parameters needed to be estimated.

For learning method Report:

● Figure-of-merit optimised. ● Fitting method. ● Parameter settings, those fixed and those estimated; specify how fixed parameter values were chosen.

● How the convergence of the learning method is monitored.

For training procedure, Describe: ● Pipeline structure applied uniformly to all cases (e.g. that could be independently applied to a new case).

● Method for hyperparameter setting. ● Data splitting (cross validation)

Evaluation metrics:

Discrete response: ● Accuracy. ● If group sizes unequal, balanced (or average) accuracy.

When there are only 2 classes, and one can be labeled “positive”: ● Precision (1 − false discovery rate). ● Recall (sensitivity). ● False positive rate (1?specificity). ● F1 (incorporates both precision and recall). ● Receiver operating characteristic (ROC) curves, e.g. summarised by area under the curve (AUC); AUC for only high specificity (e.g. false positive rates no greater than 10%) are also useful.

When there are 3 or more classes: ● Report the confusion matrix

Continuous response

Evaluation, Report “Prediction R 2 ”, the percentage of variance explained by prediction, computed as

one minus the ratio of prediction sum-of-squares to total sum-of-squares. (Note this is not the squared correlation coefficient between true and predicted values).

Representational similarity analysis: Report the Kendall Tau statistic for each candidate model considered.

1. Method used for Multivariate modelling & predictive analysis was described well and in a good level of detail (i.e. defined model and independent variables)
2. Features extraction and dimension reduction was described well
3. The learning method was described well
4. The training procedure was described well
5. The evaluation metrics computed were described well
6. When possible, they used formal test to obtain P value to assess whether evaluation metric is “significant” or consistent with noise

**MRS**

**Hardware**

**1 Hardware**

check only if it applies

for details see Lin at al., 2021

1. Model, make and field strength in tesla was stated
2. RF coils: nuclei (transmit/receive), number of channels and type were stated
3. Other

**2 a) Acquisition**

check only if it applies

for details see Lin at al., 2021

e. Total number of excitations or acquisitions per spectrum

In time series for kinetic studies i. Number of averaged spectra (NA) per time point ii. Averaging method (eg block-wise or moving average)

iii. Total number of spectra (acquired/in time series)

f. Additional sequence parameters (spectral width in Hz, number of spectral points, frequency offsets)

If STEAM:, mixing time (TM) If MRSI: 2D or 3D, FOV in all directions, matrix size, acceleration factors, sampling method

1. Pulse sequence (TR, TE) , volume of interest and size were clearly described
2. Total number of excitations or acquisitions per spectrum was clearly described
3. Additional sequence parameters (spectral width in Hz, number of spectral points, frequency offsets) were clearly described
4. Water suppression was clearly described (and any other suppression methods used, eg lipid suppression, outer volume suppression)
5. Shimming method, reference peak, and thresholds for “acceptance of shim” chosen were clearly described
6. Other

**2 b) Triggering and or motion correction method (respiratory, peripheral, cardiac triggering, incl. device used and delays was clearly described**

check only if it applies

for details see Lin at al., 2021

1. Yes
2. No
3. Unclear
4. Not reported
5. Other

**3 Data analysis methods and outputs**

check only if it applies

for details see Lin at al., 2021

1. Analysis software and processing steps deviating from quoted reference or product were clearly described
2. Output measure (eg absolute concentration, institutional units, ratio), processing steps deviating from quoted reference or product were clearly described
3. Quantification references and model, fitting model assumptions were clearly described
4. Other

**4 Data quality**

check only if it applies

for details see Lin at al., 2021

1. a Reported variables (SNR, linewidth (with reference peaks), sample spectrum and data exclusion criteria were clearly described
2. b Quality measures of post processing model fitting (eg CRLB, goodness of fit, SD of residual) were clearly described
3. c Visual inspection of the MR spectrum was performed
4. Other

**TMS**

**Checklist**

check only if it applies

see Chipchase et al., 2012 for details

1. 1 Position and contact of EMG electrodes was clearly described
2. 2 Amount of relaxation/contraction of target muscles was clearly described
3. 3 Prior motor activity of the muscle to be tested was clearly described
4. 4 Level of relaxation of muscles other than those being tested was stated
5. 5 Coil type (size and geometry), location and stability (with or without a neuronavigation system)and orientation were clearly described
6. 6 Direction of induced current in the brain was clearly described
7. 7 Type of stimulator used (e.g. brand) and intensity were clearly described
8. 8 Pulse shape (monophasic or biphasic) was clearly described
9. 9 Determination of optimal hotspot was clearly described
10. 10 The time between MEP trials and or between days of testing were clearly described
11. 11 Subject attention (level of arousal) during testing was evaluated
12. 12 Method for determining threshold (active/resting) was clearly described
13. 13 Number of MEP measures made was stated
14. 14 Paired pulse only: Intensity of test pulse, intensity of conditioning pulse, and Inter-stimulus interval were described
15. 15 Method for determining MEP size during analysis was clearly described
16. 16 Size of unconditioned MEP was stated
17. Other

**EEG**

**1 Recording characteristics and instruments**

check only if it applies

see Keil et al. 2014, for details

1. The type of EEG/MEG sensor was described, including make and model All
2. All sensor locations were specified, including reference electrode(s) for EEG
3. Sampling rate was indicated
4. Online filters were clearly described, specifying the type of filter and including roll-off and cut-off parameters (in dB, or by indicating whether the cut-off represents half-power/half amplitude)
5. Amplifier characteristics were described
6. Electrode impedance or similar information was provided

**2 Stimulus and timing parameters**

check only if it applies

see Keil et al. 2014, for details

1. Timing of all stimuli, responses, intertrial intervals, etc., were fully specified (ensure clarity that intervals were from onset or offset)
2. Characteristics of the stimuli were described such that replication is possible
3. Other

**3 Description of data preprocessing steps**

check only if it applies

see Keil et al. 2014, for details

1. The order of all data preprocessing steps was included
2. Referencing procedures (if any) were specified, including the location of all sensors contributing to the new reference
3. Method of interpolation (if any) was clearly described
4. Segmentation procedures were clearly described, including epoch length and baseline removal time period
5. Offline filters were clearly described (specifying the type of filter and including roll-off and cut-off parameters (in dB, or by indicating whether the cut-off represents half-power/half amplitude)
6. The number of trials used for averaging (if any) was clearly described, reporting the number of trials in each condition and each group of subjects (this should include both the mean number of trials in each cell and the range of trials included)
7. Other

**4 Artifact rejection and correction procedures**

check only if it applies

see Keil et al. 2014, for details

1. Artifact rejection procedures were clearly described, including the type and proportion of artifacts rejected
2. Artifact correction procedures were clearly described, including the procedure used to identify artifacts, the number of components removed, and whether they were performed on all subjects
3. Other

**5 Measurement procedures**

check only if it applies

see Keil et al. 2014, for details

1. Measurement procedures were clearly described, including the measurement technique (e.g., mean amplitude), the time window and baseline period, sensor sites, etc.
2. For peak amplitude measures, the following was included: whether the peak was an absolute or local peak, whether visual inspection or automatic detection was used, and the number of trials contributing to the averages used for measurement
3. An a priori rationale was given for the selection of time windows, electrode sites, etc.
4. Both descriptive and inferential statistics were included
5. Other

**6 Statistical analyses**

check only if it applies

see Keil et al. 2014, for details

1. Appropriate correction for any violation of model assumptions was implemented and described (e.g., Greenhouse-Geisser, Huynh-Feldt, or similar adjustment)
2. The statistical model and procedures were clearly described and results were reported with test statistics, in addition to p value
3. An appropriate adjustment was performed for multiple comparisons
4. If permutation or similar techniques were applied, the number of permutations was indicated together with the method used to identify a threshold for significance
5. Data figures for all relevant comparisons were included and described in a good level of detail
6. Other

**7 Spectral analyses**

check only if it applies

see Keil et al. 2014, for details

1. The temporal length and type of data segments (single trials or averages) entering frequency analysis were clearly define
2. The decomposition method was described, and an algorithm or reference given
3. The frequency resolution (and the time resolution in time-frequency analyses) was specified
4. The use of any windowing function is described, and its parameters (e.g., window length and type) was given
5. The method for baseline adjustment or normalization was specified, including the temporal segments used for baseline estimation, and the resulting unit
6. Other

**8 Source-estimation procedures**

check only if it applies

see Keil et al. 2014, for details

1. The volume conductor model and the source model were fully described, including the number of tissues, the conductivity values of each tissue, the (starting) locations of sources, and how the sensor positions are registered to the head geometry
2. The source estimation algorithm was described, including all user-defined parameters (e.g., starting conditions, regularization)
3. Other

**9 Principle component analysis (PCA)**

check only if it applies

see Keil et al. 2014, for details

1. The structure of the EEG/MEG data submitted to PCA was fully described
2. The type of association matrix was specified
3. The PCA algorithm was described
4. Any rotation applied to the data was described
5. The decision rule for retaining/discarding PCA components was described
6. Other

**10 Independent component analysis (ICA) The**

check only if it applies

see Keil et al. 2014, for details

1. The structure of the EEG/MEG data submitted to ICA was described
2. The ICA algorithm was described
3. Preprocessing procedures, including filtering, detrending, artifact rejection, etc., were clearly described
4. The information used for component interpretation and clustering was described
5. The number of components removed (or retained) per subject was clearly described
6. Other

**11 Multimodal imaging Single-modality**

1. Single-modality results were reported
2. Other

**12 Current source density and Laplacian transformations**

1. The algorithm used and the interpolation functions were described

**13 Single-trial analyses**

1. All preprocessing steps were clearly described
2. A mathematical description of the algorithm is included or a reference to a complete description was provided

**Results**

**Correlations**

edit if possible

|  | **Model - Statistical Test** | **Variables** | **Positive/Negative** | **Covariates** | **Sig level (i.e. 0.01 or non-sig)** | **Brain region (s)** |
| --- | --- | --- | --- | --- | --- | --- |
| **1** |  |  |  |  |  |  |
| **2** |  |  |  |  |  |  |
| **3** |  |  |  |  |  |  |
| **4** |  |  |  |  |  |  |
| **5** |  |  |  |  |  |  |
| **6** |  |  |  |  |  |  |

**Comparisons**

|  | **Model - Statistical Test** | **Variables** | **Direction of effect (i.e. NF1> control or NF1 baseline > NF1 post-treatment)** | **Covariates** | **Sig level (i.e. 0.01 or non-sig)** | **Brain region (s)** |
| --- | --- | --- | --- | --- | --- | --- |
| **1** |  |  |  |  |  |  |
| **2** |  |  |  |  |  |  |
| **3** |  |  |  |  |  |  |
| **4** |  |  |  |  |  |  |
| **5** |  |  |  |  |  |  |
| **6** |  |  |  |  |  |  |

**Outcome table (measures)**

*** Higher indicates better performance unless otherwise stated

|  | **NF1 Mean (SD) / Control Mean (SD)** | **Sig or Non-sig (effect size, p value)** | **Rater (parent, self, researcher/clinician, teacher, computer)** | **Behavioural Tasks / Questionnaires / Clinical Assessments** |
| --- | --- | --- | --- | --- |

**Source(s) of funding or other material support for the study/ Authors’ financial relationship and other potential conflicts of interest**

Data to be collected based on

Cite this chapter as: Li T, Higgins JPT, Deeks JJ (editors). Chapter 5: Collecting data. In: Higgins JPT, Thomas J, Chandler J, Cumpston M, Li T, Page MJ, Welch VA (editors). Cochrane Handbook for Systematic Reviews of Interventions version 6.3 (updated February 2022). Cochrane, 2022. Available from www.training.cochrane.org/handbook.

1. No conflicts reported
2. Other

**The study addressed an appropriate and clearly focused question**

JBI Critical Appraisal Checklist for Case Control Studies - modified

1. Yes
2. No
3. Unclear
4. Not reported
5. Not applicable

**Were participants diagnosed as patients with disease of interest using the appropriate clinical testing criteria?**

JBI Critical Appraisal Checklist for Case Control Studies - modified

1. Yes
2. No
3. Unclear
4. Not reported
5. Not applicable

**Were the groups comparable other than the presence of disease in cases or the absence of disease in controls?**

* this is to note if they excluded for example individuals with a psychiatric disorder from NF1 group, but not controls etc.

JBI Critical Appraisal Checklist for Case Control Studies - modified

1. Yes
2. No
3. Unclear
4. Not reported
5. Not applicable

**Were cases and controls matched appropriately?**

JBI Critical Appraisal Checklist for Case Control Studies - modified

1. Yes
2. No
3. Unclear
4. Not reported
5. Not applicable

**Was the quantitative brain metric measured in the same way for cases and controls?**

JBI Critical Appraisal Checklist for Case Control Studies - modified

1. Yes
2. No
3. Unclear
4. Not reported
5. Not applicable

**Was the quantitative brain metric measured in a standard, valid and reliable way?**

JBI Critical Appraisal Checklist for Case Control Studies - modified

1. Yes
2. No
3. Unclear
4. Not reported
5. Not applicable

**Was the performance on the cognitive task or clinical assessment tool measured in the same way for cases and controls?**

JBI Critical Appraisal Checklist for Case Control Studies - modified

1. Yes
2. No
3. Unclear
4. Not reported
5. Not applicable

**Was performance on the cognitive task or clinical assessment tool measured in a standard, valid and reliable way?**

JBI Critical Appraisal Checklist for Case Control Studies - modified

1. Yes
2. No
3. Unclear
4. Not reported
5. Not applicable

**Were strategies to deal with confounding factors stated?**

JBI Critical Appraisal Checklist for Case Control Studies - modified

Typical confounders include baseline characteristics, prognostic factors, or concomitant exposures - A high quality study at the level of case control design will identify the potential confounders and measure them (where possible). JBI Critical Appraisal Checklist for Case Control Studies - modified

1. Yes
2. No
3. Unclear
4. Not reported
5. Not applicable

**Were methods used to prevent and address missing data stated?**

Check if they used visual inspection, motion correction etc.

Data to be collected based on

Cite this chapter as: Li T, Higgins JPT, Deeks JJ (editors). Chapter 5: Collecting data. In: Higgins JPT, Thomas J, Chandler J, Cumpston M, Li T, Page MJ, Welch VA (editors). Cochrane Handbook for Systematic Reviews of Interventions version 6.3 (updated February 2022). Cochrane, 2022. Available from www.training.cochrane.org/handbook.

1. Yes
2. No
3. Unclear
4. Not reported
5. Not applicable

**Was appropriate statistical analysis used?**

JBI Critical Appraisal Checklist for Case Control Studies - modified

1. Yes
2. No
3. Unclear
4. Not reported
5. Not applicable

**Was an appropriate and predetermined threshold probability used?**

JBI Critical Appraisal Checklist for Case Control Studies - modified

1. Yes
2. No
3. Unclear
4. Not reported
5. Not applicable
